# Emulator-based Bayesian optimization for efficient multi-objective calibration of an individual-based model of malaria

**DOI:** 10.1101/2021.01.27.21250484

**Authors:** Theresa Reiker, Monica Golumbeanu, Andrew Shattock, Lydia Burgert, Thomas A. Smith, Sarah Filippi, Ewan Cameron, Melissa A. Penny

## Abstract

Individual-based models have become important tools in the global battle against infectious diseases, yet model complexity can make calibration to biological and epidemiological data challenging. We propose a using a Bayesian optimization framework employing Gaussian process or machine learning emulator functions to calibrate a complex malaria transmission simulator. We demonstrate our approach by optimizing over a high-dimensional parameter space with respect to a portfolio of multiple fitting objectives built from datasets capturing the natural history of malaria transmission and disease progression. Our approach quickly outperforms previous calibrations, yielding an improved final goodness of fit. Per-objective parameter importance and sensitivity diagnostics provided by our approach offer epidemiological insights and enhance trust in predictions through greater interpretability.

## INTRODUCTION

### Individual-based models of infectious diseases

Over the last century, mathematical modelling has become an important tool to analyze and understand disease- and intervention-dynamics for many infectious diseases. Individual-based models (IBMs), where each person is simulated as an autonomous agent, are now widely used. These mathematical models capture heterogeneous characteristics and behaviors of individuals, and are often stochastic in nature. This bottom-up approach of simulating individuals and transmission events enables detailed, robust and realistic predictions on population epidemic trajectories as well as the impact of interventions such as vaccines or new drugs (*1, 2*). Going beyond simpler (compartmental) models to capture stochasticity and heterogeneity in populations, disease progression, and transmission, IBMs can additionally account for contact networks, individual care seeking behavior, immunity effects, or within-human dynamics (*1-3*). As such, well-developed IBMs provide opportunities for experimentation under relatively naturalistic conditions without expensive clinical or population studies. Prominent recent examples of the use of IBMs include assessing the benefit of travel restrictions during the Ebola outbreak 2014–2016 (*4*) and guiding the public health response to the Covid-19 pandemic in multiple countries (*5*). IBMs have also been applied to tuberculosis (*6*), influenza (*7*), dengue (*8*), and many other infectious diseases (*2*). Within the field of malaria, several IBMs have been developed over the last 15 years and have been used to support understanding disease and mosquito dynamics (*9-11*), predict the public health impact or carry out economic analyses of (new) interventions (*12-15*); and investigate drug resistance (*16*). Many have had wide-reaching impact, influencing WHO policy recommendations (*12, 17-19*) or strategies of national malaria control programs (*20*).

### Calibration caveats and the curse of dimensionality

For model predictions to be meaningful, modelers need to ensure their models accurately capture abstractions of the real world. The potential complexity and realism of IBMs often come at the cost of long simulation times and potentially large numbers of input parameters, whose exact values are often unknown. Parameters may be unknown because they represent derived mathematical quantities that cannot be directly measured or require elaborate, costly experiments (for example shape parameters in decay functions (*21*)), because the data required to derive them in isolation is incomplete or accompanied by inherent biases, or because they interact with other parameters.

Calibrating IBMs poses a complex high-dimensional optimization problem and thus algorithm-based calibration is required to find a parameter set that ensures realistic model behavior, capturing the biological and epidemiological relationships of interest. Local optima may exist in the potentially highly irregular, high-dimensional goodness-of-fit surface, making iterative, purely sampling-based algorithms (e.g. Particle Swarm Optimization or extensions of Newton-Raphson) inefficient and, in light of finite runtimes and computational resources, unlikely to find global optima. Additionally, the *curse of dimensionality* means the number of evaluations of the model scales exponentially with the number of dimensions (*22*). As an example, for the model discussed in this paper, a 23-dimensional parameter space at a sampling resolution of one sample per 10 percentile cell in each dimension, would yield 10^*number of dimensions*^ = 10^23^ cells. This is larger than number of stars in the observable Universe (of order 10^22^ (*23*)). Furthermore, most calibrations are not towards one objective or dataset. For multi-objective fitting, each parameter set requires the evaluation of multiple outputs and thus multiple simulations to ensure that all outcomes of interest are captured (in the model discussed here epidemiological outcomes such as prevalence, incidence, or mortality patterns).

In this study, we applied a new approach to calibrate a well-established and used IBM of malaria dynamics called *OpenMalaria*. Malaria IBMs in particular are often highly complex (e.g. containing multiple sub-modules and many parameters), consider a two-host system influenced by seasonal dynamics, and often account for multifaceted within-host dynamics. OpenMalaria features within-host parasite dynamics, the progression of clinical disease, development of immunity, individual care seeking behavior, vector dynamics and pharmaceutical and non-pharmaceutical antimalarial interventions at vector and human level (https://github.com/SwissTPH/openmalaria.wiki.git) (*3, 21, 24*). Previously, the model was calibrated using an asynchronous genetic algorithm (GA) to fit 23 parameters to 11 objectives representing different epidemiological outcomes, including age-specific prevalence and incidence patterns, age-specific mortality rates and hospitalization rates (*3, 21, 24*) (see Supplementary Texts 1 and 2 for details on the calibration objectives and data). However, the sampling-based nature and sequential function evaluations of GAs can be too slow for high-dimensional problems in irregular spaces where only a limited number of function evaluations are possible and valleys of neutral or lower fitness may be difficult to cross (*25*), (*26*).

Other solutions to fit similarly detailed IBMs of malaria employ a combination of directly extracting parameter values from the literature where information is available, and fitting the remainder using multi-stage, modular Bayesian Markov Chain Monte Carlo (MCMC)-based methods (*27-32*). For these models, multiple fitting objectives are often not addressed simultaneously. Rather, to our knowledge, most other malaria IBMs are divided into functional modules (such as the human transmissibility model, within-host parasite dynamics model, and the mosquito or vector model), which are assumed to be influenced by only a limited number of parameters each. The modules are then fit independently and in a sequential manner (*28-32*). Modular approaches reduce the dimensionality of the problem, allowing for the use of relatively straightforward MCMC algorithms. However, these struggle with efficiency in high dimensions as their Markovian nature requires many sequential function evaluations (10^4^–10^7^ even for simple models), driving up computing time and computational requirements (*33*). Additionally, whilst allowing for the generation of posterior probability distributions of the parameters (*31*), the modular nature makes sequential approaches generally unable to account for interdependencies between parameters assigned to different modules and how their co-variation may affect disease dynamics.

### Emulators and Bayesian Optimization

Progress in recent years on numerical methods for supervised, regularized learning of smooth functions from discrete training data allows us to revisit calibration of detailed mathematical models using Bayesian methods for global optimization (*34*). Current state-of-the art calibration approaches for stochastic simulators are often based around Kennedy and O’Hagan’s approach (*35*) (KOH), where a posterior distribution for the calibration parameters is derived through a two-layer Bayesian approach involving cascade of surrogates (usually GPs) (*36*). A first GP is used to model the systematic deviation between the simulator and the real process it represents, while a second GP is used to emulate the simulator (*37*). However, this approach is computationally intense when scaling to high-dimensional input spaces and multi-objective optimization. A fully Bayesian KOH approach is likely computationally heavy (*37*) for the efficient calibration of detailed malaria simulators like OpenMalaria. Single-layer Bayesian optimization with Gaussian processes (GPs) on the other hand have gained popularity as an efficient approach to tackle expensive optimization problems, for example in hyperparameter search problems in machine learning (*38, 39*). Assuming that the parameter-solution space exhibits a modest degree of regularity, a prior distribution is defined over a computationally expensive objective function by the means of a light-weight probabilistic emulator such as a Gaussian process. The constructed emulator is sequentially refined by adaptively sampling the next training points based on acquisition functions derived from the posterior distribution. The trained emulator model is used to make predictions over the objective functions from the input space with minimum evaluation of the expensive *true* (simulator) function. Purely sampling-based iterative approaches (like genetic algorithms) are usually limited to drawing sparse random samples from proposals located nearby existing samples in the parameter space. In contrast, the use of predictive emulators permits exploration of the entire parameter space at higher resolution. This increases the chances of finding the true global optimum of the complex objective function in question and avoiding local optima.

Here, we use a single-layer Bayesian optimization approach to solve the multidimensional, multi-objective calibration of OpenMalaria (Fig. 1). Employing this single-layer Bayesian approach further allows for the direct comparison to previous calibration attempts for OpenMalaria as the objective functions are retained. We prove the strength and versatility of our approach by optimizing its 23 input parameters using real-world data on 11 epidemiological outcomes in parallel. To emulate the solution space, we explore and compare two prior distributions, namely a GP emulator and a *superlearning* algorithm in form of a Gaussian process stacked generalization (GPSG) emulator. We first use a GP emulator to emulate the solution space. Whilst GP emulators provide flexibility whilst retaining relative simplicity (*39*) and have been used previously as priors in Bayesian optimization (*38*), stacked generalization algorithms have not. They provide a potentially attractive alternative as they have been shown to outperform GPs and other machine learning algorithms in capturing complex spaces (*14, 40*). The stacked generalization algorithm (*40*) builds on the idea of creating ensemble predictions from multiple learning algorithms (*level 0 learners*). The cross-validated predictions of the level 0 learners are incorporated into a general learning system (level 1 meta-learner). This allows for the combination of memory-efficient and probabilistic algorithms in order to reduce computational time, whilst retaining probabilistic elements required for adaptive sampling. Here, we showcase the efficiency and speed of the Bayesian optimization calibration scheme and propose a novel modus operandi to parameterize complex mathematical models that harvests recent computational developments and is scalable to high dimensions in multi-objective calibration.

**Fig. 1.**
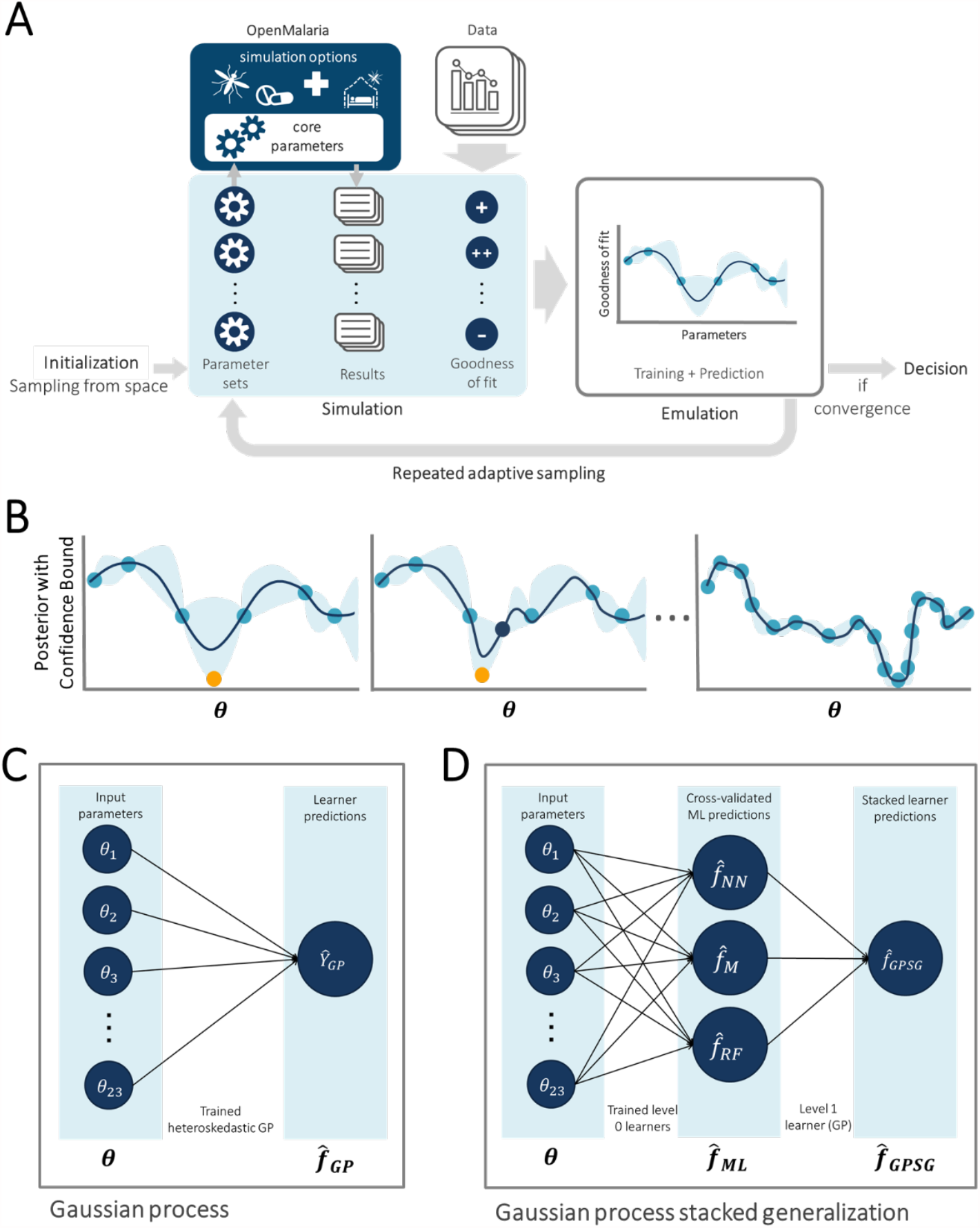
Overview of model calibration approaches by Bayesian optimization using Gaussian process and machine learning emulators. **A. General Framework.** The input parameter space is initially sampled in a space-filling manner, generating the initial core parameter sets (initialization). For each candidate set, simulations are performed with the model, mirroring the studies that yielded the calibration data. The deviation between simulation results and data is assessed, yielding goodness of fit scores for each parameter set. An emulator (C or D) is trained to capture the relationship between parameter sets and goodness of fit and used to generate out-of-sample predictions. Based on these, the most promising additional parameter sets are chosen (adaptive sampling by means of an acquisition function), evaluated, and added to the training set of simulations. Training and adaptive sampling are repeated until the emulator converges and a decision on the parameter set yielding the best fit is made. **B. Acquisition Function**. The acquisition function is used to determine new parameter space locations. Thus, ***θ*** is a vector of input parameters (23-dimensional for the model described here) to be evaluated during adaptive sampling. It incorporates both predictive uncertainty of the emulator and proximity to the minimum. **C. Gaussian process emulator**. A heteroscedastic Gaussian process is used to generate predictions on the loss functions, 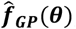, for each input parameter set ***θ*. D. Gaussian process stacked generalization emulator**. Three machine learning algorithms (level 0 learners: bilayer neural net, multivariate adaptive regression splines and random forest) are used to generate predictions on the individual objective loss functions 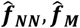 and 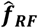 (collectively 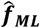) at locations ***θ***. These predictions are inputs to a heteroscedastic (level 1 learner) which is used to generate the stacked learner predictions 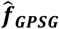 and derive predictions on the overall goodness of fit 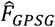.

## RESULTS

### Calibration workflow

The developed model calibration workflow approach is summarized in Fig. 1A. In brief, goodness of fit scores were first derived for randomly generated, initial parameter sets. The goodness of fit scores were defined as a weighted sum of the loss functions for each of 11 fitting objectives. These span various epidemiological measures capturing the complexity and heterogeneity of the malaria transmission dynamics, including the age-prevalence and age-incidence relationships, and are informed by a multitude of observational studies (see methods and Supplementary Text 2). Next, GP and GPSG emulators were trained on the obtained set of scores and used to approximate the relationship between parameter sets and goodness of fit for each objective. After initial investigation of different machine learning algorithms, the GPSG was constructed using a bilayer neural net, multivariate adaptive regression splines and random forest as *level 0* learners and a heteroscedastic Gaussian process as *level 1* learner (Fig. 1C-D, see methods and supplement). Using a lower confidence bound acquisition function based on the emulators’ point and uncertainty predictions for proposed new candidate parameter sets, the most promising sets were chosen. These parameter sets were simulated and added to the database of simulations for the next iteration of the algorithm. At the next iteration, the emulators are re-trained on the new simulation database and re-evaluated (Fig. 1B). This iterative process of simulation, training and emulation was repeated until a memory limit of 1024GB was hit. Approximately 130,000 simulations were completed in up to this point.

### Algorithm performance by iteration and time and convergence

Both emulators adequately captured the input-output relationship of the calculated loss-functions from the simulator, with better accuracy when close to minimal values of the weighted sum of the loss functions, *F* (Fig. 2A). This is sufficient as the aim of both emulators within the Bayesian optimization framework is to find minimal loss function values rather than an overall optimal predictive performance for all outcome values. Examples of truth vs predicted estimates on a 10% holdout set are provided in Fig. 2A (additional plots for all objectives can be found in Supplementary Fig.s S2-S5). A *satisfactory fit* of the simulator was previously defined by a loss function value of *F* = 73.2 (*21*). The *previous best* model fit derived using the GA had a weighted sum of the loss functions of *F* = 63.7 (*21*). *Satisfactory fit* was achieved by our approach in the first iteration of the GPSG-based Bayesian optimization algorithm (GPSG-BO), and after six iterations for the GP-based algorithm (GP-BO) (Fig. 2B). The *current best* fit was approximately retrieved after six iterations for the GPSG-BO algorithm and after nine iterations for GP-BO, and was improved by both algorithms after ten iterations (returning final values *F* = 58.3 for GP-BO and 59.6 for GPSG-BO). This shows that the Bayesian optimization approach with either of our emulators very quickly achieves a better simulator fit than obtained with a classical GA approach that was previously employed to calibrate OpenMalaria. Of the two emulators, the GP approach finds a parameter set associated with a better overall accuracy and the GPSG reaches *satisfactory* values faster (both in terms of iterations and time). A likely explanation for this is that the GPSG-BO is unable to propagate its full predictive variance into the acquisition function. Only uncertainty stemming from the level 1 probabilistic learner (GP) is therefore captured in the final prediction. This leads to underestimation of the full predictive variance, and a bias towards exploitation in the early stages of the GPSG-BO algorithm (as illustrated by early narrow sampling, see Supplementary Fig.s S6-7).

**Fig. 2.**
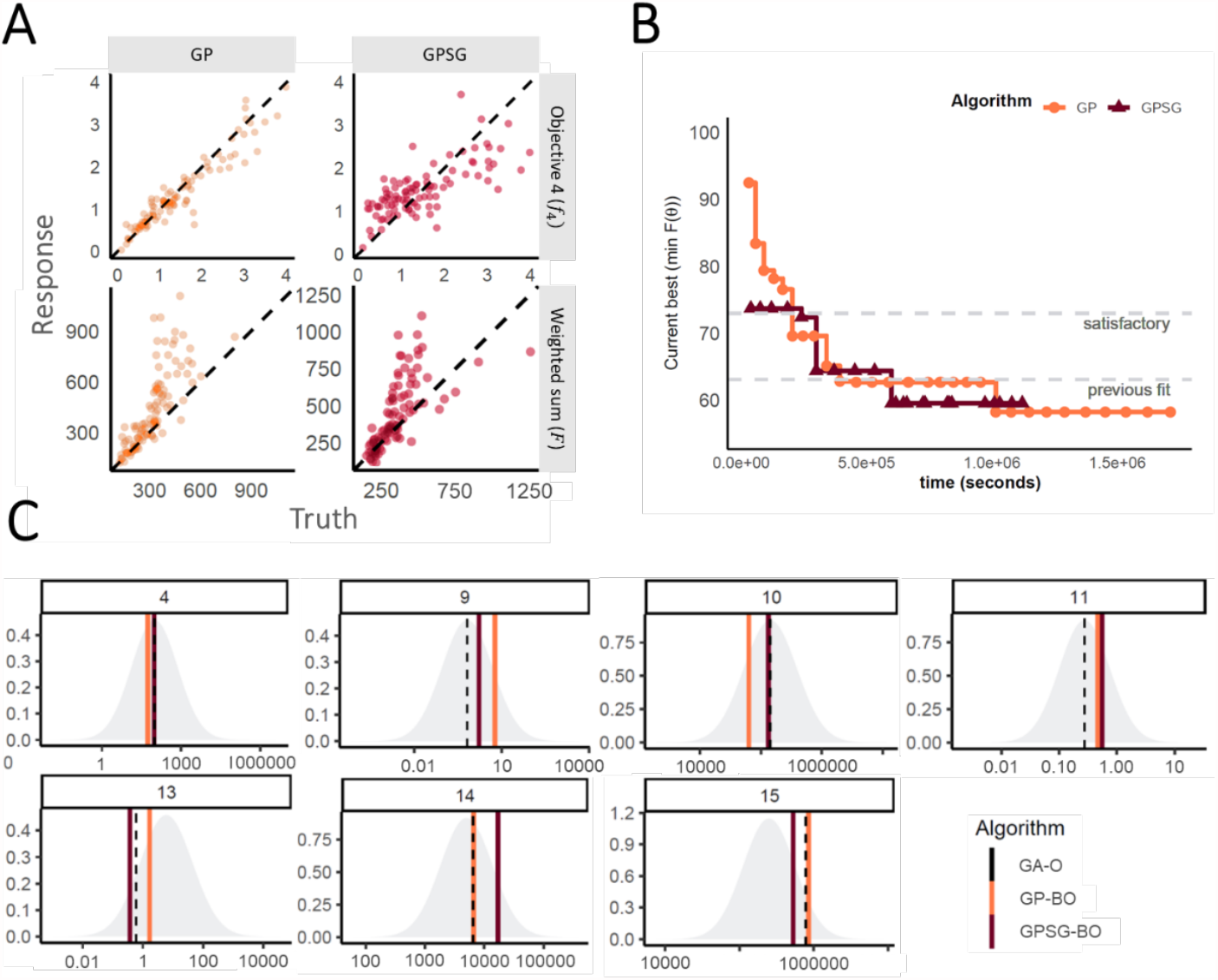
Emulator performance. **A. Example of emulator predictions vs true values on a 10% holdout set.** Predictions are shown for the final iteration of each optimization (iteration 30 for GP-BO and iteration 23 for GPSF-BO). Here, emulator performances are shown for objective 4 (the age-dependent multiplicity of infection,*f*_4_) and the weighted sum *F*. Plots for all other objectives are provided in the supplement. GP = Gaussian process emulator, GPSG = Gaussian process stacked generalization emulator **B. Convergence**. Weighted sum of loss functions over 11 objectives associated with the current best fit parameter set by time in seconds. Satisfactory fit of OpenMalaria refers to a weighted sum of loss functions value of 73.2 (as defined previously (*21*)). Previous best fit for OpenMalaria was achieved by the genetic algorithm had a loss function value of 63.7. Our new approach yields a fit of 58.2 for GP-BO in iteration 21 within in 1.02*^6^ seconds (∼12 days) and 59.6 for GPSG-BO in iteration 10 in 6.00e^5^ seconds (∼7 days). GP-BO = Gaussian process emulator Bayesian optimization, GPSG-BO = Gaussian process stacked generalization emulator Bayesian optimization). **C. Example log prior parameter distributions and posterior estimates**. The most influential parameters on the weighted sum of the loss functions are shown here (see Fig. 3C). All other plots can be found in the supplement. The posterior estimates for GP-BO and GPSG-BO are shown in relation to those previously derived through optimization using a genetic algorithm (GA-O)

Fig. 2C shows examples of the posterior estimates returned by the optimization algorithms in context of the log prior distributions for the parameters with the greatest effects on *F* (see also Fig. 3C). All algorithms return parameter values within the same range and (apart from parameter 4), clearly distinct from the prior mean. The fact that highly similar parameter values are identified by multiple algorithms strengthens confidence in the final parameter sets yielded by the algorithms.

**Fig. 3.**
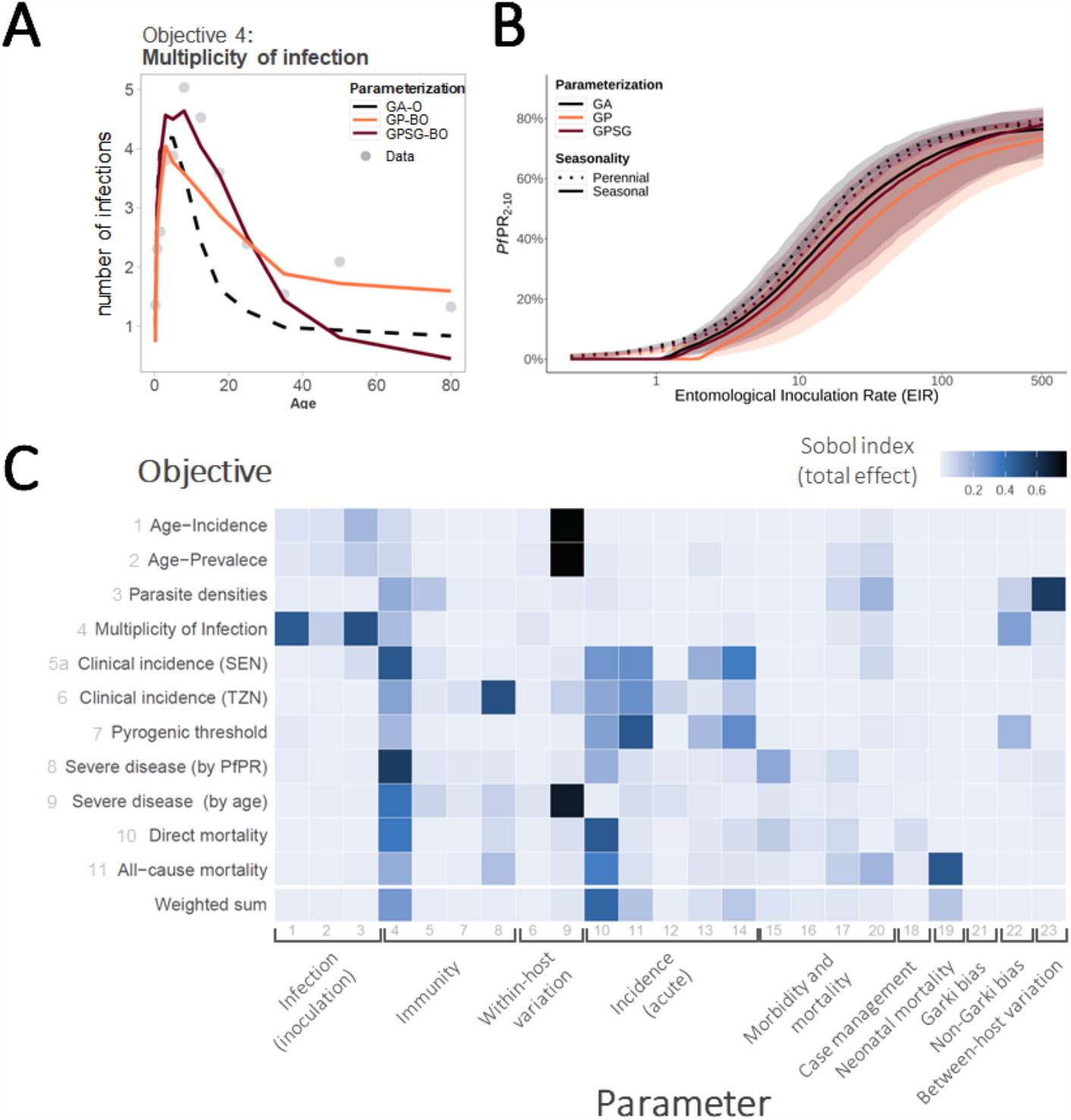
**A. Multiplicity of infection by age.** Comparison of simulator goodness of fit for objective 4, the age-specific multiplicity of infection (number of genetically distinct parasite strains concurrently present in one host). Simulations were carried out for the same random seed for all parameterizations and for a population size of N=5,000. **B. Simulated epidemiological relationship between transmission intensity (entomological inoculation rate, EIR) and P. falciparum prevalence (PfPR**_**2-10**_). Simulated epidemiological relationship between the transmission intensity (EIR in number of infectious bites per person per year) and infection prevalence in individuals aged 2-10 years (PfPR_2-10_) under the parameterizations achieved by the different optimization algorithms. Lines show the mean across 100 random seed simulations for a simulated population size N=10,000 and the shaded area shows the 95% confidence interval. **C. Parameter effects on the objective variance**. Using the GP emulator, a global sensitivity analysis (Sobol analysis) was conducted. The tile shading shows the total effect indices for all objective functions and parameters grouped by function. SEN= Senegal, TZN = Tanzania.

### Optimal Goodness of Fit

The best fit parameter sets yielded by our approach are provided in the supplement (Table S2). Importantly, after ten iterations of the GPSG-BO algorithm (approximately 7 days), and 20 iterations for the GP-BO algorithm (approximately 12 days), both approaches yielded similar values of the 11 objective loss functions, along with similar weighted total loss function values, and qualitatively similar visual fits and predicted trends to the data (Fig. 3A-B and supplement). We found this to be an unexpectedly fast result of the two algorithms. Details of the algorithm’s best fits to the disease and epidemiological data are shown in Supplementary Fig.s S8-S18. Overall, several objectives had visual and reduced loss-function improvements, for example to the objective on the multiplicity of infection (Fig. 3A).

### Impact / Parameter sensitivity analysis & External validation

An additional benefit of using emulators is the ability to understand the outcome’s dependence on and sensitivity to the input parameters. To identify the most influential parameters for each of the 11 fitting objectives, we used the GP emulator trained on all available training simulation results from the optimization process (R^2^=0.53 [objective 7] - 0.92 [objective 3]) to conduct a global sensitivity analysis by variance decomposition (here via Sobol analysis (*41*)). Fig. 3C shows Sobol total effect indices quantifying the importance of individual parameters and describing each parameter’s contributions to the outcome variance for each objective. Our results indicate that most objectives are influenced by multiple parameters from different groups, albeit to varying degrees, thus highlighting the importance of simultaneous multi-objective fitting. Clusters of influential parameters can be observed for most objectives; for example, parameters associated with incidence of acute disease influence clinical incidence and pyrogenic threshold objectives. Some parameters have strong influence on multiple objectives, such as parameter 4, the critical value of cumulative number of infections and influences immunity acquisition; and parameter 10, a factor required to determine the pyrogenic threshold, which we find to be a key parameter determining infections progressing to clinical illness.

### Algorithm validation

In order to test if our algorithms can recover a known solution, the final parameter sets for both approaches were used to generate synthetic field data sets, and our approaches were subsequently applied to recover the known parameter set. For the GP, 13 of the 23 parameters were recovered (Supplementary Fig. S19A). Those not recovered largely represented parameters to which the weighted loss function was found to be insensitive (Fig. 3C). Thus, rather than showing a shortcoming of the calibration algorithm, this suggests a potential for dimensionality reduction of the simulator and re-evaluation of its structure.

### Comparison of key epidemiological relationships and implications for predictions

The new parameterizations for OpenMalaria were further explored to assess key epidemiological relationships, in an approach similar multiple-model comparison in Penny et al. 2016 (*12*). We examined incidence and prevalence of disease, as well as incidence of mortality for multiple archetypical settings, considering a range of perennial and seasonal transmission intensity and patterns. The results are presented in Fig. 3B and Supplementary Fig.s S20-30. The new parameterizations result in increased predicted incidence of severe episodes and decreased prevalence for all transmission intensities (thus also slightly modifying the prevalence-incidence relationship). While we found that the overall implications for the other simulated epidemiological relationship were small, the differences in predictions for severe disease may carry important implications for public health decision making. We conclude that our new parameterizations do not fundamentally bring into question previous research conducted using OpenMalaria, but we do suggest re-evaluation of adverse downstream events such as severe disease and mortality.

## DISCUSSION

Calibrating individual-based models can be challenging as many techniques struggle with high dimensionality, or become infeasible with long model simulation times and multiple calibration objectives. However, ensuring adequate model fit to key data is vital, as this impacts the weighting, we should give model predictions in the public health decision making process. The Bayesian optimization approaches presented here provide fast solutions to calibrating individual-based models while improving model accuracy, and by extension prediction accuracy.

Using a Bayesian optimization approach, we calibrated a detailed simulator of malaria transmission and epidemiology dynamics with 23 input parameters simultaneously to 11 epidemiological outcomes, including age-incidence and -prevalence patterns. The use of a probabilistic emulator to predict goodness-of-fit, rather than conducting sparse sampling, allows for cheap evaluation of the simulator at many locations and increases our confidence that the final parameter set represents a global optimum. Our approach provides a fast calibration whilst also providing a better fit compared to the previous parameterization. We are further able to define formal endpoints to assess calibration alongside *visual confirmation* of goodness of fit (*21, 28*), such as the emulator’s predictive variance approaching the observed simulator variance. The emulator’s ability to quantify the input stochasticity of the simulator also enables simulation at small population sizes, contributing to fast overall computation times.

Despite the demonstrated strong performance of stacked generalization in other contexts such as geospatial mapping (*14, 40, 42-45*), we found that using a *superlearning* emulator for Bayesian optimization was not superior to traditional GP-based methods. In our context using GPSG sped up convergence of the algorithm, but both approaches, GP and GPSG, led to equally good fits. Each approach does however, have different properties with context-dependent benefits: The dimensionality reduction provided by GPSG approaches may lead to computational savings depending on the *level 0* and *level 1* learners. At the same time, only level 1 learner uncertainty is propagated into the final’ ’ predictions, which affects the efficacy of adaptive sampling and may lead to overly exploitative behavior, where sampling close to the point estimate of the predicted optimum is overemphasized, rather than exploring the entire parameter space (see supplements S2 and S3 on selected points). On the other hand, exploration/exploitation trade-offs for traditional GP-BO algorithms have long been examined and *no regret* solutions have been developed (*46*).

The methodology presented here constitutes a highly flexible framework for individual based model calibration and aligns with the recent literature on using emulation in combination with stochastic computer simulation experiments of infectious diseases (*47*). Both algorithms can be applied to other parameterization and optimization problems in disease modelling and also in other modelling fields, such as physical or mobility and transport models. Furthermore, in the GPSG approach, additional or alternative level 0 can be easily incorporated. Possible extensions to our approach include combination with methods to adaptively reduce the input space for constrained optimization problems (*48*), or other emulators may be chosen depending on the application. For example, homoscedastic GPs, which are faster than the heteroscedastic approach presented here, may be sufficient for many applications (but not for our IBM in which heteroscedastic was required due to the stochastic nature of the model).

Alternatively, the computational power required by neural net algorithms scales only linearly (compared with a nominal cubic scaling for GPs) with the sample size, and we envisage wide applications for neural net-based Bayesian optimization in high dimensions. In our example, the bilayer neural net algorithm completed training and prediction within seconds whilst maintaining very high predictive performance. Unfortunately, estimating the uncertainty required for good acquisition functions is difficult in neural networks, but solutions are being developed (*39, 49*). These promising approaches should be explored as they become more widely available in high-level programming languages. With the increased availability of code libraries and algorithms, Bayesian optimization with a range of emulators is also becoming easier to implement.

The probabilistic, emulator-based calibration approach is accompanied by many benefits, including relatively quick global sensitivity analysis. As explored in this work, GP-based methods are easily coupled with sensitivity analyses, which provide detailed insights into a model’s structural dependencies and the sensitivity of its goodness of fit to the input parameters. To the best of our knowledge, no other individual-based model calibration study has addressed this. In the case of malaria models, we have shown the interdependence of all OpenMalaria model components and a relative lack of modularity. In particular, within-host immunity-related parameters were shown to influence all fitting objectives, including downstream events such as severe disease and mortality when an infection progresses to clinical disease. Thus, calibrating within-host immunity in the absence of key epidemiology and population outcomes can lead to suboptimal calibration and ultimate failure of the model to adequately capture disease biology and epidemiology.

We have employed a different approach to calibrating OpenMalaria compared with previous methods but reach broadly similar comparisons to the natural history of disease. We also attainted a slightly improved but similar goodness of fit, the main benefit being improved fitting times and the ability to measure parameter importance. Given the high number of influential parameters for each epidemiological objective in our parameter importance investigations, and the overlap between parameter-objective associations, we argue that, where possible, multi-objective fitting should be preferred over purely sequential approaches. Our approach confirms that using a parallel approach to parameterization rather than a modular, sequential, one captures the joint effects of all parameters and ensures that all outcomes are simultaneously accounted for. To the best of our knowledge, no model of malaria transmission of comparable complexity and a comparable number of fitting objectives was simultaneously calibrated to all its fitting objectives. Disregarding the joint influence of *all* parameters on the simulated outcomes may negatively impact the accuracy of model predictions, in particular on policy-relevant outcomes of severe disease and mortality.

Despite providing relatively fast calibration towards a better fitting parameter set, several limitations remain in our work. We have not systematically tested that a global optimum has been reached in our new approach, but assume it is close to a global minimum for the current loss-functions defined, as further iterations did not yield changes, and both the GP and GPSG achieved similar weighted loss function and parameter sets. We aimed to improve the algorithm to calibrate detailed IBM, but we did not incorporate new data, which will be important moving forward as our parameter importance and validation analysis highlights several key epidemiological outcomes on severe disease and mortality are sensitive to results.

The key limitations of Bayesian optimization, particularly when using a Gaussian process emulator, are the high computational requirements in terms of memory and parallel computing nodes due to increasing runtimes and cubically scaling memory requirements of GPs. For this reason, we opted to not employ fully Bayesian KOH methods, which would double the number of GPs that would need to be run. Yet, memory limits may be reached before the predictive variance approached its limit. Furthermore, we chose an acquisition function with high probability to be *no regret* (*46*), but this likely overemphasizes exploration in the early stages of the algorithm considering the dimensionality of the problem and finite runtime. We opted here for pure exploitation every 5 iterations, but a more formal optimization of the acquisition function should be explored. The GPSG approach presented here can partially alleviate this challenge, depending on the choice of learning algorithms, but the iterative nature and need for many simulations remain. Memory- and time-saving extensions are thus worth exploring, such as incorporating GPU computing or adaptively constraining the prior parameter space, dimensionality reduction, or addressing alternative acquisition functions. Additionally, as with all calibration methodologies, many choices are left to the user, such as the size of the initial set of simulations, the number of points added per iteration, or the number of replicates simulated at each location. There is no general solution to this as the optimal choices are highly dependent on the problem at hand, and we did not aim to optimize these. Performance might be optimized further through a formal analysis of all these variables, however the methodology here is already fast, effective, and highly generalizable to different types of simulation models and associated optimization problems. Improving the loss-functions or employing alternative *Pareto front* efficiency algorithms was not the focus of our current study but would be a natural extension of our work, as would be alternative approaches to the weighting of objectives, which remains a subjective component of multi-objective optimization problems (*50*).

A model’s calibration to known input data forms the backbone of its predictions. The workflow presented here provides great advances in the calibration of detailed mathematical models of infectious diseases such as IBMs. Provided sufficient calibration data to determine goodness-of-fit, our approach is easily adaptable to any agent-based model and could become the new modus operandi for multi-objective, high-dimensional calibration of stochastic disease simulators.

## METHODS

### Preparation of calibration data and simulation experiments

Disease transmission models generally have two types of parameter inputs: core parameters, inherent to the disease and determining how its natural history is captured, and simulation options characterizing the specific setting and the interventions in place (Fig. 1A in the main manuscript). The simulation options specify the simulation context such as population demographics, transmission intensity, seasonality patterns and interventions and typically vary depending on the simulation experiment. In contrast, the core parameters determine how its epidemiology and aetiopathogenesis are captured. These include parameters for the description of immunity (e.g. decay of maternal protection), or for defining clinical severe episodes (e.g. parasitemia threshold). To inform the estimation of core parameters, epidemiological data on the natural history of malaria extracted from published literature and collated in previous calibrations of OpenMalaria(*3, 21, 24*) were re-used in this calibration round. These include demographic data such as age-stratified numbers of host individuals which are used to derive a range of epidemiological outcomes such as age-specific prevalence and incidence patterns, mortality rates and hospitalization rates.

Site-specific OpenMalaria simulations were prepared, representing the studies that yielded these epidemiological data in terms of transmission intensity, seasonal patterns, vector species, intervention history, case management, and diagnostics (*24*). The mirroring of field study characteristics in the simulation options ensured that any deviation between simulation outputs and data could be attributed to the core parameters. Age-stratified simulation outputs to match to the data include numbers of host individuals, patent infections, and administered treatments. A summary of the data is provided in the Supplementary text 2.

### General Bayesian Optimization framework with emulators

In our proposed Bayesian optimization framework (Fig. 1) we evaluated the deviation between simulation outputs and the epidemiological data by training probabilistic emulator functions that approximate the relationship between core parameter sets and goodness of fit. To test the optimization approach in this study we considered the original goodness of fit metrics for OpenMalaria detailed in (*21*) and in Supplementary Text 2, which uses either Residual Sum of Squares (RSS) or negative log-likelihood functions depending on the epidemiological data for each objective (*21, 24*). The objective function to be optimized is a weighted sum of the individual objectives’ loss functions.

We adopted a Bayesian optimization framework where a probabilistic emulator function is constructed to make predictions over the loss functions for each objective from the input space, with a minimum amount of evaluations of the (computationally expensive) simulator.

We compared two emulation approaches. Firstly, a heteroskedastic Gaussian process (GP) emulator and secondly a stacked generalization emulator (*40*). For approach 1 (*GP-BO*), we fitted a heteroskedastic Gaussian process with the input noise modelled as another Gaussian process (*51*) with a Matérn 5/2 kernel to account for the high variability in the parameter space (Fig. 1C) (*38, 52*). For approach 2 (*GPSG-BO*), we selected a two-layer neural network (*53-55*), multivariate adaptive regression splines (*56*), and a random forest algorithm (*57, 58*) as level 0 learners.

With each iteration of the algorithm, the training was extended using adaptive sampling based on an acquisition function (*lower confidence bound*) that accounts for uncertainty and predicted proximity to the optimum of proposed locations (Fig. 1B). As the emulator performance improves (as assessed by its predictive performance on the test set) we gain confidence in the currently predicted optimum.

### Malaria transmission and disease simulator

We applied our novel calibration approach to OpenMalaria (https://github.com/SwissTPH/openmalaria), an open-source modelling platform of malaria epidemiology and control. It features several related individual-based stochastic models of *P. falciparum* malaria transmission and control. Overall, the OpenMalaria IBM consists of a model of malaria in humans linked to a model of malaria in mosquitoes and accounts for individual level heterogeneity in humans (in exposure, immunity, and clinical progression) as well as aspects of vector ecology (e.g. seasonality and the mosquito feeding cycle). Stochasticity is featured by including between- and within-host stochastic variation in parasite densities with downstream effects on immunity (*24*). OpenMalaria further includes aspects of the health system context (e.g. treatment seeking behavior and standard of care) (*3, 24*) with additional probabilistic elements such as treatment seeking probabilities or the option for stochastic results of diagnostic tests. An ensemble of OpenMalaria model alternative variants is available defined by different assumptions about immunity decay, within-host dynamics, heterogeneity of transmission, along with more detailed sub-models that track parasite genetics, and pharmacokinetic and pharmacodynamics. The models allow for the simulation of interventions, such as the distribution of insecticide treated nets (ITNs), vaccines, or reactive case detection (*59, 60*), in comparatively realistic settings. Full details of the model and the history of calibration can be found in the original publications (*3, 21, 24*) and are summarized in Supplementary Texts 1 and 2. In our application, we use the term *simulator* to refer to the OpenMalaria base model variant (*21*).

### Calibrating OpenMalaria: loss functions and general approach

#### Aim

Let ***f*** (***θ***) denote a vector of loss functions obtained by calculating the goodness of fit between simulation outputs and the real data (full details of loss function can be found in supplementary Text 2). In order to ensure a good fit of the model, we aim to find the parameter set ***θ*** that achieves the minimum of the weighted sum of 11 loss functions (corresponding to the 10 fitting objectives) 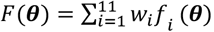, where *f*_*i*_(***θ***) is the value of objective function *i* at ***θ*** and *w*_*i*_ is the weight assigned to objective function *i*:

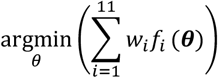

The weights are kept consistent with previous rounds of calibration and chosen such that different epidemiological quantities contributed approximately equally to *F*(***θ***) (see Supplementary Text 2).

#### Step 1: Initialization

Let *D* = 23 denote the number of dimensions of the input parameter space **Θ** and *W* = 11 the number of objective functions *f*_*i*_ (*θ*) *i* = 1, …, 11. Prior distributions consistent with previous fitting runs (*21*) were placed on the input parameters. As each parameter is measured in different units, we sampled from the *D* -dimensional unit cube **Θ** and converted these to quantiles of the prior distributions (*21*) (Supplementary Text 2and Supplementary Fig. S6). Previous research suggests that in high-dimensional spaces quasi-Monte Carlo (qMC) sampling outperforms random or Latin Hypercube designs for most function types and leads to faster rates of convergence (*61, 62*). We therefore used Sobol sequences to sample 1,000 initial locations from **Θ**. The GP can account for input stochasticity of the simulator. For each sample, we simulated 2 random seeds at a population size of 10,000 individuals. Additionally, 100 simulations were run at the centroid location of the unit cube to gain information on the simulator noise. Using small noisy simulations with small populations speeds up the fitting as the noisy simulations are less computational expensive than larger population runs. Replicates were used to detect signals in noisy settings and estimate the pure simulation variance (*51*). The 2000 unique locations were randomly split into a training set (90%) and a test set (10%). All simulator realizations at the centroid were added to the training set.

#### Step 2: Emulation

##### 2.1: Emulator Training

Each emulator type for each objective function was trained in parallel to learn the relationships between the normalized input space **Θ**, and the log-transform of the objective functions *f* (***θ***). In each dimension *d* ∈ *D*, the mean *ε*_*d*_ and standard deviation *σ*_*d*_ of the training set were recorded, *d* = 1, …, 23.

##### 2.2 Posterior prediction

We randomly sampled 500,000 test locations in **Θ** from a multivariate normal distribution with mean ***θ***_***opt***_ and covariance matrix **Σ**, where ***θ***_***opt***_ is the location of the current best location and **Σ** is determined based on previously all sampled locations, and scaled each dimension to mean *ε*_*d*_ and standard deviation *σ*_*d*_. The trained emulators were used to make predictions 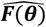 of the objective functions ***F*** (***θ***) at the test locations. Mean estimates, standard deviations, and nugget terms were recorded. The full predictive variance at each location ***θ*** ∈ **Θ** corresponds to the sum of the standard deviation and nugget terms. From this, we derived the weighted sum 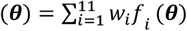, using weights *w* consistent with previous fitting runs (Smith 2012) with greater weighting for further downstream objectives. The predicted weighted loss function at location ***θ*** was denoted 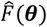 with a predicted mean 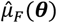 and variance 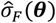. Every 15 iterations, we increase the test location sample size to 5 Million to achieve denser predictions.

#### Step 3: Acquisition

We chose the lower confidence bound (LCB) acquisition function to guide the search of the global minimum (*63*). Lower acquisition corresponds to *potentially* low values of the weighted objective function, either because of a low mean prediction value or large uncertainty (*64*). From the prediction set at iteration *t*, we sample without replacement 250 new locations 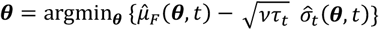, with the hyperparameter *v* = 1 and 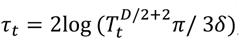, where *T*_*t*_ is the number of previous unique realisations of the simulator at iteration *t*, and *δ* = 0.01 is a hyperparameter (*46*). We choose this method as with high probability it is *no regret (46, 64)*. With increasing iterations, confidence bound-based methods naturally transition from mainly exploration to exploitation of the current estimated minimum. In addition to this, we force exploitation every 10 iterations by setting *τ*_*t*_ = 0).

#### Step 4: Simulate

The simulator was evaluated at locations identified in step 3 and the realisations were added to the training set. Steps 2-4 were run iteratively. The Euclidian distance between locations of current best realisations was recorded.

#### Step 5: Convergence

Convergence was defined as no improvement in the best realisation, argmin_***F***_***F***.

### Emulator definition

We compared two emulation approaches. Firstly, a heteroskedastic GP emulator and secondly a stacked generalization emulator (*40*) using a two-layer neural net, multivariate adaptive regression splines (MARS) and a random forest as level 0 learners and a heteroskedastic GP as level 1 learner:

#### 1.1.1 Heteroskedastic Gaussian Process (hetGP) (*65*)

We fitted a Gaussian process with the input noise modelled as another Gaussian process (*51*). After initial exploration of different kernels, we chose a Matérn 5/2 kernel to account for the high variability in the parameter space. A Matérn 3/2 correlation function was also tested performed equally. Each time the model was built (for each objective at each iteration), its likelihood was compared to that of a homoscedastic Gaussian process and the latter was chosen if its likelihood was higher. This resulted in a highly flexible approach, choosing the best option for the current task.

#### 1.1.2 Gaussian Process Stacked Generalization (GPSG)

Stacked generalization was first proposed by Wolpert 1992 (*40*) and builds on the idea of creating ensemble predictions from multiple learning algorithms (level 0 learners). In *superlearning*, the cross-validated predictions of the level 0 learners are fed into a level 1 meta-learner. We compared the 10-fold cross-validated predictive performance of twelve machine learning algorithms on the test set. All algorithms were accessed through the mlr package in R (*66*). We compared two neural network algorithms (brnn (*54*) for a two layer neural network and nnet for a single-hidden-layer neural network (*67*)), five regression algorithms (cvglmnet (*68*) for a generalised linear model with LASSO or Elasticnet Regularization and 10-fold cross validated lambda, glmboost (*69*) for a boosted generalized linear model, glmnet (*68*) for a regular GLM with Lasso or Elasticnet Regularisation, mars for multivariate adaptive regression splines (*70*), and cubist for rule-and instance-based regression modelling (*71*)), three random forest algorithms (randomForest (*58*), randomForestSRC (*72*) and ranger (*73*)), and a tree-like node harvesting algorithm (nodeHarvest (*74*)). Extreme gradient boosting and support vector regression were also tested but excluded from the comparison due to its long runtime. Their performance was compared with regards to runtimes, and correlation coefficients between predictions on the test set and the true values. Based on these, we selected the two-layer neural network (brnn) (*55*), multivariate adaptive regression spline (mars) (*70*), and random forest (randomForest) (*58*) algorithms. This ensemble of machine learning models constituted the level 0 learners and was fitted to the initialization set. Out-of-sample predictions from a 10-fold cross validation of each observation were used to fit the level 1 heteroskedastic Gaussian process. As in approach 1, we opted for a Matérn 5/2 kernel and retained the option of changing to a homoscedastic model where necessary.

### Emulator performance

We ascertained that both emulators captured the input-output relationship of the simulator by tracking the correlation between true values ***f*** and predicted values 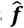on the holdout set of 10% of initial simulations with each iteration (truth vs predicted R2 0.51-0.89 for GP vs 0.37-0.77 for GPSG after initialization, see supplement S1). Transition from exploration to exploitation during adaptive sampling was tracked by recording the distribution of points selected during adaptive sampling in each iteration (Supplementary Fig.s S2 and S3).

### Sensitivity analysis

A global sensitivity analysis was conducted on a heteroskedastic GP model with Matérn 5/2 kernel that was trained on all training simulation outputs (n=5,400) from the fitting process. We used the Jansen method of Monte Carlo estimation of Sobol’ sensitivity indices for variance decomposition (*75, 76*) with 20 000 sample points and 1000 bootstrap replicates. Sobol’ indices were calculated for all loss functions ***f*** as well as for their weighted sum ***F*** and in all dimensions. Whilst keeping the number of sample points to as low as possible for computational reasons, we ascertained that first-order indices summed to 1 and total effects >1. We further ensured that the overall results of the Sobol’ analysis were consistent with the results of other global sensitivity analyses, namely the relative parameter importance derived from training a random forest (Supplementary Fig. S32).

### Synthetic data validation

Synthetic field data was generated by forward simulation using the final parameter sets from each optimization process. The two optimization algorithms were run anew using the respectively generated synthetic data to calculate the goodness of fit statistics. The parameter sets retrieved by the validation were compared against the parameterization yielded by the optimization process.

### Epidemiological outcome comparison

We conducted a small experiment to compare key epidemiological outcomes from the new parameterizations with the original model and that detail in a four malaria model comparison in Penny et al. 2016 (*12*). We simulated malaria in archetypical transmission and seasonality settings using the different parameterizations. The experiments were set up in a full-factorial fashion, considering the simulation options described in Table 1. Monitored outcomes were the incidence of uncomplicated, severe disease, hospitalizations, and indirect and direct malaria mortality over time and by age, prevalence over time and by age, the prevalence-incidence relationship, and the EIR-prevalence relationship. Simulations were conducted for a population of 10,000 individuals over 10 years.

**Table 1:** Full experimental design in setting archetypes. Experiments were run at 36% probability that an infected individual receives effective care within 14 days.

| Number of stochastic realization | Seasonality | Transmission (EIR) | Parameterization |
| --- | --- | --- | --- |
| 10 | Perennial | 0.25, 0.5, 0.75, 1, 1.1, 1.25, 1.35, | GA |
|  | Seasonal (sinusoidal) | 1.5, 1.75, 2, 2.5, 3, 4, 5, 6, 7, 8, 9, | GP-BO |
|  |  | 10, 12, 14, 16, 18, 20, 22, 25, 30, | GPSG-BO |
|  |  | 35, 40, 45, 50, 64, 73, 80, 100, |  |
|  |  | 128, 150, 200, 256, 512 |  |

### Software

Consistent with previous calibration work, we used OpenMalaria version 35, an open-source simulator written in C++ and further detailed in full in the supplement, OpenMalaria wiki (https://github.com/SwissTPH/openmalaria/wiki) or in the original publications (*3, 21, 24*). Calibration was performed using R 3.6.0. For the machine learning processes, all algorithms were accessed through the mlr package version 2.17.0(*66*). The heteroskedastic Gaussian process utilised the hetGP package under version 1.1.2(*65*). The sensitivity analysis was conducted using the soboljansen function of the sensitivity package version 1.21.0 in R (*77*). All algorithms were adapted to the operating system (CentOS 7.5.1804) and computational resources available at the University of Basel Center for Scientific Computing, SciCORE, which uses a Slurm queueing system. The full algorithm code is available on GitHub (https://github.com/reikth/BayesOpt_Calibration) and can be easily adapted to calibrate any simulation model. The number of input parameters and objective functions are flexible. Thus, to adapt the code to other simulators, code should be updated to run the respective model simulator, and tailored to user’s operating system. Further requirements to adapt the workflow are sufficient calibration data, and a per-objective goodness-of-fit metric.

## Data Availability

All code is available on git: https://github.com/reikth/BayesOpt_Calibration

## DATA AND CODE AVAILABILITY

Code is publicly available on GitHub under https://github.com/reikth/BayesOpt_Calibration and all calibration data was detailed in (*24*) and further available from the researchers on request.

## ACKNOWLEDGMENTS

We acknowledge and thank our colleagues in the Swiss TPH Disease Modelling unit. Calculations were performed at sciCORE (http://scicore.unibas.ch/) scientific computing core facility at University of Basel

## AUTHOR CONTRIBUTIONS

MAP and EC conceived the study. Algorithm development by EC, TR, MAP, and SF with implementation and preparation for sharing on GitHub by TR. Loss functions by MAP and TAS. Sensitivity analysis by TR with inputs from MG and LB. First draft was written by TR and MAP, all authors contributed to writing and interpretation of results and approved the final manuscript.

## SUPPLEMENTARY MATERIALS

Supplementary Texts 1 and 2

Supplementary Figures S1-S32

Supplementary Tables S1-S6

## Supplementary Information for

## 1 SUPPLEMENTARY TEXT 1: MALARIA TRANSMISSION MODEL

### 1.1 Main features

We test our calibration algorithm on OpenMalaria, an individual-based model of malaria dynamics. To provide context of the model’s structure and the role of the fitted parameters (see supplementary text 1), we here briefly describe its main features and key equations. This description is adapted from that provided in Smith et al. 2012 (*1*) and Smith et al. 2006 (*2*). Full details of all model components can be found in *The American Journal of Tropical Medicine and Hygiene*, Volume 75, Issue 2 Supplement (2006).

OpenMalaria features discrete individual-based stochastic simulations of malaria in humans in 5-day time steps. Every infection and individual are characterized by a set of continuous state variables, namely, parasite densities, infection durations, and immune status. Key processes and relationships regarding the course of infection simulated by model include the attenuation of inoculations, acquired pre-erythrocytic immunity, acquired blood-stage immunity, morbidity (acute and severe) and mortality (malaria-specific and indirect), anemia, and the infection of vectors as a function of parasite densities in the human. Other model components include a vector model and a case management system. All individual components have previously been well documented (*1, 2*). A visual summary of the model with references to further details on each component is provided in Fig. S1.

In our current recalibration only the original (base) model variant is used to test our new approach (*1*). Parameters estimated during the calibration process are highlighted and summarized in Table S1 at the end of this section. Other parameter values were drawn from the literature or were calibrated to separate data: for example, the empirical parasite density model of Maire et al. 2006 (*3*) was calibrated to malariatherapy (*4*) data and not recalibrated at the population level.

**Figure S1.**
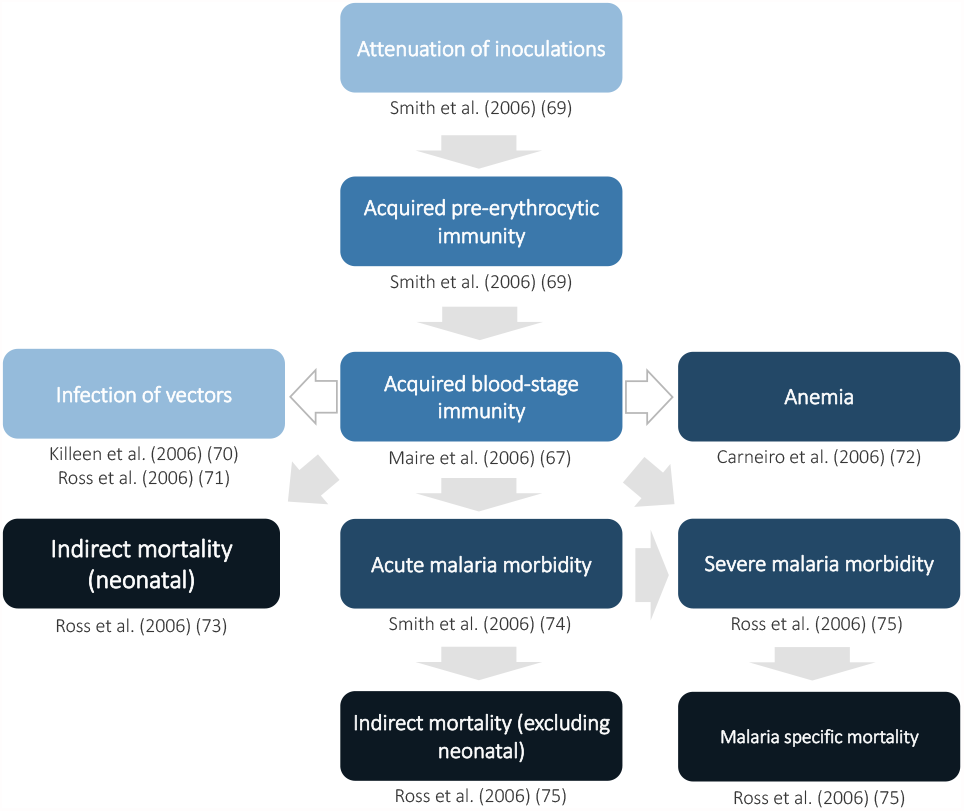
Visual summary of OpenMalaria with references to original publications on the model components. Adapted from Smith et al. 2006, Fig.3 (*2*).References from top to bottom and left to right (Attenuation of inoculations (*5*)),(Acquired pre-erythrocytic immunity (*5*)),(Infection of vectors [(*6*), (*7*)], Acquired blood-stage immunity (*3*), Anemia (*8*)),(Indirect mortality (neonatal) (*9*), Acute malaria morbidity (*10*), severe malaria morbidity (*11*)),(Indirect mortality excluding neonatal (*11*), Malaria specific mortality (*11*))

### 1.2 Infection of the human host

The seasonal pattern of entomological inoculation rate (EIR) determines seasonal pattern of transmission and thus the parasite densities in the individual, modified by natural or acquired immunity and interventions (*2*).

#### 1.2.1 Differential feeding by mosquitoes depending on body surface area

In the base model, the expected number of entomological inoculations experienced by individual *i* of age *a* at time *t* is

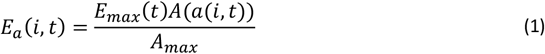

where *E*_*max*_(*t*) refers to the annual entomological inoculation rate (EIR) computed from human bait collections on adults and *A*(), is the individual’s availability to mosquitoes, assumed to be proportional to average body surface area, depending only on age *a*(*i, t*). *A*(*a*(*i,t*)) increases with age up to age 20 years where it reaches a value of *A*_*max*_ (the average body surface of people ≥ 20 years old in the same population).

The biting rate in relation to human weight is based on data from The Gambia published by Port and others (*12*), where the proportion of mosquitoes that had fed on a host were analyzed in relation to the host’s contribution to the total biomass and surface area of people sleeping in one mosquito net (*5*).

### 1.2.2 Control of pre-erythrocytic stages

The number of infective bites received per unit time for each individual *i*, adjusted by age, is given by Eq. 1 above. A survival function *S*(*i, t*) defines the probability that the progeny of an inoculation survives to give rise to a patent blood stage infection, i.e. the proportion of inoculations that result in infections or the susceptibility of individual *i* at time *t*. The force of infection is modelled as

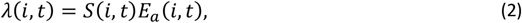

where *E*_*a*_(*i, t*) is the expected number of entomological inoculations endured by individual *i* at time *t*, adjusted for age and individual factors, and the number of infections *h*(*i, t*) acquired by individual ! in five-day time step *t*, follows a Poisson distribution:

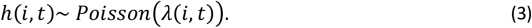

The susceptibility of individual *i* at time *t, S*(*i, t*)is defined as:

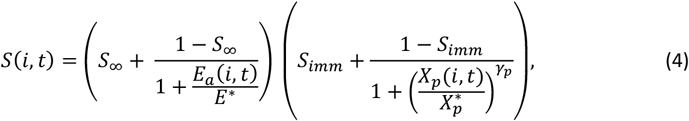

where 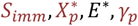and *S*_∞_ are constants representing the lower limit of success probability of inoculations in immune individuals, critical value of cumulative number of entomologic inoculations, critical value of *E*_*a*_(*i, t*), steepness of relationship between success of inoculation and *X*_*p*_(*i, t*), and, the lower limit of success probability of inoculations at high where *E*_*a*_(*i, t*), respectively. Here

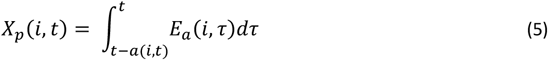

*S*_∞_ and *E*^∗^ are fixed to *S*_∞_ = 0.049, and *E*^∗^ = 0.032 inoculations/person-night and are detailed in (*5*).

#### 1.2.3 Course of infection in the human host

The model for each individual infection N in host ! comprises a time series of parasite densities. The base model for infection within humans is described in Maire et al. 2006 (*3*). In brief, the duration of each infection, *τ*_*max*_ is sampled from

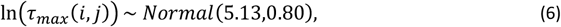

parameterised against malaria therapy data (*4*) and detailed in Maire et al. 2006 (*3*). In the absence of previous exposure or concurrent infections, the log density of infection *j* in host *i* at each time point, *τ* = 0,1, …, *τ*_*max*_ (*i*, N) is normally distributed with expectation

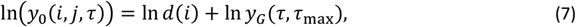

Where *y*_*G*_(*τ, τ*_*max*_) is taken from a statistical description of parasite densities in malariatherapy patients and *d*(*i*) describes between-host variation with a log-normal distribution with variance 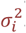.

We consider the possibility of multiple concurrent infections in the same individual at the same time. Exposure to asexual blood stages is measured by

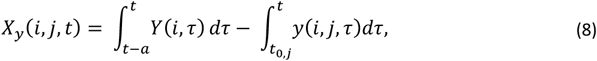

where *Y*(*i, τ*) is the total parasite density of individual *i* at time *τ* and *y*(*i, j, τ*) is the density of infection N in individual ! at time *τ* and

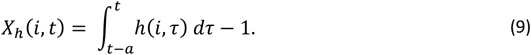

In the presence of previous exposure and co-infection, the expected log density for each concurrent infection is then:

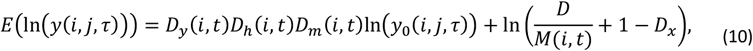

where *M*(*i, t*)is the total multiplicity of infection of in individual *i* at time *t*, and

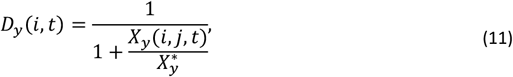

where 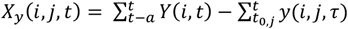 (note that a continuous time approximation to this is given in the original publications (*3, 5*) and hence measures the cumulative parasite load. Furthermore

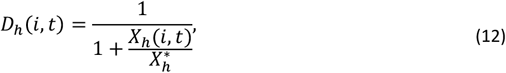

where,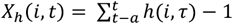, the number of inoculations since birth, excluding the one under consideration, which measures the diversity of inocula experienced by the host up to the time point under consideration.

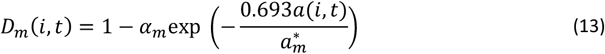

which measures the effect of maternal immunity. 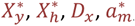, and *α*_*m*_ are all constants estimated in the fitting process. These constants are described in Table S1, or further in Maire et al. 2006 (*3*).

Variation within individuals described as 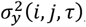, where

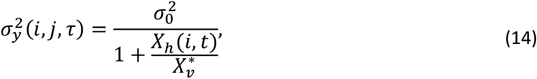

and 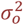 and 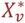are constants, described in Table S1.

The simulated density of infection *j* in individual *i* at time *τ* is then drawn from a normal distribution:

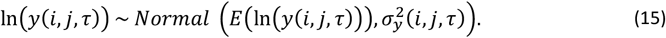

The total density of all infections in individual *i* at time t is then the sum of the densities of concurrent infections *j*

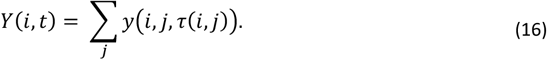

#### 1.2.4 Infectivity of the human host

The model infectivity of the human host is described in Ross 2006 where infectivity of individual I at time t is given by the distributed lag model:

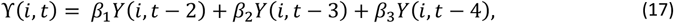

where *t* is in 5-day units and

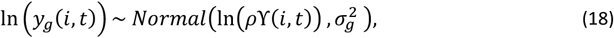

where 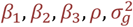 are constants representing contributions of past infections to gametocyte densities (detailed in Table S1), and to be calibrated at the population level. We define

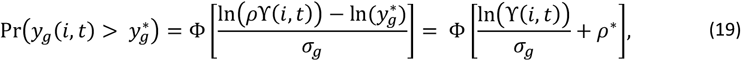

where Φ is the cumulative normal distribution, 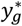 is the density of female gametocytes necessary for infection of the mosquito, and 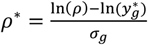 is constant (depending on the blood meal volume, gametocyte viability and system variability). Thus, the proportion of mosquitoes infected by individual *i* at time *t* is defined as

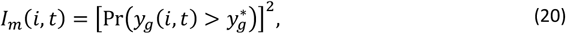

and the probability of a mosquito becoming infected during any feed is

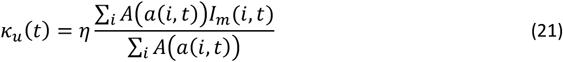

where *η* is a constant scale factor and to be calibrated.

We define 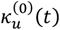 as the value of *k*_*u*_(*t*) in the simulation of an equilibrium scenario to which an intervention has been applied. Let 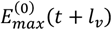 be the corresponding entomologic inoculation rate. 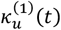 and 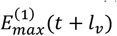 are the corresponding values for the intervention scenario. Then

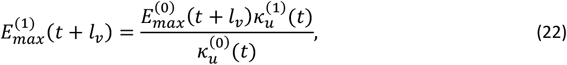

where *l*_*v*_ corresponds to the duration of the sporogenic cycle in the vector, which we approximate with two time steps (10 days). 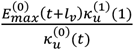 is the total vectorial capacity)

### 1.3 Morbidity

In order to simulate the clinical state of individual *i* at time *t*, for each five-day time step 5 independent samples from the simulated parasite density distribution are drawn for each concurrent infection *j*.

#### 1.3.1 Acute morbidity (uncomplicated clinical cases)

The model for an episode of acute morbidity was originally described in (*10*) and occurs in individual *I* at time *t* with probability

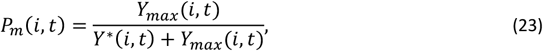

where *Y*^∗^is the pyrogenic threshold and *Y*_*max*_ is the maximum density of five daily densities sampled during the five-day interval *t*.

The pyrogenic threshold changes over time following

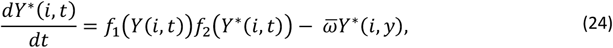

where *f*_1_(*Y*(*i, t*)) is a function describing the relationship between accrual of tolerance and the parasite density *Y*(*i, y*); *f*_2_(*Y*^∗^(*i, t*)) describes the saturation of this accrual process at high values of *Y*^∗^ and 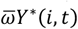 determines the decay threshold with first-order kinetics, ensuring that the parasite tolerance is short-lived (*10*).

Here *f*_1_(*Y* (*i, t*)) is defined to ensure that the stimulus is not directly proportional to *Y* but rather that it asymptotically reaches a maximum at high values of *Y*:

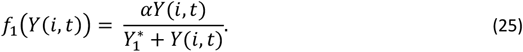

At high values of *Y*^∗^, a higher parasite load is required to achieve the same increase:

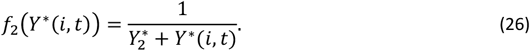

Thus, the pyrogenic threshold *Y*^∗^is defined to follow

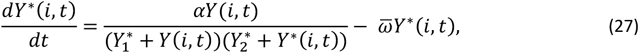

and the initial condition 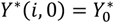 at the birth of the host, where 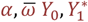 and 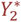 are targets of the calibration, and are defined in Table S1.

#### 1.3.2 Severe disease

The model for severe disease was described in Ross et al 2006 (*11*) and two different classes of severe episodes are considered by the model, *B*_1_and *B*_2_. *P*_*B*1_(*i, t*) is the probability that an acute episode (*A*) is of class *B*_1_ and

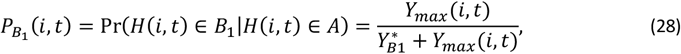

where 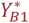 is a constant to be calibrated and *H*(*i, t*) is the clinical status of individual *i* at time *t*.

Class *B*_2_ of severe malaria episodes occurs when an otherwise uncomplicated episode coincides with some other insult, which occurs with risk

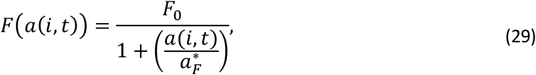

where *F*_0_ is the limiting value of *F*(*a*(*i, t*)) at birth and 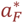 is the age at which it is halved and both are to be calibrated.

The probability that individual *i* experiences an episode belonging to class *B*_2_ at time *t*, conditional on there being a clinical episode at that time is

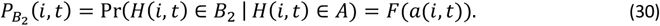

The age ant time specific risk of severe malaria morbidity conditional on a clinical episode is then given by

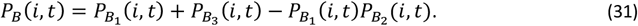

#### 1.3.3 Mortality

Malaria deaths in hospital are a random sample of admitted severe malaria cases, with age-dependent sampling fraction *Q*_*h*_(*a*), the hospital case fatality rate, derived from the data of Reyburn et al (2004) (*13*). The original model was described in Ross et al. 2006 (*11*).

The severe malaria case fatality in the community for age group *a, Q*_*c*_(*a*) is estimated as

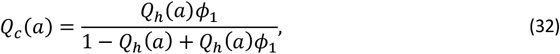

where ϕ_1_ the estimated odds ratio for death in the community compared to death in in-patients is an age-independent constant to be calibrated and *Q*_*h*_(*a*) is the hospital case fatality rate. The total malaria mortality is the sum of the hospital and community malaria deaths.

The risk of neonatal mortality attributable to malaria (death in class *D*_1_) in first pregnancies is set equal to 0.3 *μ*_*PG*_ where

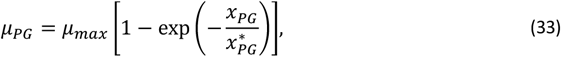

where *x*_*PG*_ is related to *x*_*MG*_, the prevalence in simulated individuals 20-24 ears of age via

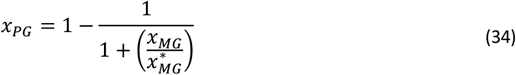

and 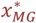 and 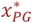 are constants to be calibrated and are detailed in Table S1.

An indirect death in class *D*_2_ is provoked at time *t*, conditional on there being a clinical episode at that time with probability *P*_*D*2_(*i, t*) where

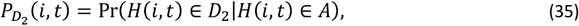

and

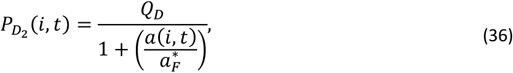

where *Q*_*D*_ is limiting value of 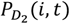 at birth and 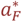 is a constant to be calibrated. Deaths in class *D*_2_ occur 30 days (six time steps) after the provoking episode.

**Table S1:** Names and details of OpenMalaria core parameters. GA-O = Genetic algorithm optimization, GP-BO = Gaussian process-based Bayesian optimization, GPSG-BO = Gaussian process stacked generalization-based Bayesian optimization

| No.* | $\theta^+$ | Parameter | Meaning | Unit/<br>dimension | Prior | GA-O estimate<br>(Smith et al. 2012, model<br>R0001)(1) | New estimate GP-BO<br>(Reiker et al.2020) | New estimate GPSG-BO<br>(Reiker et al.2020) |
| --- | --- | --- | --- | --- | --- | --- | --- | --- |
| 1 | -- | $-\ln(1 - S_\infty)$ | $S_\infty$ = Lower limit of success probability of inoculations at high $E_a(i, t)$ | Proportion | -- | 0.051 | 0.051 | 0.051 |
| 2 | -- | $E^*$ | Critical value of $E_a(i, t)$ | Inoculations/ person-night | -- | 0.032 | 0.032 | 0.032 |
| 3 | 1<sup>a</sup> | $S_{imm}$ | Lower limit of success probability of inoculations in immune individuals | Proportion | $\exp(N(\log(0.14), 2))$ | 0.138 | 0.196 | 0.036 |
| 4 | 3 | $X_p^*$ | Critical value of cumulative number of entomologic inoculations | Inoculations | $\exp(N(\log(1514), 2))$ | 1,514.4 | 1,954.8 | 4,972.2 |
| 5 | 2 | $\gamma_p$ | Steepness of relationship between success of inoculation and $X_p(i, t)$ | Dimensionless constant | $\exp(N(\log(1), 1))$ | 2.037 | 1.291 | 1.871 |
| 6 | 23 | $\sigma_i^2$ | Variation between hosts on parasite densities (variance of log-normal distribution) | | $\exp(N(\log(10.17), 0.6))$ | 10.174 | 11.729 | 9.689 |
| 7 | 5 | $X_y^*$ | Critical value of cumulative number of parasite days | Parasite-days/$\mu L \times 10^{-7}$ | $\exp(N(\log(3.52 \times 10^7), 2))$ | 3.516 | 593.661 | 1.216 |
| 8 | 4 | $X_h^*$ | Critical value of cumulative number of infections | Infections | $\exp(N(\log(97.3), 2))$ | 97.335 | 54.082 | 89.759 |
| 9 | 7 | $\ln(1 - \alpha_m)$ | $\alpha_m$ = Maternal protection at birth | Dimensionless | $-\log(1 - \text{Beta}(8, 2))$ | 2.330 | 1.770 | 1.266 |
| 10 | 8 | $\alpha_m^*$ | Decay of maternal protection | Per year | $\exp(N(\log(1.8), 0.5))$ | 2.531 | 1.279 | 1.551 |
| 11 | 9 | $\sigma_0^2$ | Fixed variance component for densities | $[\ln(\text{density})]^2$ | $\exp(N(\log(0.66), 2))$ | 0.656 | 5.838 | 1.440 |
| 12 | 6 | $X_v^*$ | Critical value of cumulative number of infections for variance in parasite densities | Infections | $\exp(N(\log(5), 1))$ | 0.916 | 3.959 | 7.226 |
| 13 | 14 | $Y_2^*$ | Critical value of $Y^*(i, t)$ in determining increase in $Y^*$ | Parasites/$\mu L$ | $\exp(N(\log(5000), 1))$ | 6,502.26 | 6,560.08 | 13,485.57 |
| 14 | 10 | $\alpha$ | Factor determining increase in $Y^*(i, t)$ | $\text{Parasites}^2 \mu L^{-2} \text{day}^{-1}$ | $\exp(N(\log(142602), 1))$ | 142,602 | 63,220.5 | 119,502 |
| 15 | 22 | $\nu_1$ | Density bias (non Garki) | Dimensionless | $\exp(N(\log(0.177), 0.6))$ | 0.177 | 0.123 | 0.159 |
| 16 | -- | $\sigma_2$ | Mass action parameter | Dimensionless | | 1 | 1 | 1 |
| 17 | 18 | $\log \phi_1$ | Case fatality for severe episodes in the community compared to hospital | Log odds | $\exp(N(\log(2.09), 0.3))$ | 0.736 | 0.340 | 0.285 |
| 18 | 20<sup>b</sup> | $Q_D$ | Co-morbidity intercept relevant to indirect mortality | Proportion | $\exp(N(\log(0.019), 1))$ | 0.019 | 0.019 | 0.023 |
| 19 | 19<sup>c</sup> | $Q_n$ | Non-malaria intercept for infant mortality | Deaths / 1000 live births | $\exp(N(\log(49.5), 1))$ | 49.539 | 46.5095 | 40.163 |
| 20 | 21 | $\nu_0$ | Density bias (Garki) | Dimensionless | $\exp(N(\log(4.79), 0.2))$ | 4.796 | 3.739 | 5.618 |
| 21 | 15 | $Y_{B_1}$ | Parasitaemia threshold for severe episodes type $B_1$ | Parasites/μL | $\exp(N(\log(250000), 0.8))$ | 784,456 | 849,046 | 484,122 |
| 22 | -- | -- | Immune penalty |  | -- | 1 | 1 | 1 |
| 23 | -- | -- | Immune effector decay |  | -- | 0 | 0 | 0 |
| 24 | 16<sup>d</sup> | $F_0$ | Prevalence of co-morbidity/susceptibility at birth relevant to severe episodes ($B_2$) | proportion | $\exp(N(\log(0.092), 0.5))$ | 0.097 | 0.078 | 0.094 |
| 25 | 11 | $\frac{\log 2}{\bar{\omega}}$ | $Y^*$(pyrogenic threshold) half-life | Years | $\frac{\log(2)}{\exp(N(\log(2.52), 1))}$ | 0.275 | 0.468 | 0.516 |
| 26 | 13 | $Y_1^*$ | Critical value of parasite density in determining increase in $Y^*$ | Parasites/μL | $\exp(N(\log(6), 2))$ | 0.597 | 1.665 | 0.477 |
| 27 | -- | -- | Asexual immunity decay |  | -- | 0 | 0 | 0 |
| 28 | 12 | $Y_0^*$ | Pyrogenic threshold at birth | Parasites/μL | $\exp(N(\log(296.3), 1))$ | 296.302 | 90.938 | 201.671 |
| 29 | -- | -- | Idete multiplier | Dimensionless | -- | 2.798 | 2.799 | 2.799 |
| 30 | 17 | $\alpha_F^*$ | Critical age for co-morbidity | Years | $\exp(N(\log(0.225), 0.8))$ | 0.117 | 0.138 | 0.087 |

\* Parameter number assigned for simulations in OpenMalaria scenarios, some parameters here are used in model variants and not in the base model. Listed for completeness; \*Parameter number $\theta_i$ assigned for the optimisation problem. $\theta$ is drawn from the unit cube and determines the quantiles of the prior for the parameter value. <sup>a</sup> quantile = $\theta * 0.8372102$ . <sup>b</sup> quantile = $\theta * 0.9999991$ . <sup>c</sup> quantile = $\theta * 0.9986755$ . <sup>d</sup> quantile = $\theta * 0.999963$ .

## 2 SUPPLEMENTARY TEXT 2: CALIBRATION APPROACH AND DATA SUMMARY

A comprehensive epidemiological calibration dataset was collated in order to parameterize OpenMalaria. This calibration dataset covers a total of eleven different epidemiological relationships (or objectives for fitting) that span important aspects of the natural history of malaria. Data were collated from different settings (see Table S2 for summary) and were detailed in the original model descriptions (*2, 10*) and a later parameterization (*1*). A total of 61 simulation scenarios were setup to parameterize OpenMalaria, constructed to simulate the study surveys and study sites that yielded the calibration dataset. The study site observations were replicated in OpenMalaria by reproducing the timing of the surveys and their endpoints (such as prevalence and incidence) and matching simulation options to the setting with regards to transmission intensity and seasonality, vector species, treatment seeking behavior and anti-malarial interventions. The objectives and data are further detailed below.

The parameter estimation process is a multi-objective optimization problem with each of the epidemiological quantities in Table S2 representing one objective. The aim of the optimization is to find a parameter set that maximizes the goodness of fit by minimizing a loss statistic computed as the weighted sum of the loss functions for each objective. Building a weighted average reduces the multiple loss terms to a single overall loss statistic, defined as:

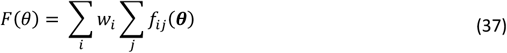

where *f*_*ij*_(***θ***) is the loss function for parameter vector ***θ***, epidemiological quantity *i* and dataset *j*, and the weights *w*_*i*_ were chosen so that different epidemiological quantities contribute approximately equally to *F*(***θ***).

For the current calibration, we utilised the loss functions from Smith et al. 2012 (*1*), the loss function *f*_*i*_(***θ***) for each objective *i* use either (negative) log-likelihoods or Residual Sum of Squares (RSS) with an unknown minimum. We did not update these loss-functions in order to compare to our previous approaches.

The likelihood functions are given by

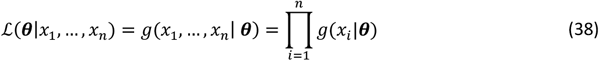

where the observed values are *x*_1_, …, *x*_*n*_ and the model parameters ***θ***. In practice, it is easier to work with the log likelihood, namely

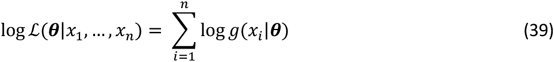

The loss functions *f*_*i*_(***θ***) used for each objective are detailed in the following sections.

### 2.1 Objectives: Epidemiological data and loss functions

Below we described each fitting objective in terms of the data (setting, surveys, observations, references) along with the associated loss function and original references. Table S3 provides an overview of the 61 simulation scenarios used for calibration, and which objective they contribute to.

#### 2.1.1 Age pattern of incidence after intervention

##### 2.1.1.1 Data

The data used for the calibration of objective 1 (Age pattern of incidence) consists of eight cross-sectional surveys of infection rates by age and EIR in Matsari village, capturing 12 age groups each. Matsari village was monitored entomologically for four years (Nov 1970 - Nov1973) during the Garki Project and multiple anti-malaria interventions were administered (*14*). From October 1970 to March 1972 (the baseline / pre-intervention phase), eight cross-sectional malariologic surveys of the whole village population and intensive entomologic surveillance (human bait collection of mosquitoes and dissections of the mosquito salivary glands for sporozoites) were carried out. The latter was used to estimate a baseline transmission intensity of 67 inoculations per person per year (EIR) and to derive seasonal transmission patterns. Mid-1972 marked the beginning of the intervention phase, during which an additional eight surveys were carried out at 10-week intervals (surveys 9-16). During this time, indoor residual spraying with Propoxur was carried out comprehensively in the village, along with mass treatment of the population with Sulfadoxine-pyrimethamine at 10 week-intervals immediately after assessment of individuals’ parasitologic status. The experimental setup is summarised in Fig. 3 of Smith et al 2006 (*5*). Incidence data (number of patent infections and number of hosts by age) from surveys 9-16 was used for our calibration.

###### Sites and scenario numbers

Matsari, Nigeria (30)

###### Original reference detailing data and model fits

*Smith TA, Maire N, Dietz K, Killeen GF, Vounatsou P et al. Relationship between the entomological inoculation rate and the force of infection for Plasmodium falciparum malaria. Am J Trop Med Hyg. Volume 75, No. 2 Supplement. 2006 (5)*

##### 2.1.1.2 Loss function: Binomial Log Likelihood

We denote the Binomial log likelihood for this objective to be

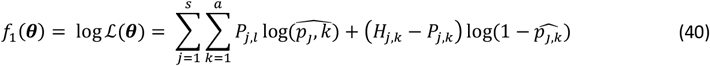

where *a* is the number of age groups,. the number of surveys, *p*_*j,k*_ the scenario data number of parasite positive hosts and *H*_*j,k*_ the scenario data number of hosts for age group *k* and survey *j*. Parameter 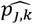 is associated with the model predictions and is given by

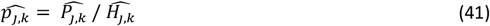

where 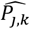 are the predicted number of parasite positive hosts and 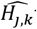 the predicted number of hosts for age group *k* and survey *j*.

#### 2.1.2 Age patterns of prevalence

##### 2.1.2.1 Data

The data used for the calibration of objective 2 (age-patterns of prevalence) consists of six cross-sectional malariology surveys conducted in the Rafin Marke, Matsari, Sugungum villages in Nigeria 1970-1972 (12 age groups each, part of the Garki Project during the pre-intervention period) (*14*), Navrongo in Ghana 2000 (12 age groups) (*15*) and Namawala 1990-1991 (*16*) and Idete in Tanzania (11 and 6 age groups, respectively) 1992-1993 (*17*). In all study sites, annual transmission intensity (EIR) and seasonal patterns were assessed using light trap or human night bait collections and dissections of the salivary glands (see Fig. 2 in Maire et al. 2006 (*3*)). In all sites except Idete, the health system at the time of the surveys treated only a small proportion of the clinical malaria episodes. In the Idete, the village dispensary was assumed to treat approximately 64% of clinical malaria (based on the published literature). During simulation, prevalence was defined by comparing each predicted parasite density with the limit of detection used in the actual study.

###### Sites and scenario numbers

Sugungum, Nigeria (24); Rafin-Marke, Nigeria (28); Matsari, Nigeria (29); Idete, Tanzania (31); Navrongo, Ghana (34); Namawala, Tanzania (35)

###### Original reference detailing data and model fits

*Maire N, Smith TA, Ross A, Owusu-Agyei S, Dietz K, et al. A model for natural immunity to asexual blood stages of Plasmodium falciparum malaria in endemic areas. Am J Trop Med Hyg. Volume 75, No. 2 Supplement. 2006 (3)*

##### 2.1.2.2 Loss function: Binomial Log Likelihood

We denote the binomial log likelihood for each scenario of this objective to be

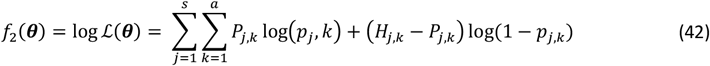

where *a* is the number of age groups, *s* the number of surveys, *p*_*j,k*_ the scenario data number of parasite positive hosts and *H*_*j,k*_ the scenario data number of hosts for age group k and survey j. Parameter *p*_*j,k*_ is associated with the model predictions and is given by

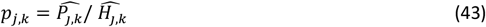

where 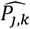 are the predicted number of parasite positive hosts and 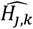 the predicted number of hosts for age group *k* and survey *j*.

#### 2.1.3 Age patterns of parasite density

##### 2.1.3.1 Data

The same data sources as for objective 2 (age pattern of prevalence) were used for calibration of objective 3 (age pattern of parasite density). Parasite densities in sites that were part of the Garki project (Sugungum, Rafin-Make and Matsari, Nigeria) were recorded by scanning a predetermined number of microscope fields on the thick blood film and recording how many had one or more asexual parasites visible. These were converted to numbers of parasites visible by assuming Poisson distribution for the number of parasites per field and a blood volume of 0.5 mm^3^ per 200 fields. In the other studies (Idete and Namawala, Tanzania and Navrongo, Ghana), parasites were counted against leukocytes and converted to nominal parasites/microliter assuming the usual standard of 8,000 leukocytes/microliter. The biases in density estimates resulting from these different techniques were accounted for by multiplying the observed parasite densities with constant values estimated for Garki (*v*_0_) and non-Garki (*v*_1_) studies to rescale them to the values in malariatherapy patients (*18*).

###### Sites and scenario numbers

Sugungum, Nigeria (pre-intervention, 24); Rafin-Marke, Nigeria (pre-intervention, 28); Matsari, Nigeria (pre-intervention, 29); Idete, Tanzania (31); Navrongo, Ghana (34); Namawala, Tanzania (35)

###### Original reference detailing data and model fits

*Maire N, Smith TA, Ross A, Owusu-Agyei S, Dietz K, et al. A model for natural immunity to asexual blood stages of Plasmodium falciparum malaria in endemic areas. Am J Trop Med Hyg. Volume 75, No. 2 Supplement. 2006* (*3*)

##### 2.1.3.2 Loss function: Log-normal log likelihood

For objective 3 (age pattern of parasite densities) we denote the log-Normal log likelihood for each scenario to be

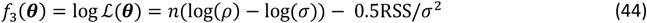

where *n* is the number of observations in the data set, *ρ* = exp(−0.5 log(2 *π*)), a constant from the log-normal likelihood, RSS is the residual sum of squares given by

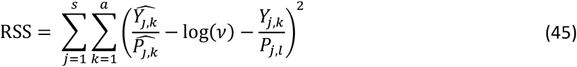

and *σ* is the standard deviation given by

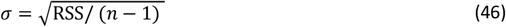

Here, *v* is the appropriate density bias, which is a fitting parameter, *a* is the number of age groups, *s* is the number of surveys, *P*_*j,k*_ the scenario number of parasite positive hosts, and *Y*_*j,k*_ the sum of the log densities, 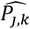 the predicted number of parasite positive hosts and 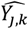 the predicted sum of the log densities for age group *k* and survey *j*. The density bias are fitting parameters *v*_0_ and *v*_1_.

#### 2.1.4 Age pattern of number of concurrent infections

##### 2.1.4.1 Data

For objective 4 (age pattern of number of concurrent infections), the dataset from Navrongo, Ghana (also used in the calibration of objectives 2 and 3) was used to calibrate to the total numbers of distinct parasite infections in one individual in each age group, and at each survey. Distinct infections were detected by polymerase chain reaction-restriction fragment length polymorphism in the sampled individuals.

###### Sites and scenario numbers

Navrongo, Ghana (34)

###### Original reference detailing data and model fits

*Maire N, Smith TA, Ross A, Owusu-Agyei S, Dietz K, et al. A model for natural immunity to asexual blood stages of Plasmodium falciparum malaria in endemic areas. Am J Trop Med Hyg. Volume 75, No. 2 Supplement. 2006 (3)*

##### 2.1.4.2 Loss function: Poisson Log Likelihood

Assuming that both the data and the simulations are Poisson distributed about the correct value and thereby also allowing for over-dispersion, we denote the Poisson log likelihood for each scenario to be for the objective of age pattern of number of concurrent infections to be

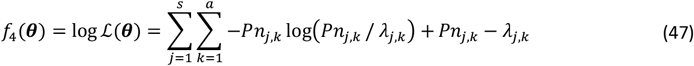

where *a* is the number of age groups, *s* the number of surveys, *Pn*_*j,k*_ the scenario data total patent infections for age group *k* and survey *j*. Parameter *λ* _*j,k*_ is associated with the model predictions and is given by

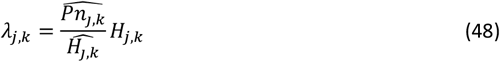

where 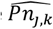 are the predicted total of patent infections and 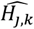 the predicted number of hosts for age group *k* and survey *j* and *H*_*j,k*_ is the scenario data number of hosts for age group *k* and survey *j*.

#### 2.1.5 Age pattern of incidence of clinical malaria

##### 2.1.5.1 Data

Two distinct datasets representing three study sites (Table S4) were used for the calibration of objective 5 and objective 6 (age pattern of incidence of clinical malaria). For Objective 5, the dataset contains data on the age pattern of clinical episodes in the villages of Ndiop and Dielmo in Senegal (*19, 20*). During the study period of July 1990 - June 1992, the village populations were visited daily to detect and treat any clinical malaria attacks with quinine. Cases were detected by reporting of symptoms (fever) during daily active case detection and subsequent thick blood smear microscopy. Only symptomatic individuals (axillary temperature ≥ 38.0°C or rectal temperature ≥ 38.5°C). Due to the active case detection and rapid treatment all symptomatic episodes are assumed to be effectively treated in these villages during the study period. No effective treatment of clinical malaria was assumed prior to the study period. The annual patterns of transmission were replicated as reported by Charlwood et al (1998) (*21*). A proportion *P*_*t*_ =35.75% are assumed to be treated effectively in Idete. As all individuals reporting to the village dispensary were treated presumptively with chloroquine, this proportion corresponds to the proportion of episodes reported to the village dispensary.

###### Sites and scenario numbers

Ndiop, Senegal (232), Dielmo, Senegal (233)

###### Original reference detailing data and model fits

*Smith TA, Ross A, Maire N, Rogier C, Trape J-F et al. An epidemiologic model of the incidence of acute illness in Plasmodium falciparum malaria. Am J Trop Med Hyg. Volume 75, No. 2 Supplement. 2006 (10)*

##### 2.1.5.2 Loss function: RSS-biased

We denote a loss function based on biased residual sum of squares:

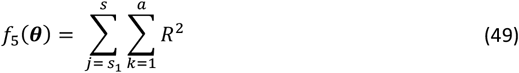

where *a* is the number of age groups, *s* the number of surveys, *s*_1_ the initial survey number, and *R* is the residual given by

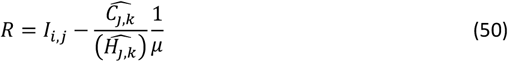

where *I*_*j,k*_ is the observed recorded incidence rate, 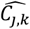 are the predicted total cases (severe and uncomplicated), 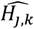 the predicted number of hosts for age group *k* and survey *j* and *μ* is a bias related to the scenario. For scenarios 232 and 233 (representing Ndiop and Dielmo, Senegal) this bias is *μ* = 5 indicating the duration in years for which episodes are collected. For scenario 49 in Objective 6 (Idete, Tanzania) the bias is *μ* = 0.357459 and represents the proportion of episodes reported to the village dispensary.

#### 2.1.6 Age pattern of incidence of clinical malaria: infants

##### 2.1.6.1 Data

Objective 6 (age pattern of incidence of clinical malaria in infants) is informed by a dataset on incidence that contains passive case detection data on the age-incidence in infants recorded at the health centre in Idete, Tanzania from June 1993-October 1994 (*17*). The annual patterns of transmission were replicated as reported by Charlwood et al (1998) (*21*).

###### Sites and scenario numbers

Idete, Tanzania (49))

###### Original reference

*Smith TA, Ross A, Maire N, Rogier C, Trape J-F et al. An epidemiologic model of the incidence of acute illness in Plasmodium falciparum malaria. Am J Trop Med Hyg. Volume 75, No. 2 Supplement. 2006 (10)*

##### 2.1.6.2 Loss function: RSS-biased

The loss function for Objective 6 is the same as Objective 5. For scenario 49 (Idete, Tanzania) the bias is *μ* = 0.357459 and represents the proportion of episodes reported to the village dispensary.

#### 2.1.7 Age pattern of threshold parasite density for clinical attacks

##### 2.1.7.1 Data

Objective 7 (Age pattern of threshold parasite density for clinical attacks), uses the dataset from Dielmo, Senegal (see objective 5) for calibration. The pyrogenic threshold in the (OpenMalaria) predictions is output as the sum of the log threshold values across age groups. The pyrogenic threshold per age group is given as the parasite:leucocyte ratio for recorded incidence of disease. To adjust these densities to the same scale as that used in fitting the simulation model to other datasets, the parasite:leukocyte ratios were multiplied by a factor of 1,416 to give a notional density in parasites/microliter of blood. This number was derived as follows: Parasites were counted against leukocytes and converted to nominal parasites/microliter assuming the usual (though biased) standard of 8,000 leukocytes/microliter. The biases in density estimates resulting from these different techniques was accounted for by multiplying the observed parasite densities with constant values estimated for Garki (*v*_0_) and non-Garki (*v*_1_) studies to rescale them to the values in malariatherapy patients (*18*).The value 1416 comes from

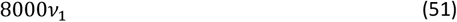

where the original *v*_1_ ≈ 0.18.

###### Sites and scenario numbers

Dielmo, Senegal (234)

###### Original reference detailing data and model fits

*Smith TA, Ross A, Maire N, Rogier C, Trape J-F et al. An epidemiologic model of the incidence of acute illness in Plasmodium falciparum malaria. Am J Trop Med Hyg. Volume 75, No. 2 Supplement. 2006 (10)*

##### 2.1.7.2 Loss function: RSS-biased (log)

For the objective 7 (Age pattern of threshold parasite density for clinical attacks) we denote a residual sum of squares loss function given by (13) with

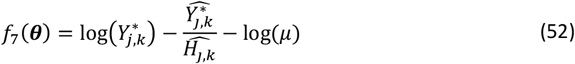

where *Y*^∗^ is the observed pyrogenic threshold, 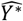 are the predicted sum log pyrogenic threshold, 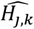 the predicted number of hosts for age group *k* and survey *j* and is a bias related to the scenario. Here, this bias is related to the log parasite/leucocyte ratio and thus *μ* = 1/(8000*v*_1_) where *v*_1_ is the non-Garki density bias.

#### 2.1.8 Hospitalization rate in relation to prevalence in children

##### 2.1.8.1 Data

Data on the relative incidence of severe malaria-related morbidity and mortality in children <9 years old across different transmission intensities were originally collated by Marsh and Snow (1999) (*22*) (Table 4). Data measurements per age group were available as the relative risk of severe disease compared to age group 1 and the proportion/prevalence of severe episodes. A total of 26 entries on the relationship between severe malaria hospital admission rates and *P. falciparum* prevalence were used to calibrate objective 8 (Hospitalisation rate in relation to prevalence in children), each represented in a separate simulation scenario, with one observation per scenario. These are summarised in Table S5. To obtain a continuous function relating hospital incidence rates to prevalence, linear interpolation between data points was performed. To convert hospital incidence rates to community severe malaria incidence, the hospital admission rates was divided by the assumed proportion of severe episodes representing to hospital (48%). There was assumed to be no effective treatment of uncomplicated malaria episodes or malaria mortality.

###### Sites and scenario numbers

Bo, Sierra Leone (501); Niakhar, Senegal (502), Farafenni, The Gambia (503); Areas I-V, The Gambia (504-508); Gihanga, Burundi (509); Katumba, Burundi (510); Karangasso, Burkina Faso (511); Kilifi North, Kenya (512); Manhica, Mozambique (514); Namawala, Tanzania (515); Navrongo, Ghana (516); Saradidi, Kenya (517); Yombo, Tanzania (518); Ziniare, Burkina Faso (519); Matsari, Nigeria (520); ITC control, Burkina Faso (521); Mlomp, Senegal (522); Ganvie, Benin (523); Kilifi Town, Kenya (524); Chonyi, Kenya (525); Bandafassi, Senegal (526); Kongodjan, Burkina Faso (527)

###### Original reference detailing data and model fits

*Ross A, Maire N, Molineaux L and Smith TA. An epidemiologic model of severe morbidity and mortality caused by Plasmodium falciparum. Am J Trop Med Hyg. Volume 75, No. 2 Supplement. 2006 (11)*

##### 2.1.8.2 Loss function: squared deviation

The loss function is denoted as the log of residual sum of squares

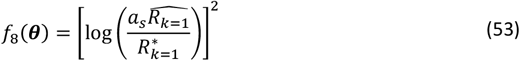

where *a*_*s*_ is the access to treatment of severe cases (0.48, estimated in base model), 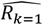 is the scenario predicted rate of severe episodes per 1000 person-years for age group *k* = 1 (0-9 years), and parameter 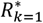 is the interpolated observed rate of severe episodes per 1000 person year given by

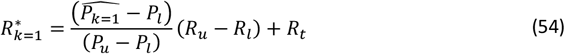

where 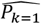 is the predicted prevalence summed over all surveys, *P*_*u*_ and *P*_*l*_ are the observed prevalences above and below the predicted prevalence 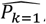, respectively and *R*_*u*_ and *R*_*l*_ are the corresponding severe episode rates to the observed prevalences.

The predicted prevalence is given by

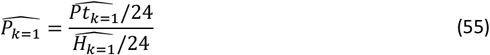

where 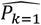 is the total number of parasite positive predicted and 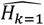 are the total number of hosts (division by 24 to give mean values). The predicted rate of episodes per 1000 person year is given by

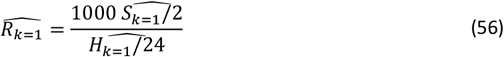

where 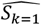 is the number of severe cases predicted and with division by 2 to convert to from 2 years to 1 year and the division by 24 to give mean number of hosts.

#### 2.1.9 Age pattern of hospitalization: severe malaria

##### 2.1.9.1 Data

For objective 9 (Age pattern of hospitalisation), a subset of the data collated by Marsh and Snow (1999) (*22*) (see objective 8) is used. Detailed age-specific severe hospital admission rates were available for 5 of the sites (Table S6). The patterns of incidence by age were summarised by age in 1-4 and 5-9 year-old children and compared with 1-11 month old infants by calculating the relative risk.

Of the five sites, four were selected for fitting objective 9 based on the predicted prevalence. Baku, The Gambia was excluded as the very low (2%) prevalence here could not be matched.

###### Sites and scenario number(s)

Area V, The Gambia (158); Saradidi, Kenya (167); Ganvie, Benin (173); Bandafassi, Senegal (176)

###### Original reference detailing data and model fits

*Ross A, Maire N, Molineaux L and Smith TA. An epidemiologic model of severe morbidity and mortality caused by Plasmodium falciparum. Am J Trop Med Hyg. Volume 75, No. 2 Supplement. 2006 (11)*

##### 2.1.9.2 Loss function: Residual sums of squares for relative risk

We denote a loss function based on residual sum of squares:

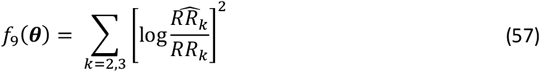

where *RR*_*k*_ is the relative risk of severe episode for age group *k* compared to age group 1 and 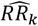 is the predictive relative risk for age group k compared to age group 1. The predicted relative risk is given by

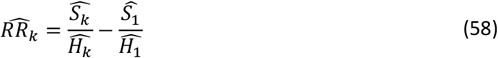

where 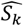 is the number of severe cases predicted for age group *k* and 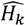 the total number of hosts for age group *k*.

#### 2.1.10 Malaria specific mortality in children (< 5 years old)

##### 2.1.10.1 Data

For objective 10 (Malaria specific mortality in children (<5 years old)), a subset of the data collated by Marsh and Snow (1999) (*22*) (see objective 8) was used (*24*). Mortality data were derived from verbal autopsy studies in sites with prospective demographic surveillance and were adjusted for the effect of malaria transmission intensity on the sensitivity and specificity of the cause of death determination. The odds ratio for death of a case in the community relative to that in hospital was estimated by fitting to the malaria-specific mortality rates in children less than five years of age assuming the published hospital case fatality rate. Nine sites for which both malaria-specific mortality rates and seasonal transmission patterns were available were included for calibration.

There is one observation per study site and simulation scenario, and predicted values are for one survey at the end of 2 years.

###### Sites and scenario number(s)

Bo, Sierra Leone (301); Niakhar, Senegal (302); Farafenni, The Gambia (303); Kilifi North, Kenya (312); Navrongo, Ghana (316); Saradidi, Kenya (317); Yombo, Tanzania (318); Bandafassi, Senegal (326); Kongodjan, Burkina Faso (327)

###### Original reference detailing data and model fits

*Ross A, Maire N, Molineaux L and Smith TA. An epidemiologic model of severe morbidity and mortality caused by Plasmodium falciparum. Am J Trop Med Hyg. Volume 75, No. 2 Supplement. 2006* (*11*)

##### 2.1.10.2 Loss function: Residual sums of squares

For objective 10 on Malaria specific mortality in children, the loss function minimizes the log sum of squares

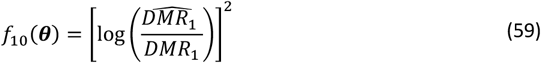

where *DMR*_1_ is the observed direct mortality rate for age group 1 (0-5 years) and 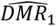 is the predicted direct mortality rate for age group 1. The predicted direct mortality rate is given by

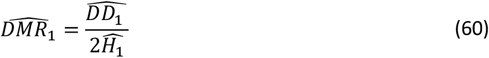

where 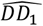 is the number of direct malaria deaths cases predicted for age group 1 and 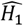 the total number of predicted hosts for age group 1. The division by 2 is to convert to yearly rate as the survey was conducted at the end of 2 years.

#### 2.1.11 Indirect malaria infant mortality rate

##### 2.1.11.1 Data

For objective 11 (indirect malaria infant mortality rate), a subset of the data collated by Marsh and Snow (1999) (*22*) (see objective 8) was used. These constitute a library of sites for which entomologic data were collected at least monthly and all-cause infant mortality rates (IMR) were available. There is one observation per scenario: all cause infant mortality rate (returned as a single number over whole intervention period).

###### Sites and scenario number(s)

Bo, Sierra Leone (401); Niakhar, Senegal (402); Area V, The Gambia (408); Karangasso, Burkina Faso (411); Manhica, Mozambique (414); Namawala, Tanzania (415); Navrongo, Ghana (416); Saradidi, Kanya (417); Yombo, Tanzania (418); Mlomp, Senegal (422); Bandafassi, Senegal (426)

###### Original reference detailing data and model fits

*Ross A, Maire N, Molineaux L and Smith TA. An epidemiologic model of severe morbidity and mortality caused by Plasmodium falciparum. Am J Trop Med Hyg. Volume 75, No. 2 Supplement. 2006 (11)*

##### 2.1.11.2 Loss function: Residual sums of squares

The loss function minimises the log sum of squares:

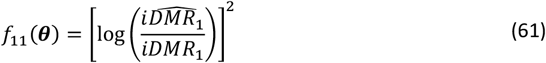

where *iDMR*_1_ the observed indirect mortality rate for age group 1 and 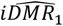 is the predicted indirect mortality rate for age group 1.

### 2.2 Tables S3-S4

**Table S2:** Epidemiological quantities and data sources used for parameterizing models. (a) Some scenarios are used to predict several outcomes, so the total of this column does not equal the total of 61 scenarios involved in fitting the models. (b) The number of data points is the sum over all scenarios and simulated survey periods of the number of age groups into which the data were disaggregated for comparison with the model predictions. (c) In relation to the EIR specified as a seasonal pattern. (d) Model predictions for this objective are compared with linear interpolations between the field data points. *The number of data points is the sum over all scenarios and simulated survey periods of the number of age groups into which the data were disaggregated for comparison with the model predictions. Table adapted from Table S1 in Smith et al 2012 **(*1*)**.

| Epidemiological quantity | Data sources | No. of scenarios | No. of data points* | Publication for fitting of base model | Prior | Weighting in GOF statistic | Scenario numbers | Loss vector number ($f_i$) | Loss function |
| --- | --- | --- | --- | --- | --- | --- | --- | --- | --- |
| Age pattern of incidence of infection after intervention | Molineaux and Gramiccia (1980) (14) | 1 | 12 | Maire et al 2006 (3) | Binomial | 0.001 | 30 | 1 | Binomial log-likelihood |
| Age patterns of prevalence of infection | Molineaux and Gramiccia (1980) (14) | 6 | 563 | Maire et al 2006 (3) | Binomial | 0.001 | 24, 28, 29, 35, 34, 31 | 2 | Binomial log-likelihood |
| Age patterns of parasite density | Molineaux and Gramiccia (1980) (14) | 6 | 563 | Maire et al 2006 (3) | Log Normal | 0.01 | 24, 28, 29, 35, 34, 31 | 3 | log likelihood |
| Age pattern of number of concurrent infections | Maire et al. 2006 (3); Owusu-Agyei et al 2002 (15) | 1 | 12 | Maire et al 2006 (3) | Poisson | 0.01 | 34 | 4 | Poisson log-likelihood |
| Age pattern of incidence of clinical malaria: age-specific Ndiop & Dielmo, Senegal | Trape and Rogier 1996 (19); Kitua et al 1996 (17) | 2 | 26 | Smith et al 2006 (5) | Log Normal | 1 | 232, 233, 49 | 5 | RSS |
| Age pattern of incidence of clinical malaria: infants Idete, Tanzania | Kitua et al 1996 (17) | 1 | 4 | Smith et al 2006 (5) | Log Normal | 1 | 49 | 6 | RSS |
| Age pattern of threshold parasite density for clinical attacks | Rogier et al 1996 (25) | 1 | 13 | Smith et al 2006 (5) | Log Normal | 1 | 234 | 7 | RSS |
| Hospitalisation rate in relation to prevalence in children | See Ross et al 2006 (11) | 26 | 10 | Ross et al 2006 (11) | Log Normal | 2 | 501, 502, 503, 504, 505, 506, 507, 508, 509, 510, 511, 512, 514, 515, 516, 517, 518, 519, 520, 521, 522, 523, 524, 525, 526, 527 | 8 | Squared deviation |
| Age pattern of hospitalisation: severe malaria | Marsh and Snow 1999 (22) | 4 | 12 | Ross et al 2006 (11) | Log Normal | 2 | 158, 167, 173, 176 | 9 | RSS |
| Malaria specific mortality in children (<5y) | Snow et al 1997 (23) | 9 | 9 | Ross et al 2006 (11) | Log Normal | 1 | 301, 302, 303, 312, 316, 317, 318, 326, 327 | 10 | Squared deviation logRate |
| All-cause infant mortality rate | See Ross et al 2006 (11) | 11 | 11 | Ross et al 2006 (11) | Log Normal | 10 | 401, 402, 408, 411, 414, 415, 416, 417, 418, 422, 426 | 11 | Squared deviation logRate |

**Table S3:**
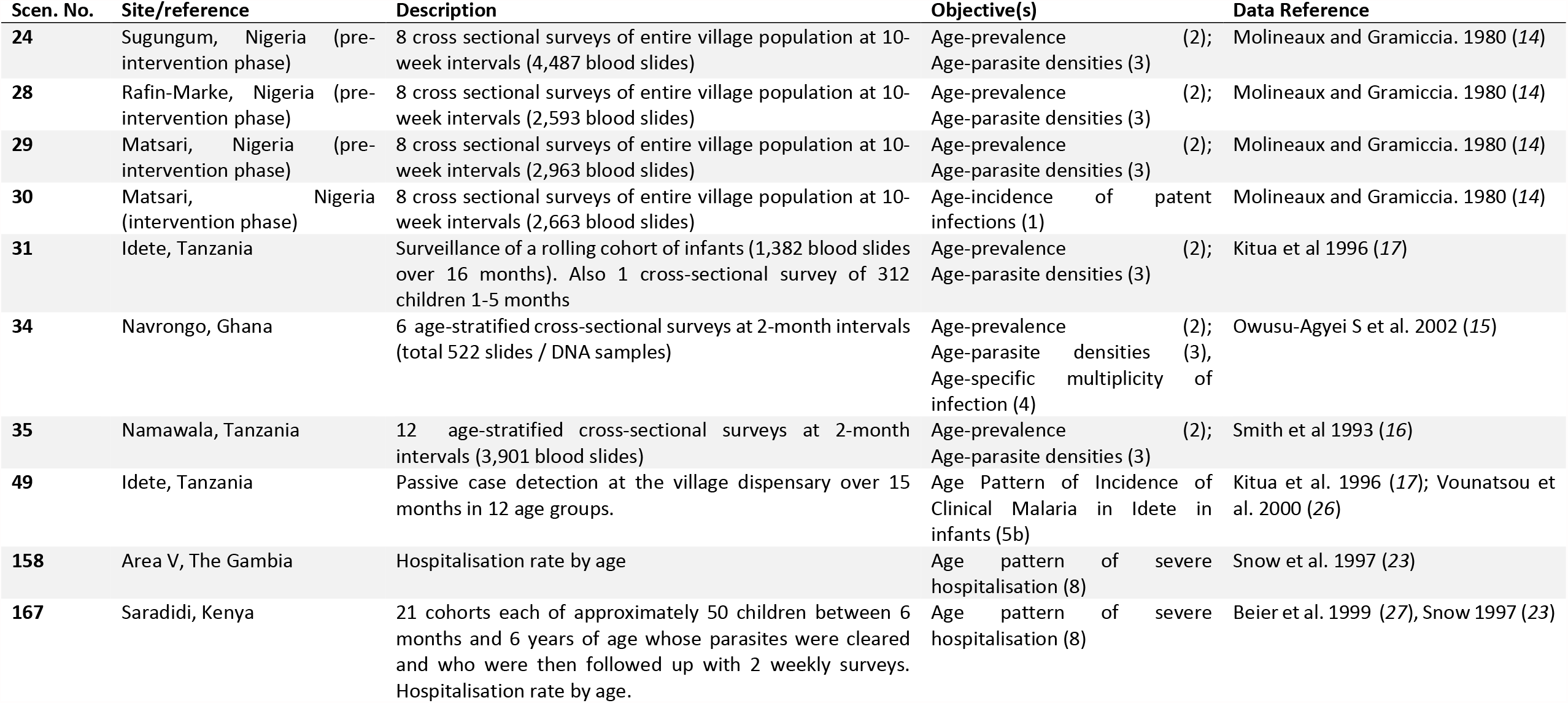
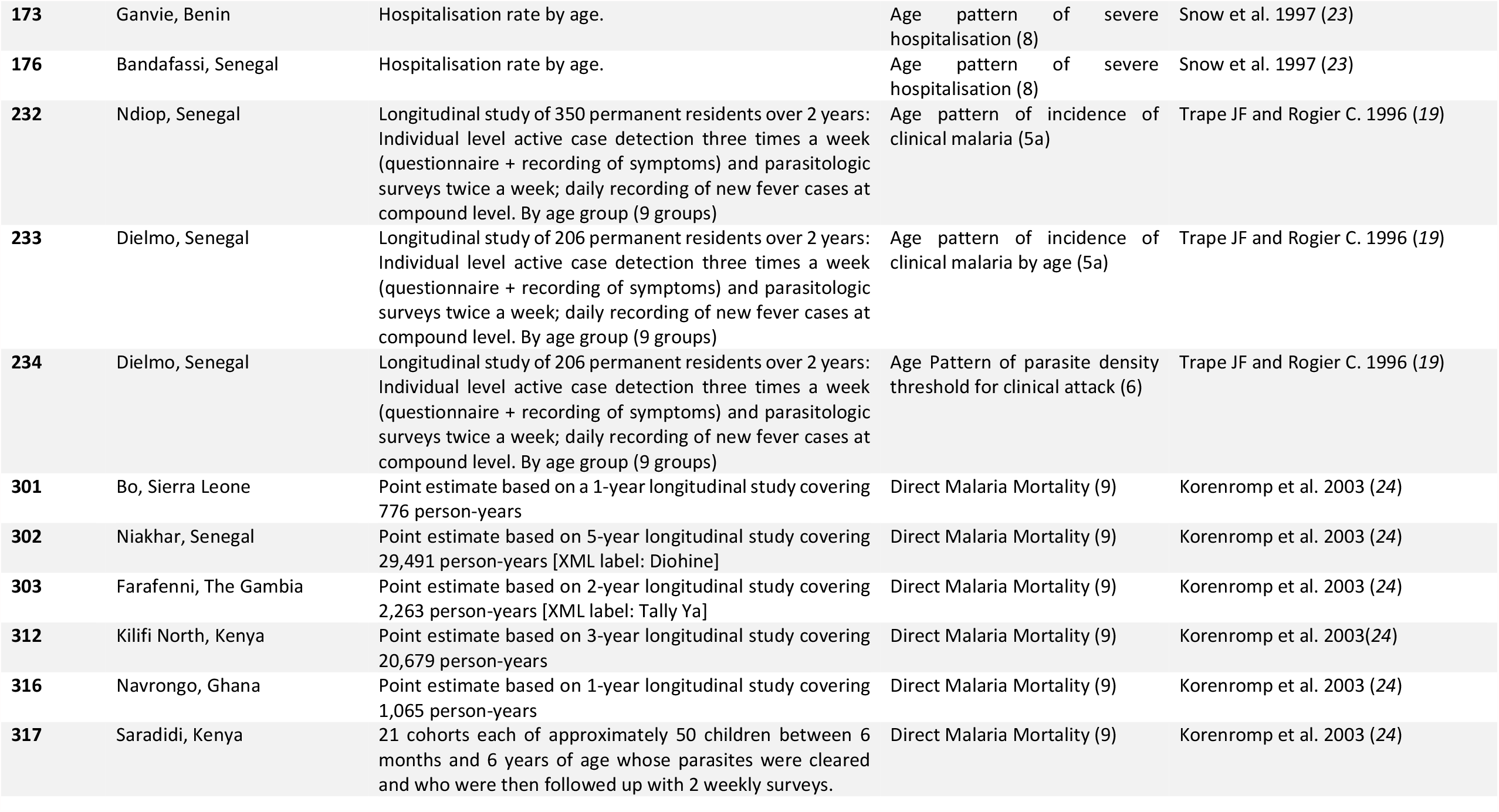
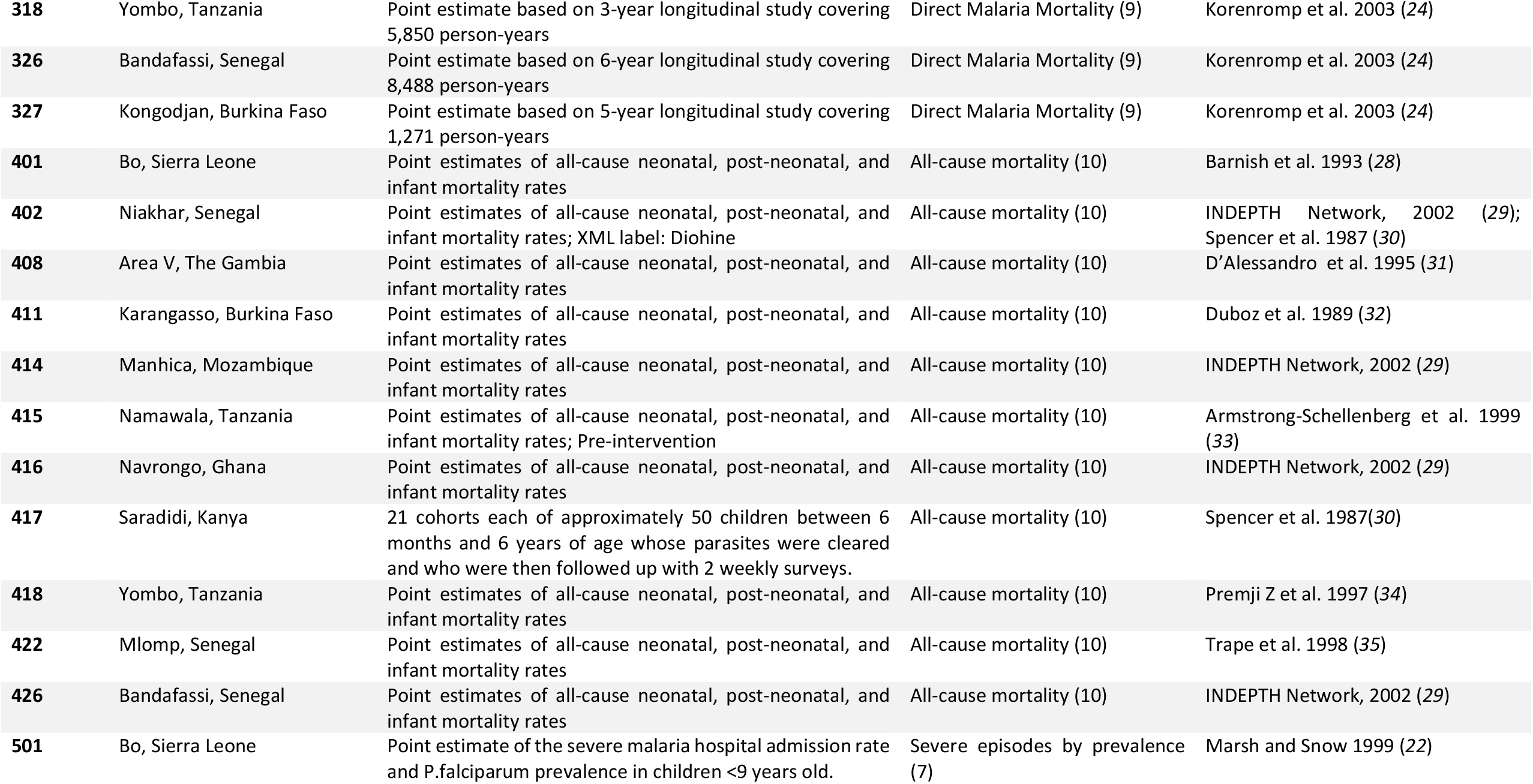
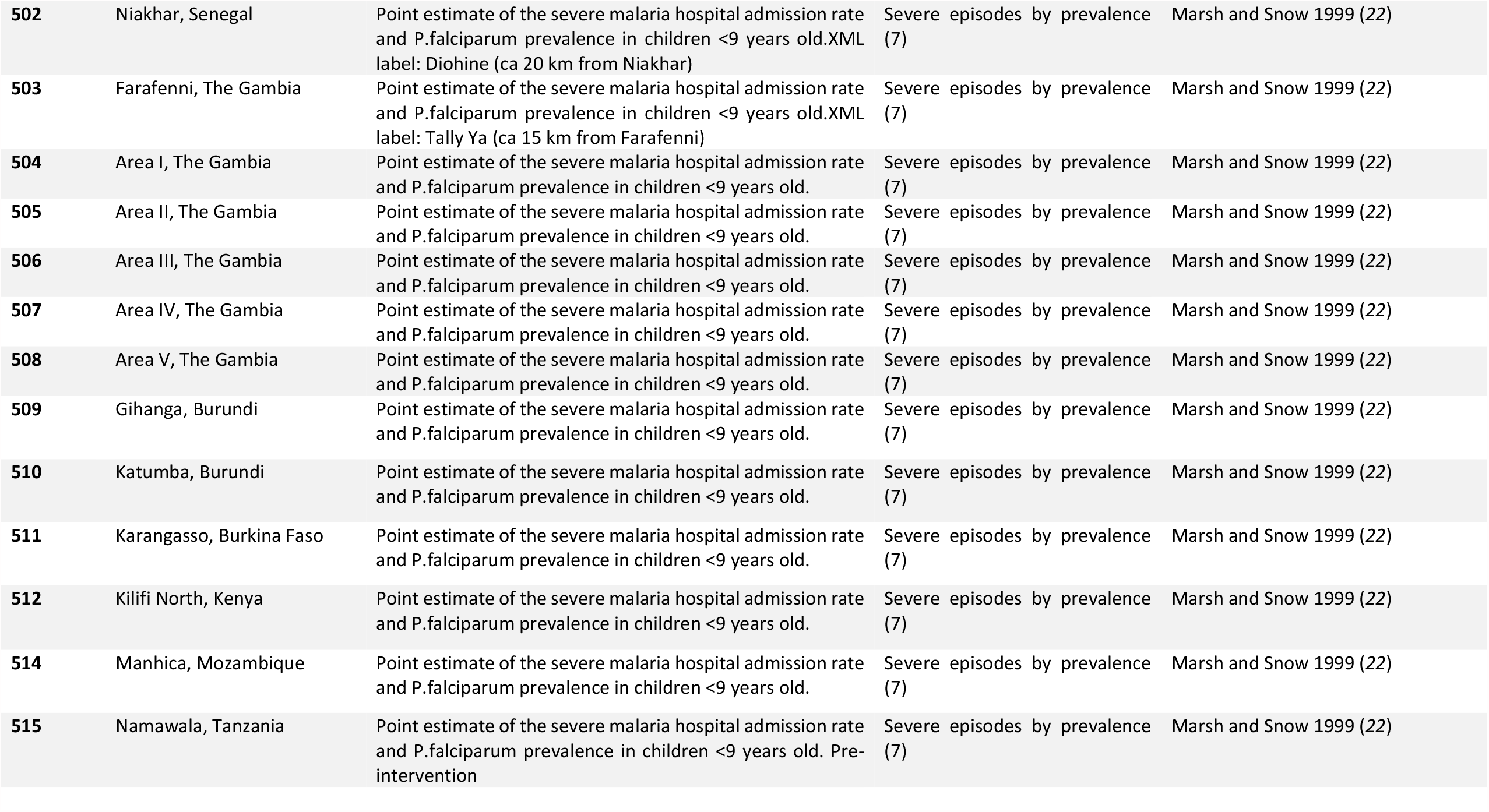
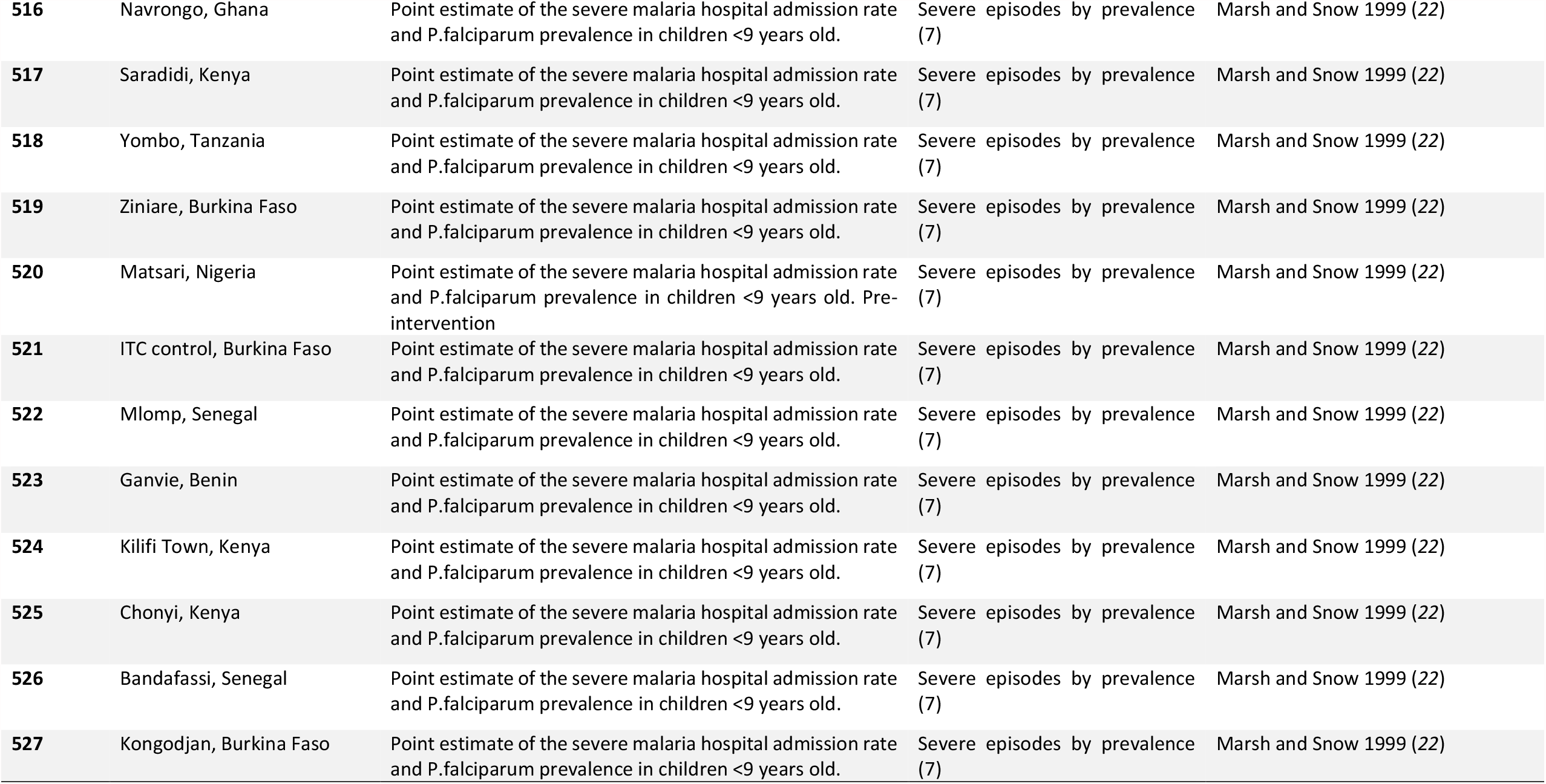
Calibration data for objectives 2-4, age patterns of prevalence, parasite densities, and multiplicity of infection.

| Scen. No. | Site/reference | Description | Objective(s) | Data Reference |
| --- | --- | --- | --- | --- |
| 24 | Sugungum, Nigeria (pre-intervention phase) | 8 cross sectional surveys of entire village population at 10-week intervals (4,487 blood slides) | Age-prevalence (2);<br>Age-parasite densities (3) | Molineaux and Gramiccia. 1980 (14) |
| 28 | Rafin-Marke, Nigeria (pre-intervention phase) | 8 cross sectional surveys of entire village population at 10-week intervals (2,593 blood slides) | Age-prevalence (2);<br>Age-parasite densities (3) | Molineaux and Gramiccia. 1980 (14) |
| 29 | Matsari, Nigeria (pre-intervention phase) | 8 cross sectional surveys of entire village population at 10-week intervals (2,963 blood slides) | Age-prevalence (2);<br>Age-parasite densities (3) | Molineaux and Gramiccia. 1980 (14) |
| 30 | Matsari, Nigeria (intervention phase) | 8 cross sectional surveys of entire village population at 10-week intervals (2,663 blood slides) | Age-incidence of patent infections (1) | Molineaux and Gramiccia. 1980 (14) |
| 31 | Idete, Tanzania | Surveillance of a rolling cohort of infants (1,382 blood slides over 16 months). Also 1 cross-sectional survey of 312 children 1-5 months | Age-prevalence (2);<br>Age-parasite densities (3) | Kitua et al 1996 (17) |
| 34 | Navrongo, Ghana | 6 age-stratified cross-sectional surveys at 2-month intervals (total 522 slides / DNA samples) | Age-prevalence (2);<br>Age-parasite densities (3),<br>Age-specific multiplicity of infection (4) | Owusu-Agyei S et al. 2002 (15) |
| 35 | Namawala, Tanzania | 12 age-stratified cross-sectional surveys at 2-month intervals (3,901 blood slides) | Age-prevalence (2);<br>Age-parasite densities (3) | Smith et al 1993 (16) |
| 49 | Idete, Tanzania | Passive case detection at the village dispensary over 15 months in 12 age groups. | Age Pattern of Incidence of Clinical Malaria in Idete in infants (5b) | Kitua et al. 1996 (17); Vounatsou et al. 2000 (26) |
| 158 | Area V, The Gambia | Hospitalisation rate by age | Age pattern of severe hospitalisation (8) | Snow et al. 1997 (23) |
| 167 | Saradidi, Kenya | 21 cohorts each of approximately 50 children between 6 months and 6 years of age whose parasites were cleared and who were then followed up with 2 weekly surveys. Hospitalisation rate by age. | Age pattern of severe hospitalisation (8) | Beier et al. 1999 (27), Snow 1997 (23) |
| **173** | Ganvie, Benin | Hospitalisation rate by age. | Age pattern of severe hospitalisation (8) | Snow et al. 1997 (23) |
| **176** | Bandafassi, Senegal | Hospitalisation rate by age. | Age pattern of severe hospitalisation (8) | Snow et al. 1997 (23) |
| **232** | Ndiop, Senegal | Longitudinal study of 350 permanent residents over 2 years: Individual level active case detection three times a week (questionnaire + recording of symptoms) and parasitologic surveys twice a week; daily recording of new fever cases at compound level. By age group (9 groups) | Age pattern of incidence of clinical malaria (5a) | Trape JF and Rogier C. 1996 (19) |
| **233** | Dielmo, Senegal | Longitudinal study of 206 permanent residents over 2 years: Individual level active case detection three times a week (questionnaire + recording of symptoms) and parasitologic surveys twice a week; daily recording of new fever cases at compound level. By age group (9 groups) | Age pattern of incidence of clinical malaria by age (5a) | Trape JF and Rogier C. 1996 (19) |
| **234** | Dielmo, Senegal | Longitudinal study of 206 permanent residents over 2 years: Individual level active case detection three times a week (questionnaire + recording of symptoms) and parasitologic surveys twice a week; daily recording of new fever cases at compound level. By age group (9 groups) | Age Pattern of parasite density threshold for clinical attack (6) | Trape JF and Rogier C. 1996 (19) |
| **301** | Bo, Sierra Leone | Point estimate based on a 1-year longitudinal study covering 776 person-years | Direct Malaria Mortality (9) | Korenromp et al. 2003 (24) |
| **302** | Niakhar, Senegal | Point estimate based on 5-year longitudinal study covering 29,491 person-years [XML label: Diohine] | Direct Malaria Mortality (9) | Korenromp et al. 2003 (24) |
| **303** | Farafenni, The Gambia | Point estimate based on 2-year longitudinal study covering 2,263 person-years [XML label: Tally Ya] | Direct Malaria Mortality (9) | Korenromp et al. 2003 (24) |
| **312** | Kilifi North, Kenya | Point estimate based on 3-year longitudinal study covering 20,679 person-years | Direct Malaria Mortality (9) | Korenromp et al. 2003(24) |
| **316** | Navrongo, Ghana | Point estimate based on 1-year longitudinal study covering 1,065 person-years | Direct Malaria Mortality (9) | Korenromp et al. 2003 (24) |
| **317** | Saradidi, Kenya | 21 cohorts each of approximately 50 children between 6 months and 6 years of age whose parasites were cleared and who were then followed up with 2 weekly surveys. | Direct Malaria Mortality (9) | Korenromp et al. 2003 (24) |
| **318** | Yombo, Tanzania | Point estimate based on 3-year longitudinal study covering 5,850 person-years | Direct Malaria Mortality (9) | Korenromp et al. 2003 (24) |
| **326** | Bandafassi, Senegal | Point estimate based on 6-year longitudinal study covering 8,488 person-years | Direct Malaria Mortality (9) | Korenromp et al. 2003 (24) |
| **327** | Kongodjan, Burkina Faso | Point estimate based on 5-year longitudinal study covering 1,271 person-years | Direct Malaria Mortality (9) | Korenromp et al. 2003 (24) |
| **401** | Bo, Sierra Leone | Point estimates of all-cause neonatal, post-neonatal, and infant mortality rates | All-cause mortality (10) | Barnish et al. 1993 (28) |
| **402** | Niakhar, Senegal | Point estimates of all-cause neonatal, post-neonatal, and infant mortality rates; XML label: Dihine | All-cause mortality (10) | INDEPTH Network, 2002 (29);<br>Spencer et al. 1987 (30) |
| **408** | Area V, The Gambia | Point estimates of all-cause neonatal, post-neonatal, and infant mortality rates | All-cause mortality (10) | D'Alessandro et al. 1995 (31) |
| **411** | Karangasso, Burkina Faso | Point estimates of all-cause neonatal, post-neonatal, and infant mortality rates | All-cause mortality (10) | Duboz et al. 1989 (32) |
| **414** | Manhica, Mozambique | Point estimates of all-cause neonatal, post-neonatal, and infant mortality rates | All-cause mortality (10) | INDEPTH Network, 2002 (29) |
| **415** | Namawala, Tanzania | Point estimates of all-cause neonatal, post-neonatal, and infant mortality rates; Pre-intervention | All-cause mortality (10) | Armstrong-Schellenberg et al. 1999 (33) |
| **416** | Navrongo, Ghana | Point estimates of all-cause neonatal, post-neonatal, and infant mortality rates | All-cause mortality (10) | INDEPTH Network, 2002 (29) |
| **417** | Saradidi, Kenya | 21 cohorts each of approximately 50 children between 6 months and 6 years of age whose parasites were cleared and who were then followed up with 2 weekly surveys. | All-cause mortality (10) | Spencer et al. 1987(30) |
| **418** | Yombo, Tanzania | Point estimates of all-cause neonatal, post-neonatal, and infant mortality rates | All-cause mortality (10) | Premji Z et al. 1997 (34) |
| **422** | Mlomp, Senegal | Point estimates of all-cause neonatal, post-neonatal, and infant mortality rates | All-cause mortality (10) | Trape et al. 1998 (35) |
| **426** | Bandafassi, Senegal | Point estimates of all-cause neonatal, post-neonatal, and infant mortality rates | All-cause mortality (10) | INDEPTH Network, 2002 (29) |
| **501** | Bo, Sierra Leone | Point estimate of the severe malaria hospital admission rate and P.falciparum prevalence in children <9 years old. | Severe episodes by prevalence (7) | Marsh and Snow 1999 (22) |
| **502** | Niakhar, Senegal | Point estimate of the severe malaria hospital admission rate and P.falciparum prevalence in children <9 years old.XML label: Diohine (ca 20 km from Niakhar) | Severe episodes by prevalence (7) | Marsh and Snow 1999 (22) |
| **503** | Farafenni, The Gambia | Point estimate of the severe malaria hospital admission rate and P.falciparum prevalence in children <9 years old.XML label: Tally Ya (ca 15 km from Farafenni) | Severe episodes by prevalence (7) | Marsh and Snow 1999 (22) |
| **504** | Area I, The Gambia | Point estimate of the severe malaria hospital admission rate and P.falciparum prevalence in children <9 years old. | Severe episodes by prevalence (7) | Marsh and Snow 1999 (22) |
| **505** | Area II, The Gambia | Point estimate of the severe malaria hospital admission rate and P.falciparum prevalence in children <9 years old. | Severe episodes by prevalence (7) | Marsh and Snow 1999 (22) |
| **506** | Area III, The Gambia | Point estimate of the severe malaria hospital admission rate and P.falciparum prevalence in children <9 years old. | Severe episodes by prevalence (7) | Marsh and Snow 1999 (22) |
| **507** | Area IV, The Gambia | Point estimate of the severe malaria hospital admission rate and P.falciparum prevalence in children <9 years old. | Severe episodes by prevalence (7) | Marsh and Snow 1999 (22) |
| **508** | Area V, The Gambia | Point estimate of the severe malaria hospital admission rate and P.falciparum prevalence in children <9 years old. | Severe episodes by prevalence (7) | Marsh and Snow 1999 (22) |
| **509** | Gihanga, Burundi | Point estimate of the severe malaria hospital admission rate and P.falciparum prevalence in children <9 years old. | Severe episodes by prevalence (7) | Marsh and Snow 1999 (22) |
| **510** | Katumba, Burundi | Point estimate of the severe malaria hospital admission rate and P.falciparum prevalence in children <9 years old. | Severe episodes by prevalence (7) | Marsh and Snow 1999 (22) |
| **511** | Karangasso, Burkina Faso | Point estimate of the severe malaria hospital admission rate and P.falciparum prevalence in children <9 years old. | Severe episodes by prevalence (7) | Marsh and Snow 1999 (22) |
| **512** | Kilifi North, Kenya | Point estimate of the severe malaria hospital admission rate and P.falciparum prevalence in children <9 years old. | Severe episodes by prevalence (7) | Marsh and Snow 1999 (22) |
| **514** | Manhica, Mozambique | Point estimate of the severe malaria hospital admission rate and P.falciparum prevalence in children <9 years old. | Severe episodes by prevalence (7) | Marsh and Snow 1999 (22) |
| **515** | Namawala, Tanzania | Point estimate of the severe malaria hospital admission rate and P.falciparum prevalence in children <9 years old. Pre-intervention | Severe episodes by prevalence (7) | Marsh and Snow 1999 (22) |
| 516 | Navrongo, Ghana | Point estimate of the severe malaria hospital admission rate and P.falciparum prevalence in children <9 years old. | Severe episodes by prevalence (7) | Marsh and Snow 1999 (22) |
| 517 | Saradidi, Kenya | Point estimate of the severe malaria hospital admission rate and P.falciparum prevalence in children <9 years old. | Severe episodes by prevalence (7) | Marsh and Snow 1999 (22) |
| 518 | Yombo, Tanzania | Point estimate of the severe malaria hospital admission rate and P.falciparum prevalence in children <9 years old. | Severe episodes by prevalence (7) | Marsh and Snow 1999 (22) |
| 519 | Ziniare, Burkina Faso | Point estimate of the severe malaria hospital admission rate and P.falciparum prevalence in children <9 years old. | Severe episodes by prevalence (7) | Marsh and Snow 1999 (22) |
| 520 | Matsari, Nigeria | Point estimate of the severe malaria hospital admission rate and P.falciparum prevalence in children <9 years old. Pre-intervention | Severe episodes by prevalence (7) | Marsh and Snow 1999 (22) |
| 521 | ITC control, Burkina Faso | Point estimate of the severe malaria hospital admission rate and P.falciparum prevalence in children <9 years old. | Severe episodes by prevalence (7) | Marsh and Snow 1999 (22) |
| 522 | Mlomp, Senegal | Point estimate of the severe malaria hospital admission rate and P.falciparum prevalence in children <9 years old. | Severe episodes by prevalence (7) | Marsh and Snow 1999 (22) |
| 523 | Ganvie, Benin | Point estimate of the severe malaria hospital admission rate and P.falciparum prevalence in children <9 years old. | Severe episodes by prevalence (7) | Marsh and Snow 1999 (22) |
| 524 | Kilifi Town, Kenya | Point estimate of the severe malaria hospital admission rate and P.falciparum prevalence in children <9 years old. | Severe episodes by prevalence (7) | Marsh and Snow 1999 (22) |
| 525 | Chonyi, Kenya | Point estimate of the severe malaria hospital admission rate and P.falciparum prevalence in children <9 years old. | Severe episodes by prevalence (7) | Marsh and Snow 1999 (22) |
| 526 | Bandafassi, Senegal | Point estimate of the severe malaria hospital admission rate and P.falciparum prevalence in children <9 years old. | Severe episodes by prevalence (7) | Marsh and Snow 1999 (22) |
| 527 | Kongodjan, Burkina Faso | Point estimate of the severe malaria hospital admission rate and P.falciparum prevalence in children <9 years old. | Severe episodes by prevalence (7) | Marsh and Snow 1999 (22) |

**Table S4:** summary of study data set for objective 5: Age pattern of incidence of clinical malaria.

| Scenario No. | Study site | Age groups | Observations |
| --- | --- | --- | --- |
| 232 | Ndiop, Senegal | 22 | One per age group |
| 233 | Dielmo, Senegal | 22 | One per age group |
| 49 | Idete, Tanzania | 4 | One per age group |

**Table S5.** Settings used for calibrating the incidence of severe malaria. (Adapte from Table1 from Ross et al. 2006 (*11*))

| Site | EIR data |  |
| --- | --- | --- |
|  | Year | EIR |
| **Burkina Faso** |  |  |
| ITC Control | 1994-1995 | 389 |
| Karangasso | 1985 | 244 |
| Kongodjan | 1984 | 133 |
| Ziniare | 1994-1995 | 70 |
| **Burundi** |  |  |
| Gihanga | 1983 | 205 |
| Katumba | 1982 | 13.6 |
| **Kenya** |  |  |
| Chonyi | 1992-1993 | 50 |
| Kilifi North | 1992-1003 | 10.5 |
| Kilifi Town | 1990-1991 | 2.8 |
| Saradidi | 1986-1987 | 239 |
| **Senegal** |  |  |
| Bandafassi | 1995-1996 | 363 |
| MIomp | 1995 | 30 |
| Niakhar | 1995 | 11.6 |
| **Tanzania** |  |  |
| Namawala | 1990-1991 | 329 |
| Yombo | 1992 | 234 |
| **The Gambia** |  |  |
| Area I-V | 1991 | + |
| Farafenni | 1987 | 8.9 |
| **Others** |  |  |
| Bo, Sierra Leone | 1990-1991 | 34.7 |
| Ganvie, Benin | 1993-1995 | 11 |
| Manhica, Mozambique | 2001-2002 | 38 |
| Matsari, Nigeria | 1971 | 68 |
| Navrongo, Ghana | 2001-2002 | 418 |

\*EIR = entomological inoculation rate, ITC = control group of randomised trial of insecticide-treated curtains. \*Five sites with annual EIR between 1 and 10

**Table S6:**
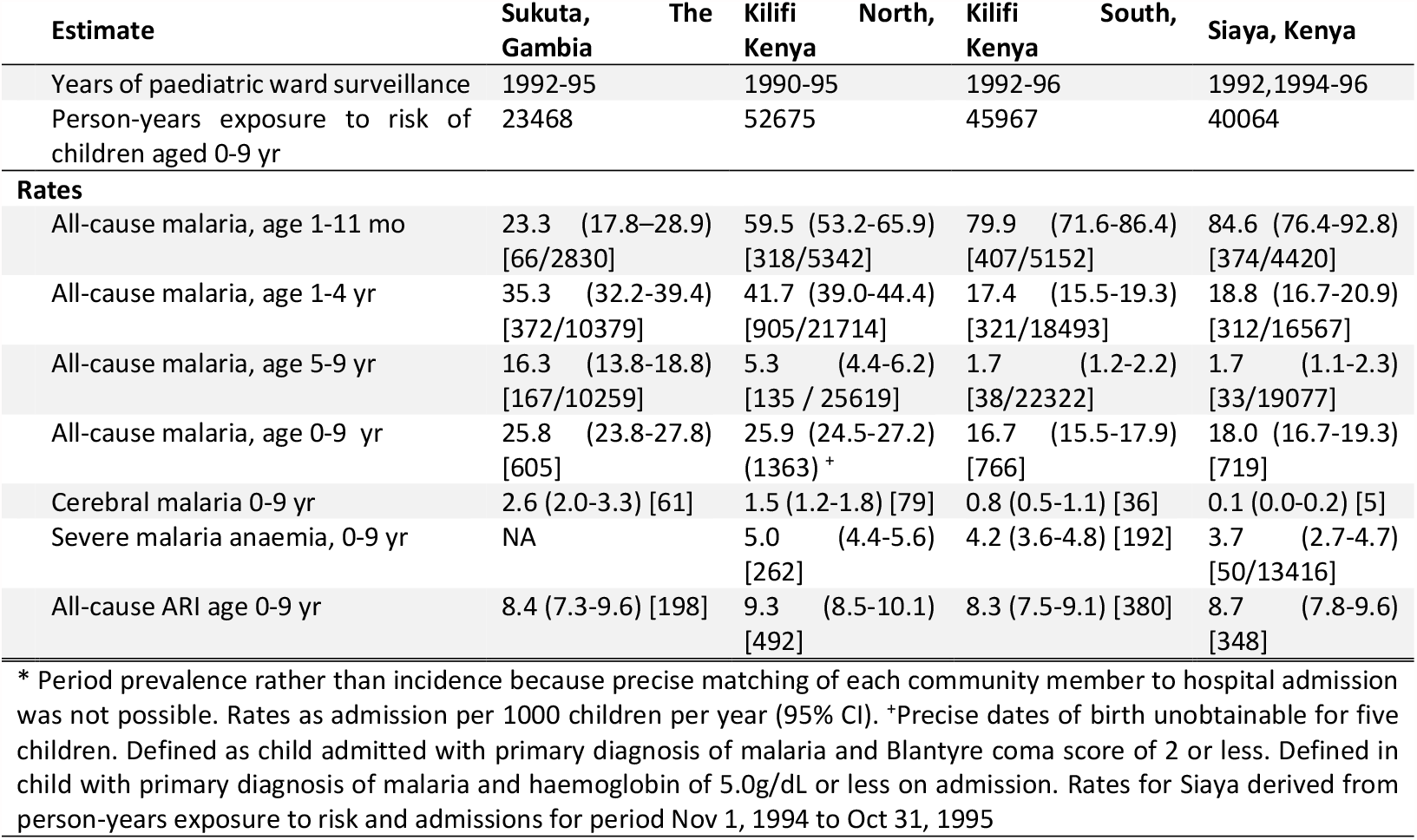
Age-specific period prevalence rates* of severe malaria, severe malaria, severe malaria anaemia and acute respiratory-tract infections from five communities in The Gambia and Kenya. (Adapted from Table 2 from Snow et al 1997 (*23*))

## 3 EMULATOR PERFORMANCE

**Figure S2.**
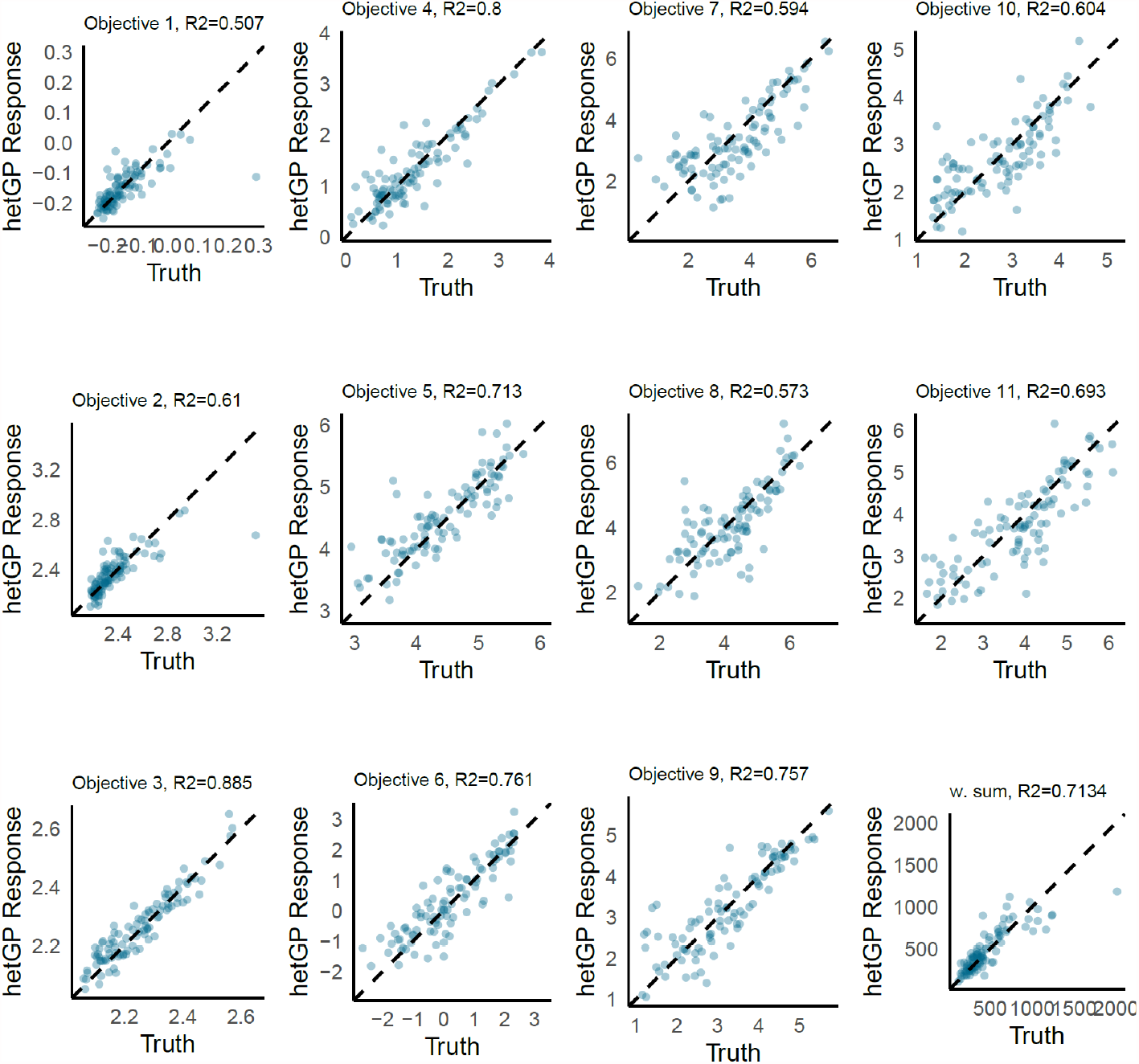
GP emulator performance. Emulator predictions vs true values on a holdout set compromising 10% of initial samples in iteration 1. w.sum is the weighted sum *F*, of the 11 objectives. Here, predictions are generated as the weighted sum of individual objective predictions.

**Figure S3.**
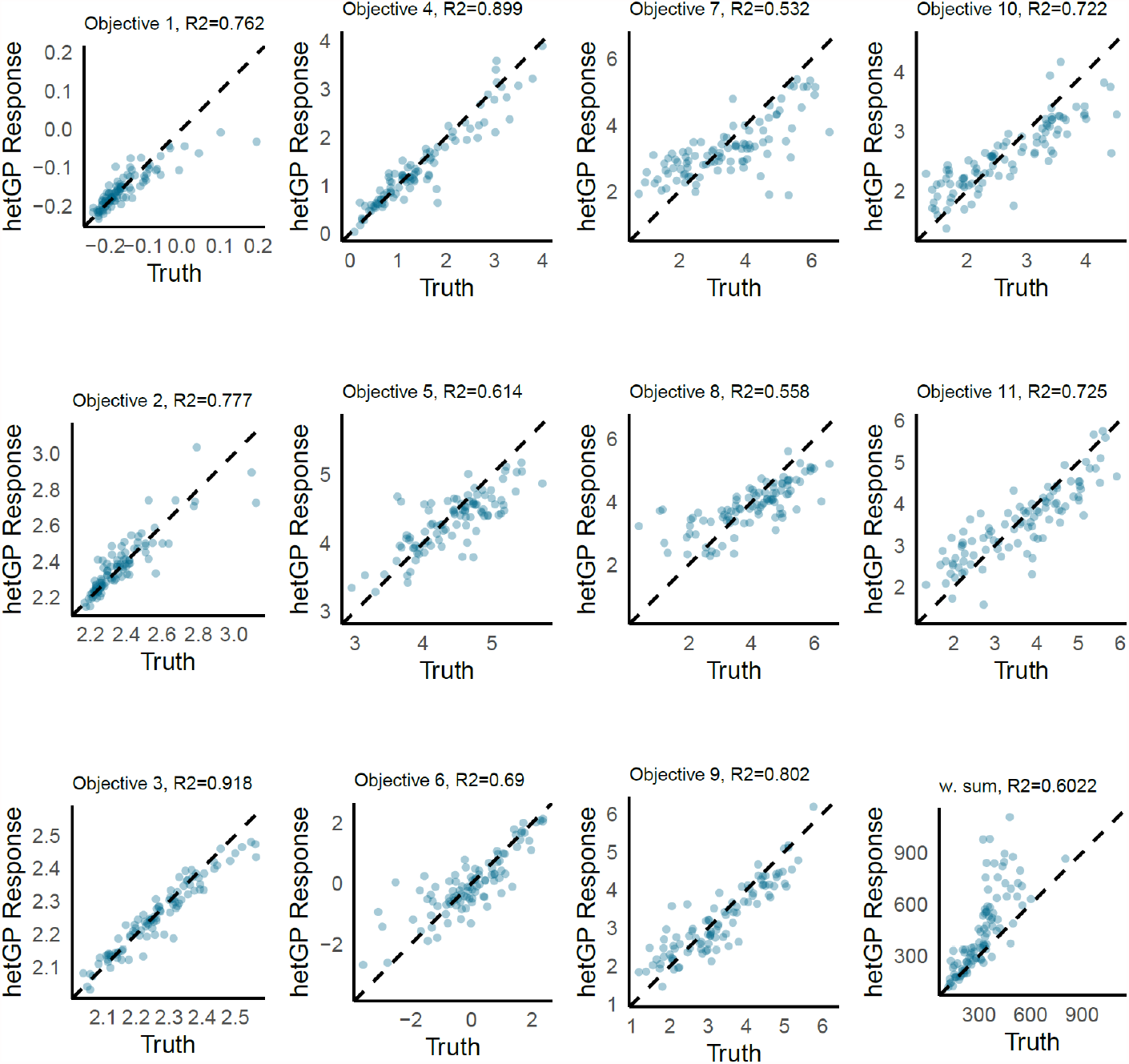
GP emulator performance. Emulator predictions vs true values on a holdout set compromising 10% of initial samples in iteration 30 (final iteration). w.sum is the weighted sum *F*, of the 11 objectives. Here, predictions are generated as the weighted sum of individual objective predictions.

**Figure S4.**
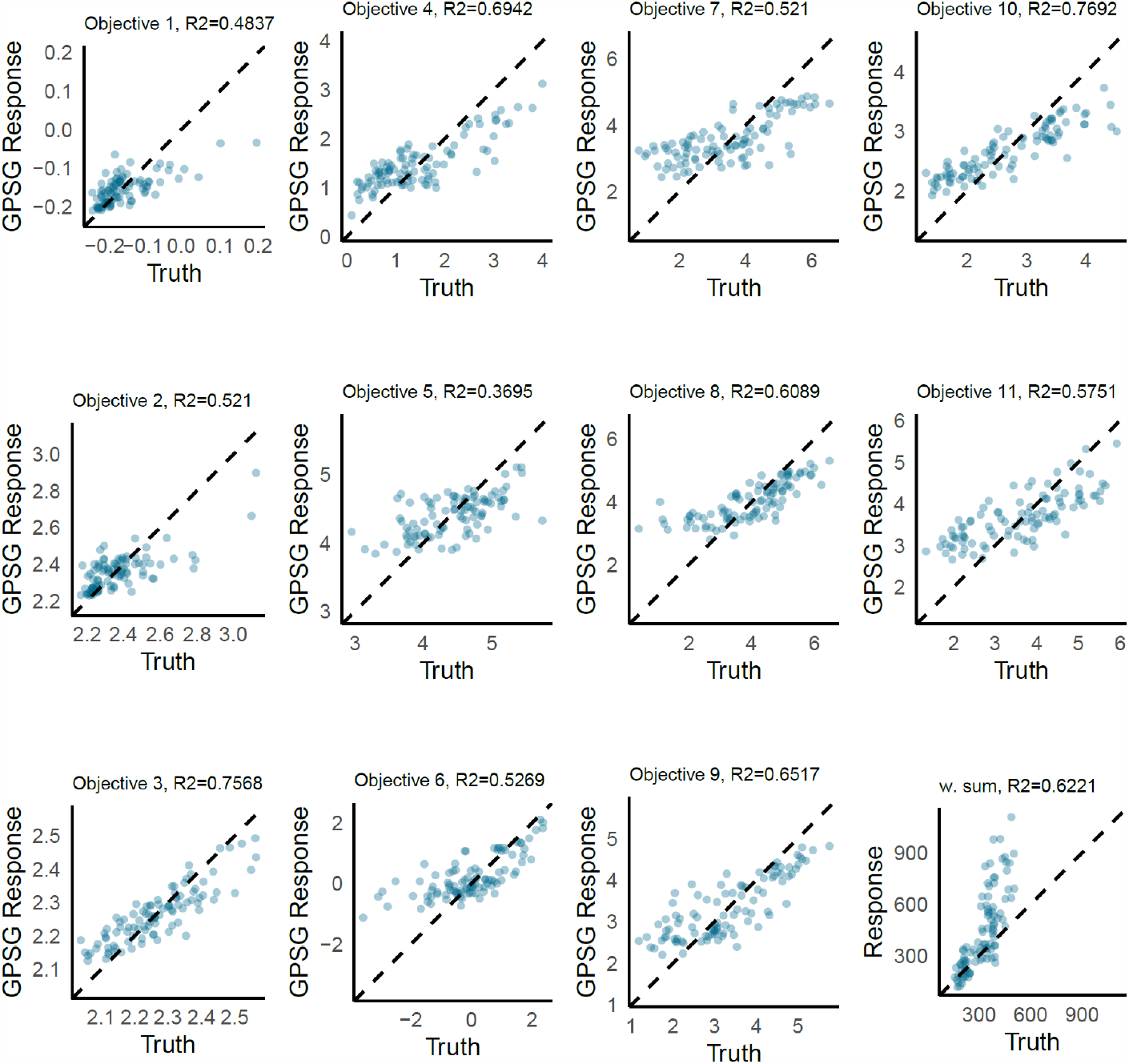
GPSG emulator performance. Emulator predictions vs true values on a holdout set compromising 10% of initial samples in iteration 1. w.sum is the weighted sum *F*, of the 11 objectives. Here, predictions are generated as the weighted sum of individual objective predictions.

**Figure S5.**
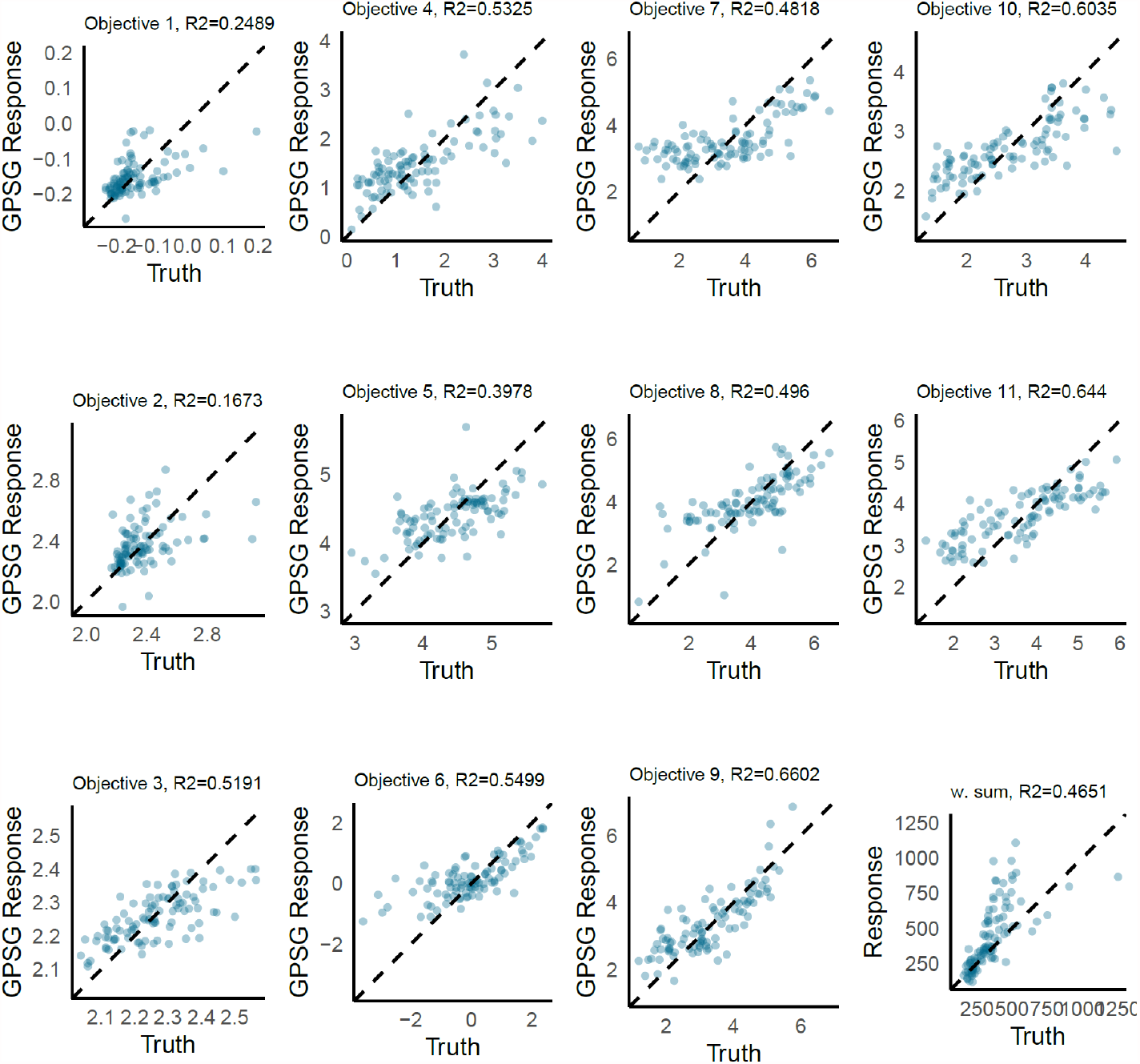
GPSG emulator performance. Emulator predictions vs true values on a holdout set compromising 10% of initial samples in iteration 23 (final iteration). w.sum is the weighted sum *F*, of the 11 objectives. Here, predictions are generated as the weighted sum of individual objective predictions.

## 4 ADAPTIVE SAMPLING: SELECTED POINTS

### 4.1 GP-BO

**Figure S6.**
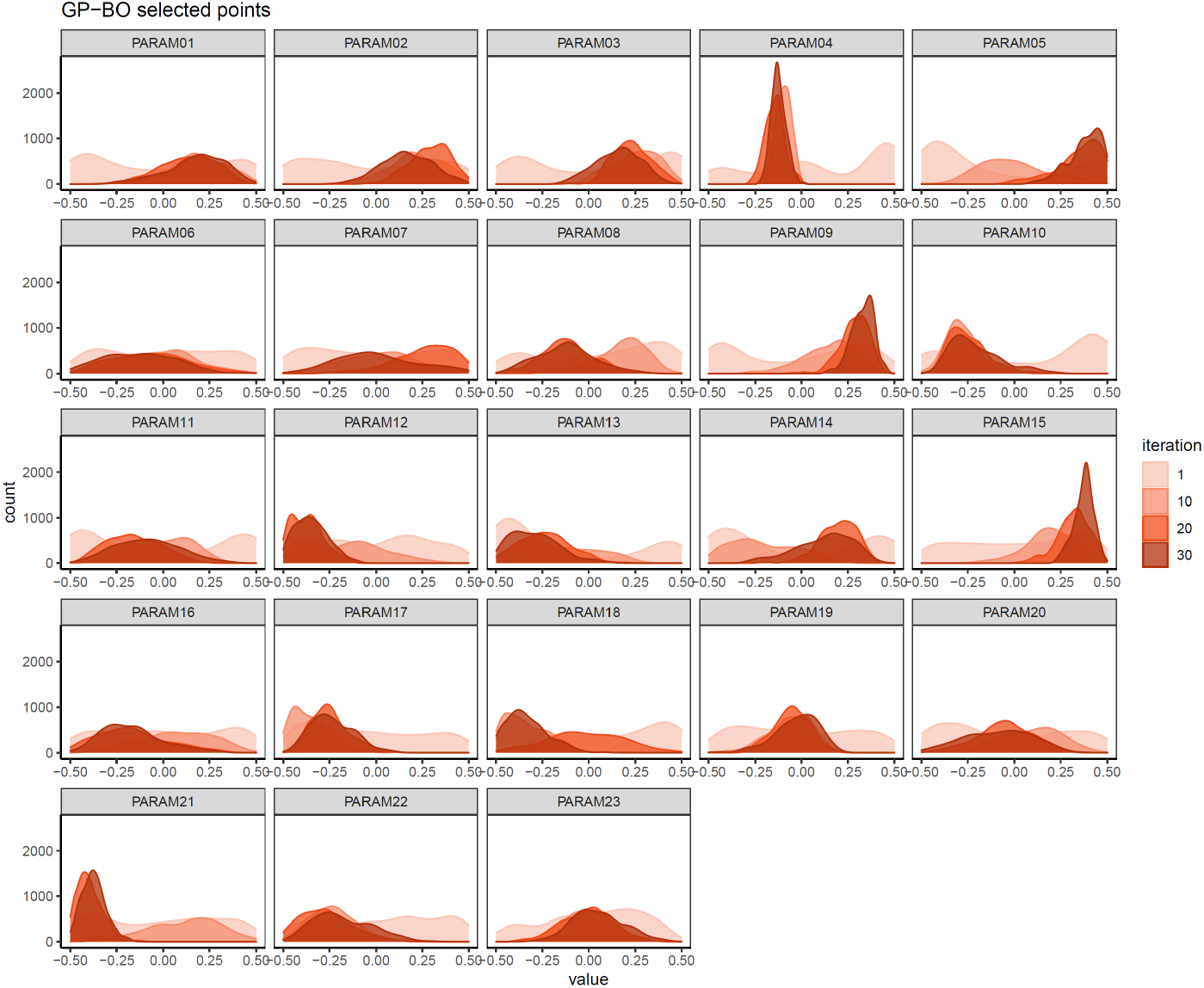
GP-BO sampling behavior. Values in each dimension of the points sampled during adaptive sampling of GP-BO algorithm in iterations 1,10, 20, and 30.

### 4.2 GPSG-BO

**Figure S7.**
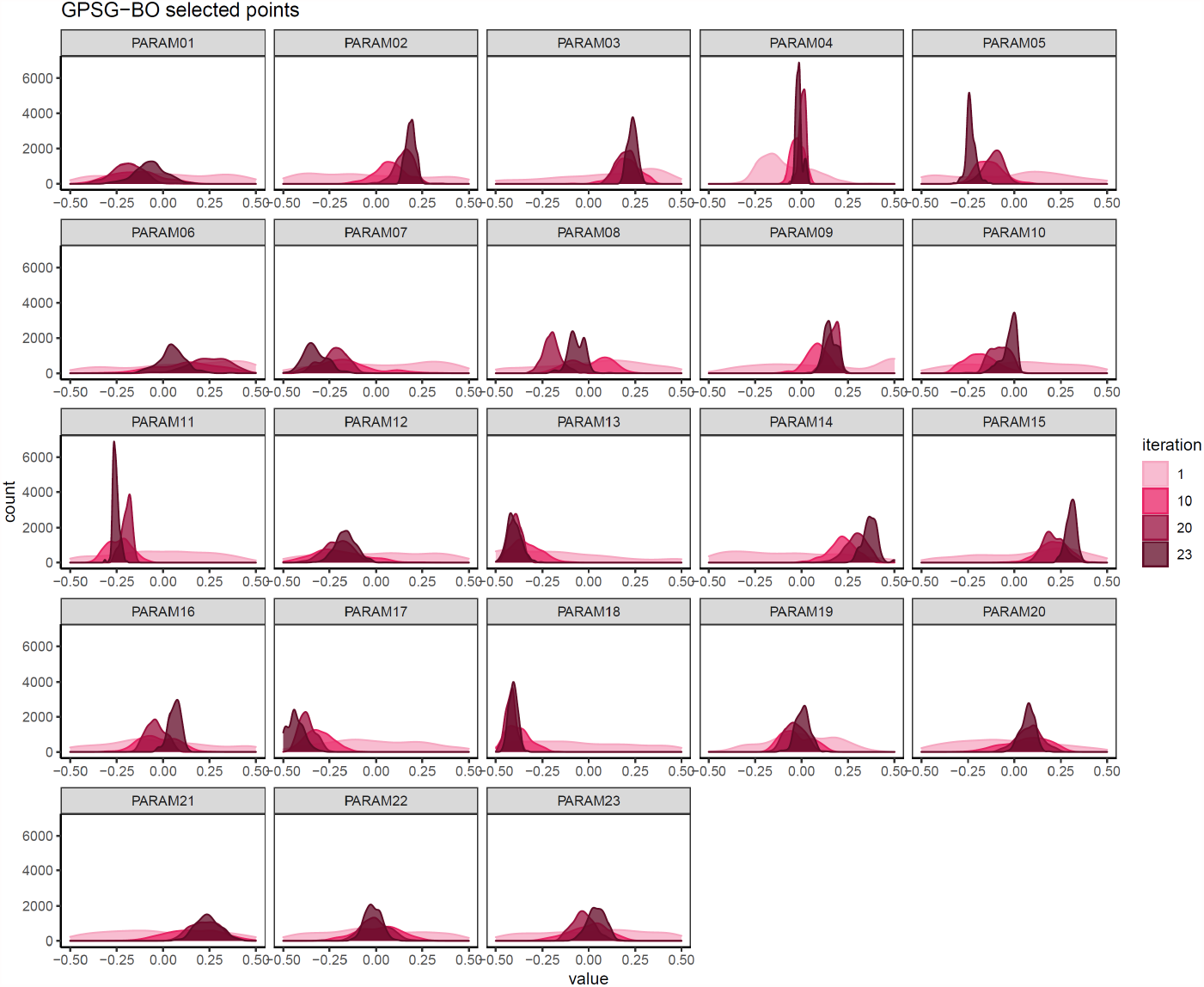
GPSG-BO sampling behavior. Values in each dimension of the points sampled during adaptive sampling of GPSG-BO algorithm in iterations 1,10, 20, and 23.

## 5 OPENMALARIA: FINAL SIMULATOR FIT

**Figure S8.**
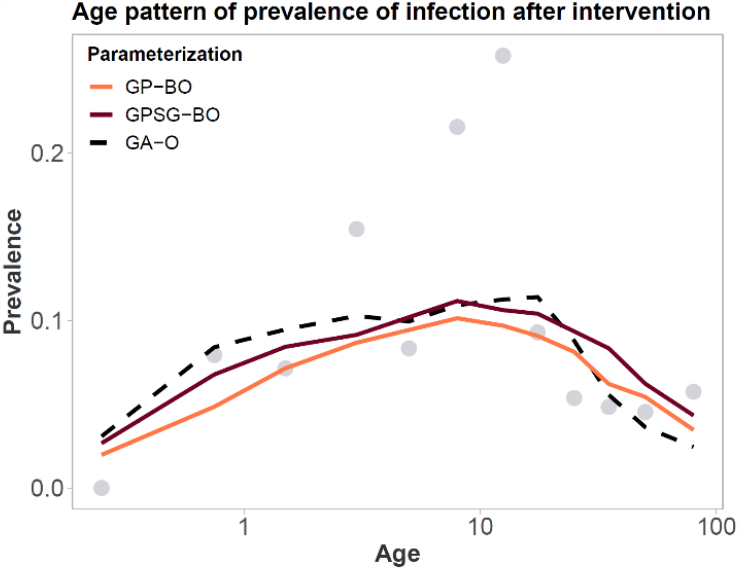
Objective 1: Age pattern of prevalence in Matsari, Nigeria during the intervention. Final simulator fit using the parameter sets yielded using GP-BO and GPSG-BO compared to the previous parameterization (derived using optimization with a genetic algorithm, GA-O).

**Figure S9.**
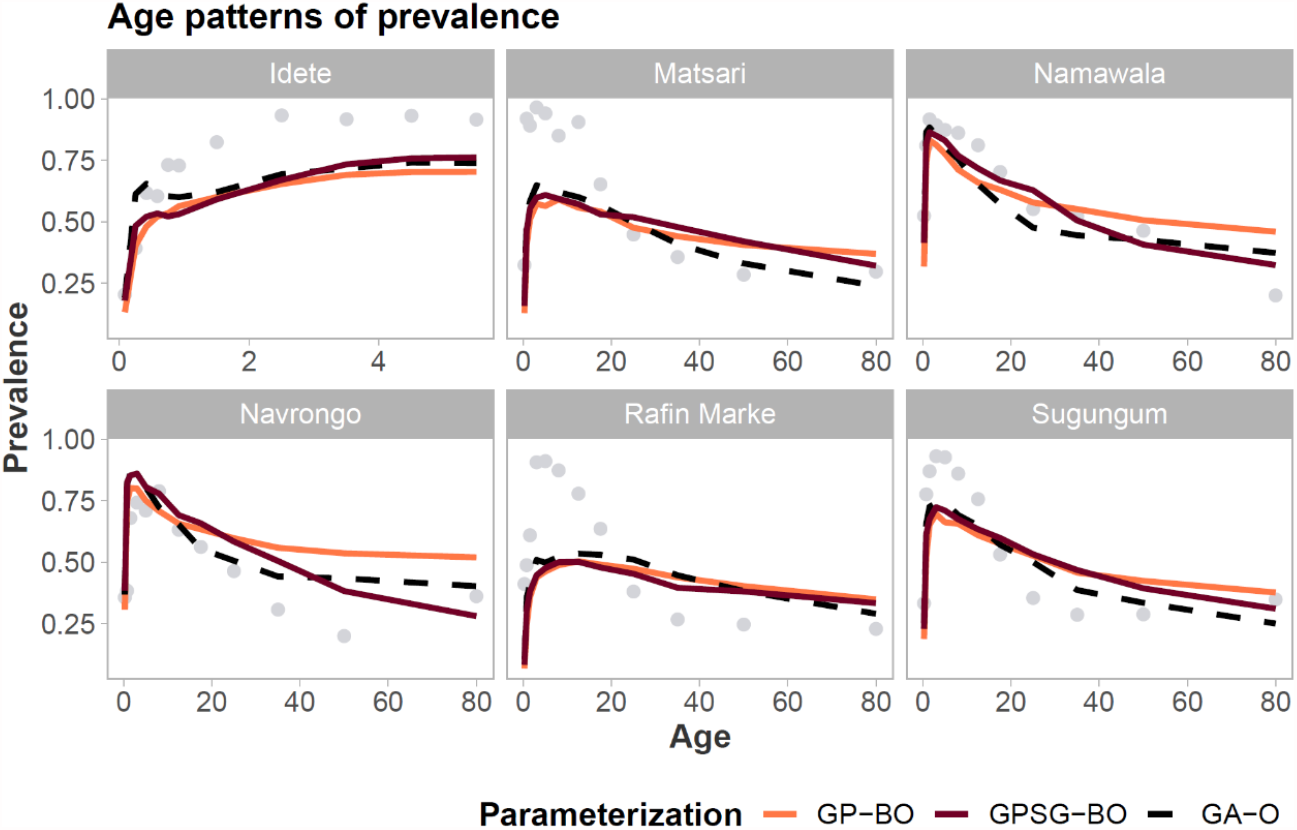
Objective 2: Age pattern of prevalence. Final simulator fit using the parameter sets yielded using GP-BO and GPSG-BO compared to the previous parameterization (derived using optimization with a genetic algorithm, GA-O).

**Figure S10.**
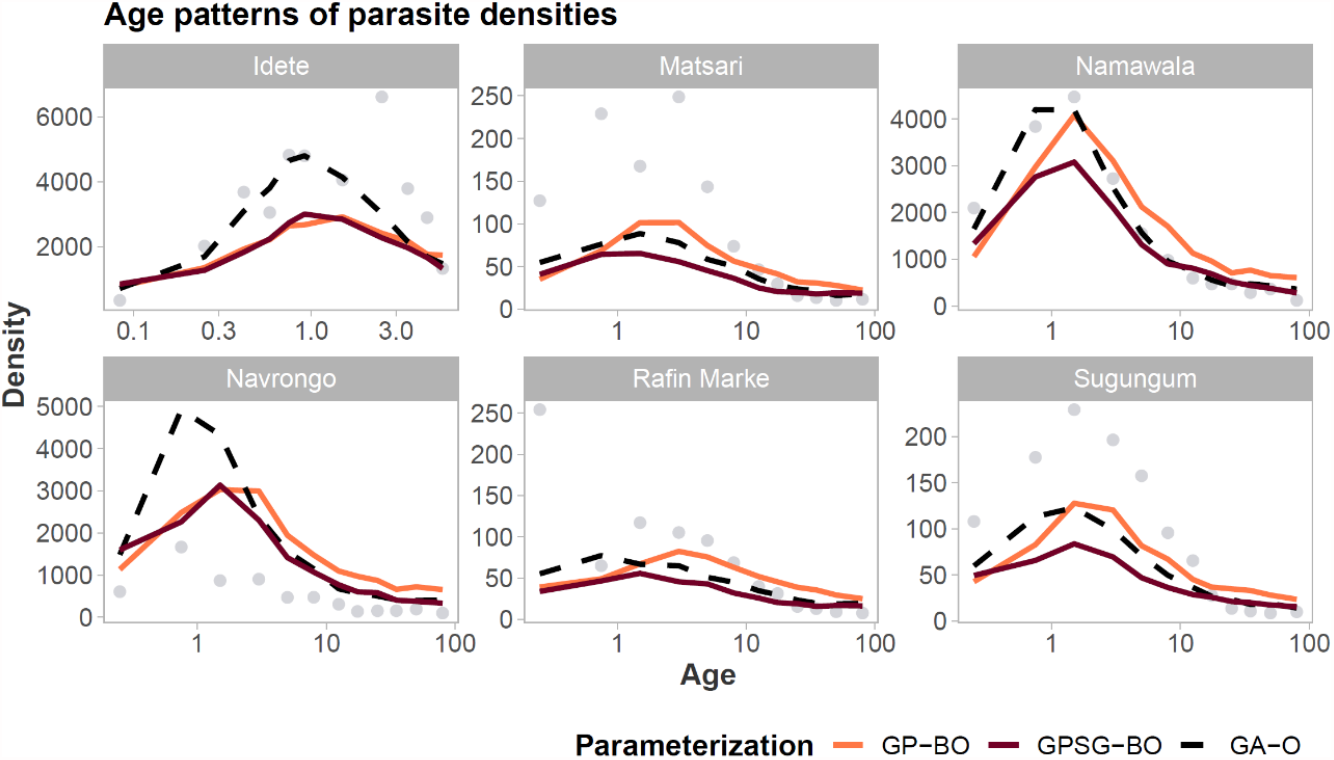
Objective 3: Age pattern of parasite densities (geometric mean). Final simulator fit using the parameter sets yielded using GP-BO and GPSG-BO compared to the previous parameterization (derived using optimization with a genetic algorithm, GA-O).

**Figure S11.**
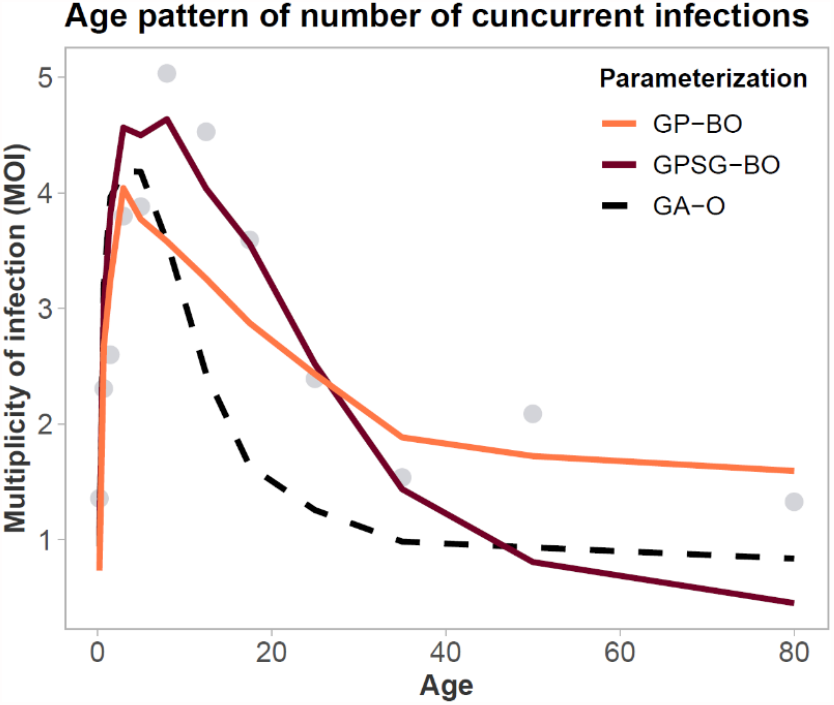
Objective 4: Age pattern of number of concurrent infections. Final simulator fit using the parameter sets yielded using GP-BO and GPSG-BO compared to the previous parameterization (derived using optimization with a genetic algorithm, GA-O).

**Figure S12.**
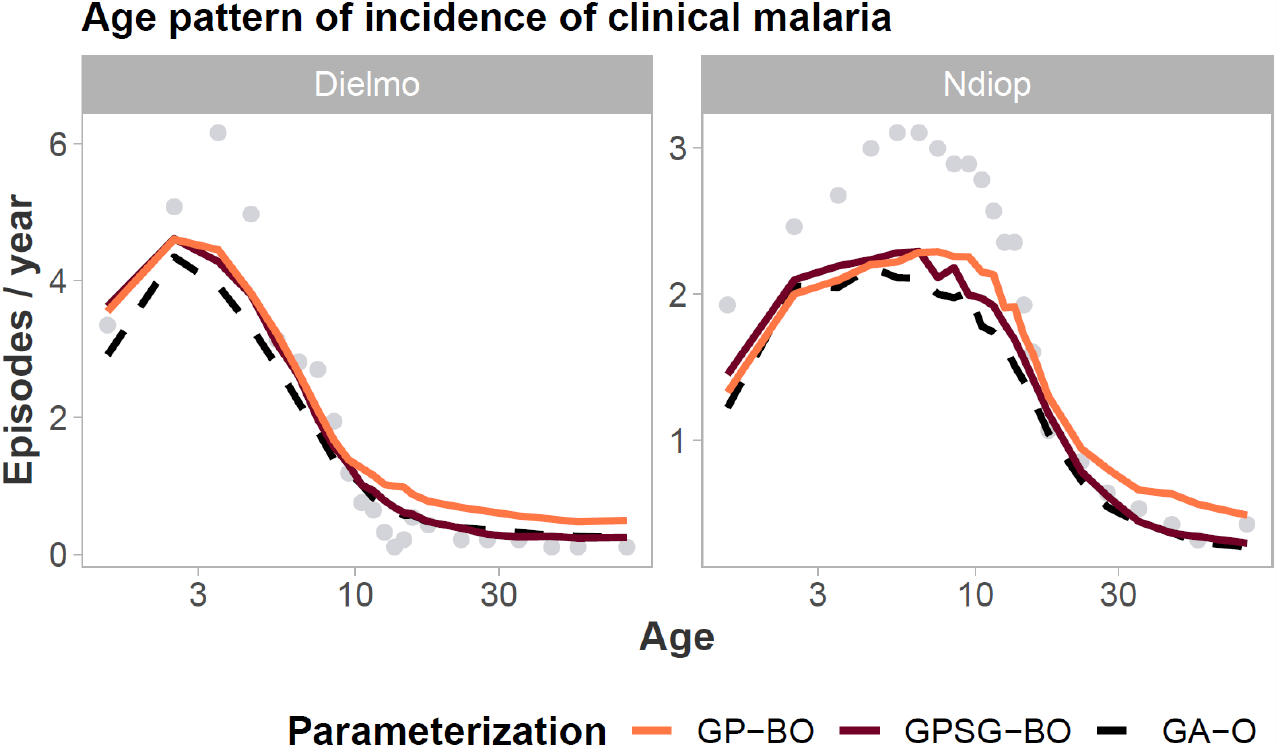
Objective 5: Age pattern of incidence of clinical malaria in Dielmo and Ndiop, Senegal. Final simulator fit using the parameter sets yielded using GP-BO and GPSG-BO compared to the previous parameterization (derived using optimization with a genetic algorithm, GA-O).

**Figure S13.**
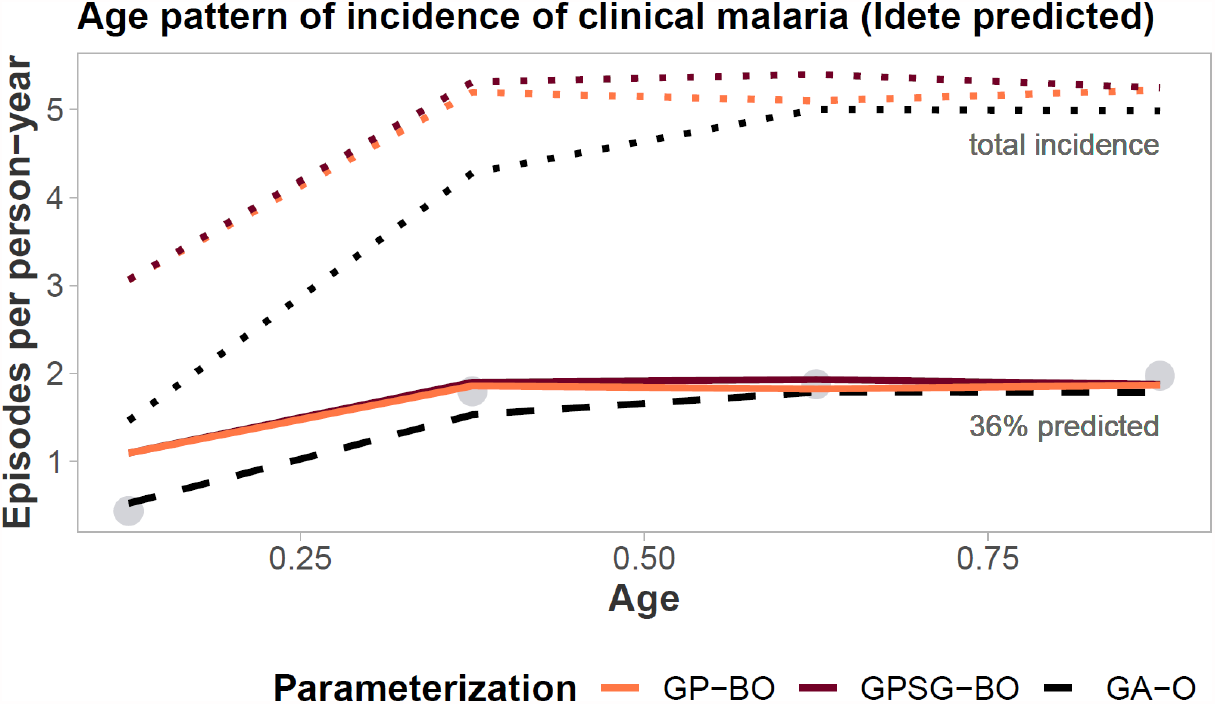
Objective 6: Age pattern of incidence of clinical malaria in Idete, Tanzania. Final simulator fit using the parameter sets yielded using GP-BO and GPSG-BO compared to the previous parameterization (derived using optimization with a genetic algorithm, GA-O).

**Figure S14.**
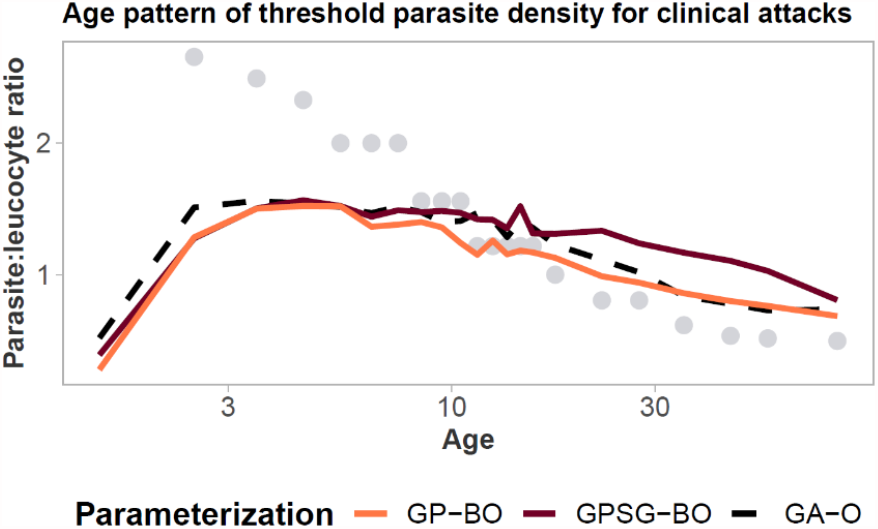
Objective 6. Age pattern of threshold parasite density for clinical attacks. Final simulator fit using the parameter sets yielded using GP-BO and GPSG-BO compared to the previous parameterization (derived using optimization with a genetic algorithm, GA-O).

**Figure S15.**
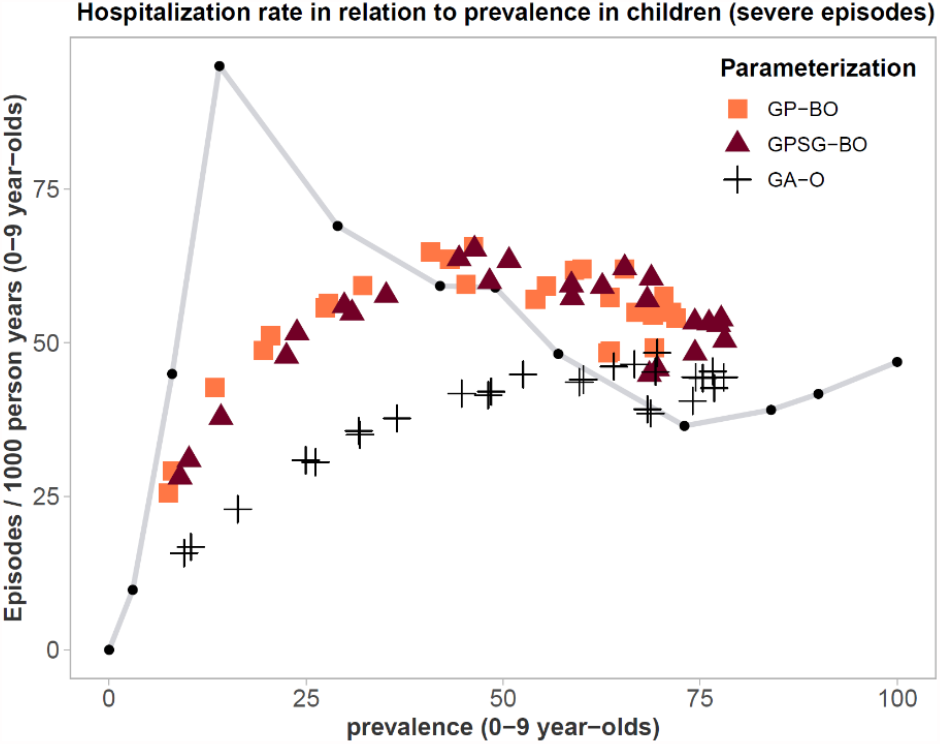
Objective 7: Hospitalization rate in relation to prevalence in children. Final simulator fit using the parameter sets yielded using GP-BO and GPSG-BO compared to the previous parameterization (derived using optimization with a genetic algorithm, GA-O).

**Figure S16.**
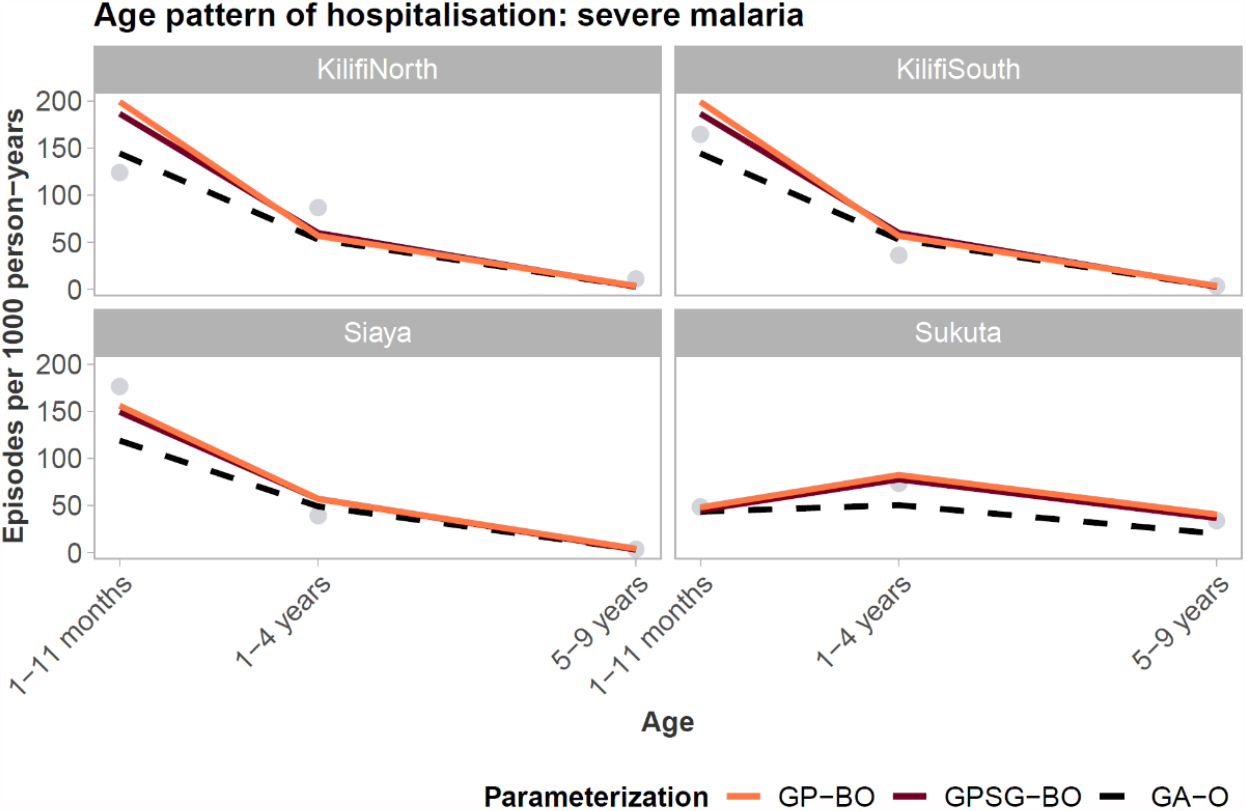
Objective 8. Age pattern of hospitalization. Final simulator fit using the parameter sets yielded using GP-BO and GPSG-BO compared to the previous parameterization (derived using optimization with a genetic algorithm, GA-O).

**Figure S17.**
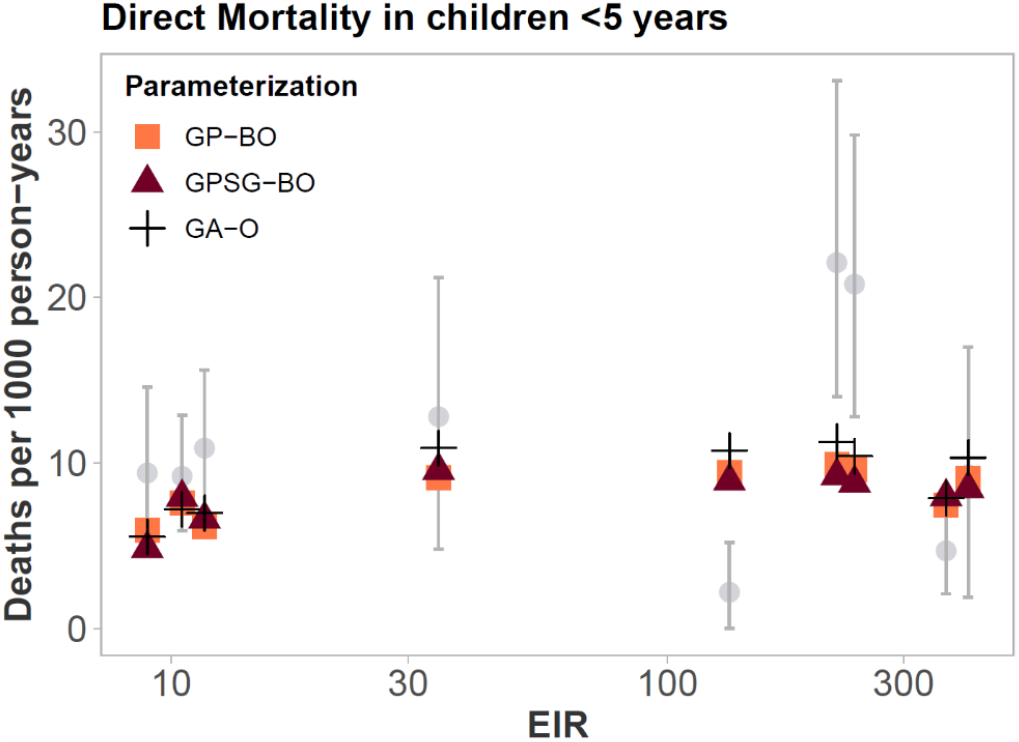
Objective 9: Direct mortality in children <5 years old. Final simulator fit using the parameter sets yielded using GP-BO and GPSG-BO compared to the previous parameterization (derived using optimization with a genetic algorithm, GA-O).

**Figure S18.**
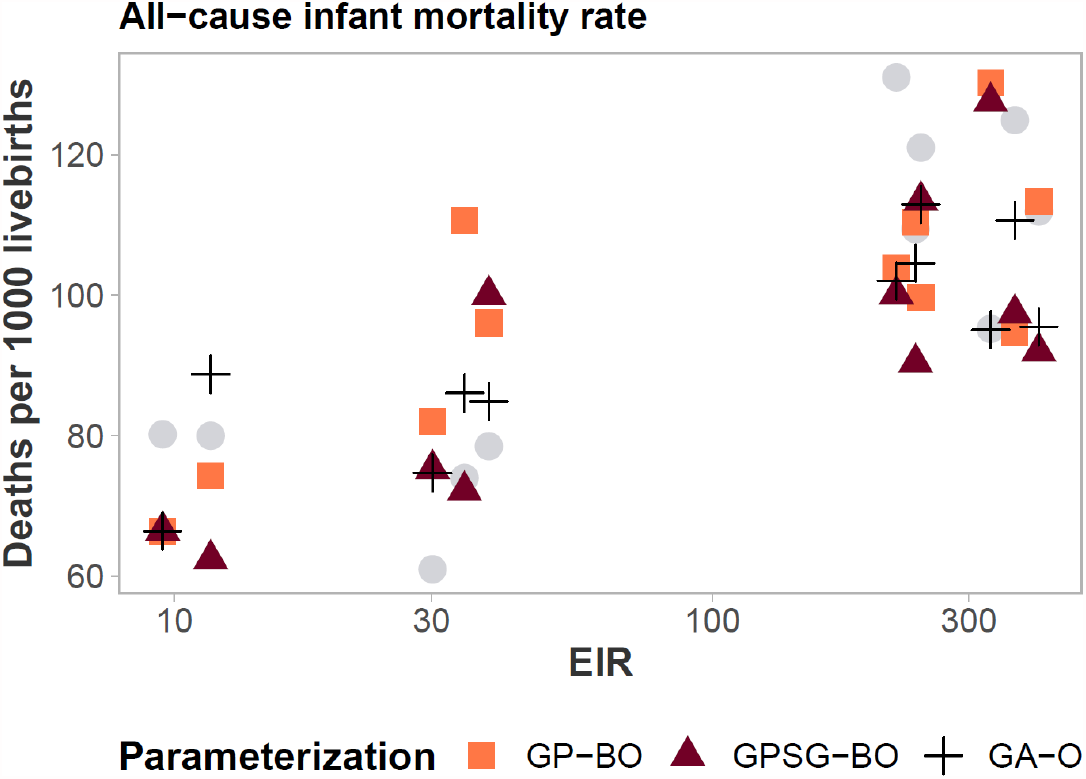
Objective 10: All-cause infant mortality rate. Final simulator fit using the parameter sets yielded using GP-BO and GPSG-BO compared to the previous parameterization (derived using optimization with a genetic algorithm, GA-O).

## 6 VALIDATION

**Figure S19.**
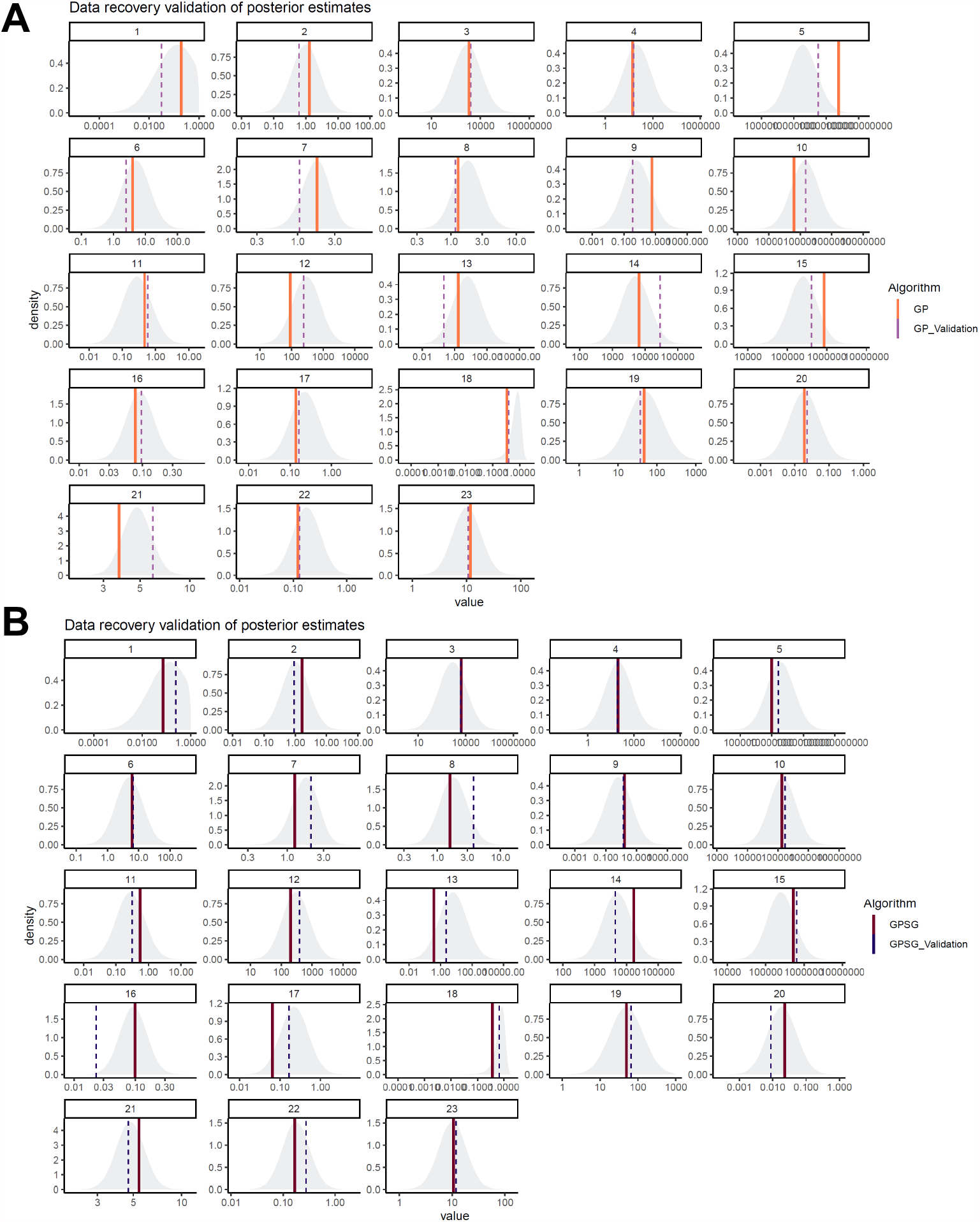
Data recovery validation of posterior estimates. Prior distributions of each parameter and parameter value identified by the optimization algorithm. The final parameter set was used to generate synthetic field data by simulating each of the 61 scenarios with the respective core parameter sets. The simulation outputs were reformatted to match the original field data, generating a synthetic field data set. The optimization with both algorithms was repeated using this synthetic field data. The plot shows the best parameter values in each dimension identified at the end of the validation optimization compared to the values identified in the original optimization. The grey area shows the prior distribution. **A. GP-BO validation. B. GPSG-BO validation**

## 7 OPENMALARIA SIMULATED EPIDEMIOLOGY

**Figure S20.**
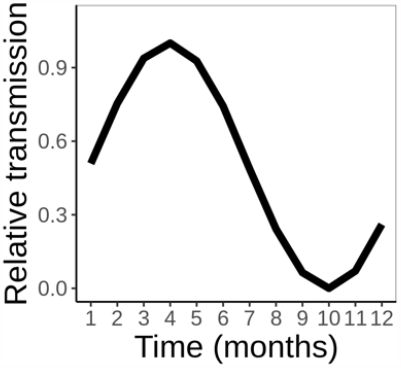
Seasonal pattern assumed for subsequent analyses. The monthly transmission intensity is equivalent to the annual transmission intensity (EIR) scaled by these values and forced to sum to the annual EIR.

**Figure S21.**
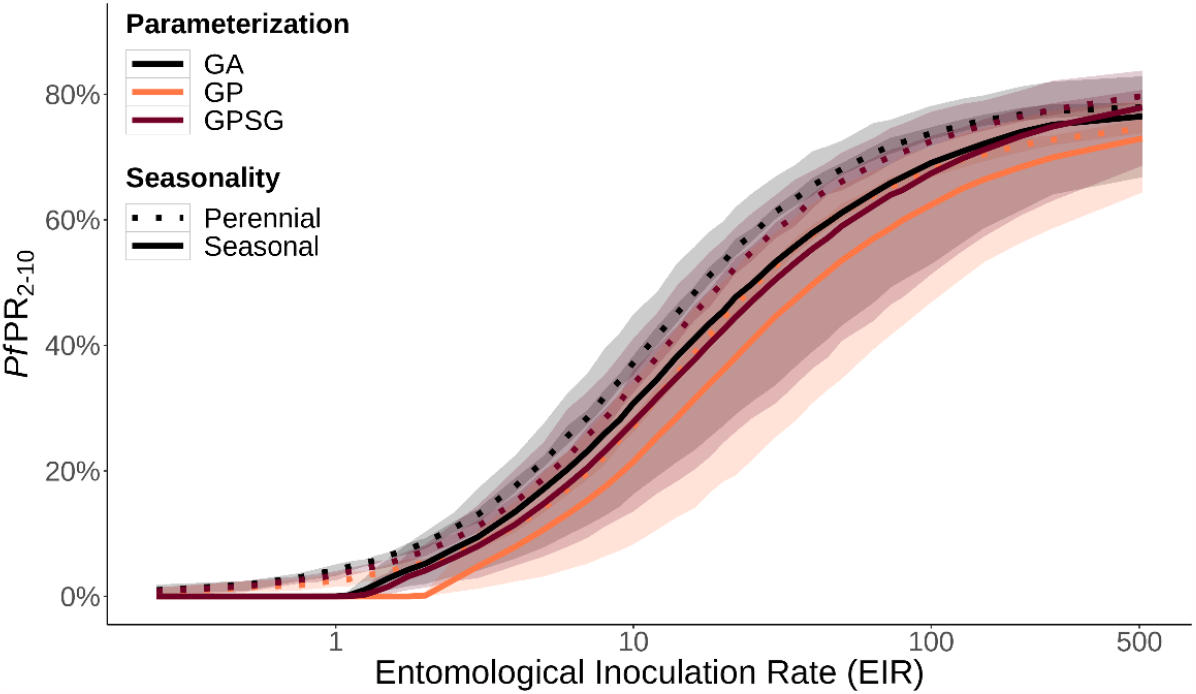
Relationship between EIR and *Pf*PR_2-10_ under three parameterizations. Solid lines show medians and shaded regions show 95% credible intervals. EIR denotes the entomological inoculation rate.

**Figure S22.**
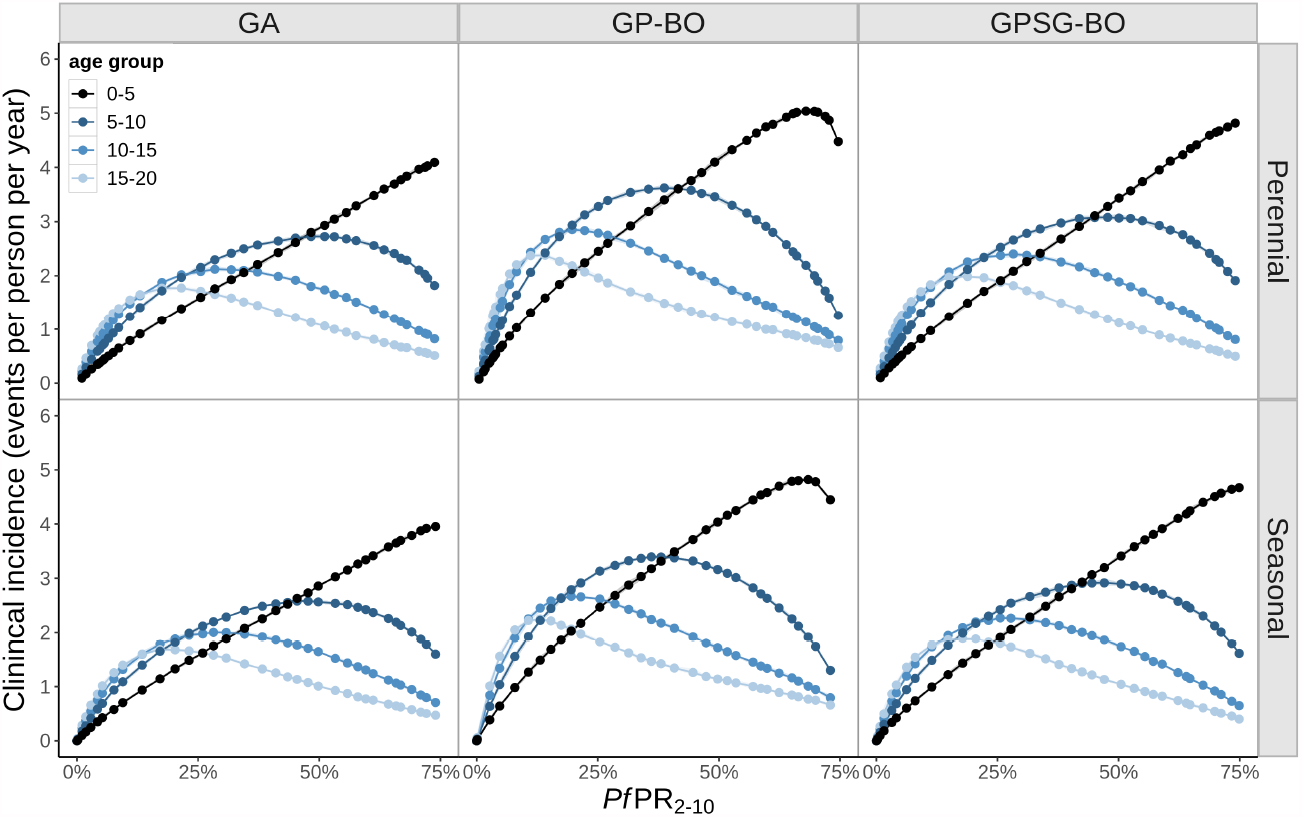
Yearly incidence of clinical (uncomplicated) malaria as a function of PfPR_2-10_ displayed by parameterization and age group. Clinical incidence is presented in terms of the yearly number of events per person. We assume a probability of effective treatment within 14 days of uncomplicated malaria of 36%

**Figure S23.**
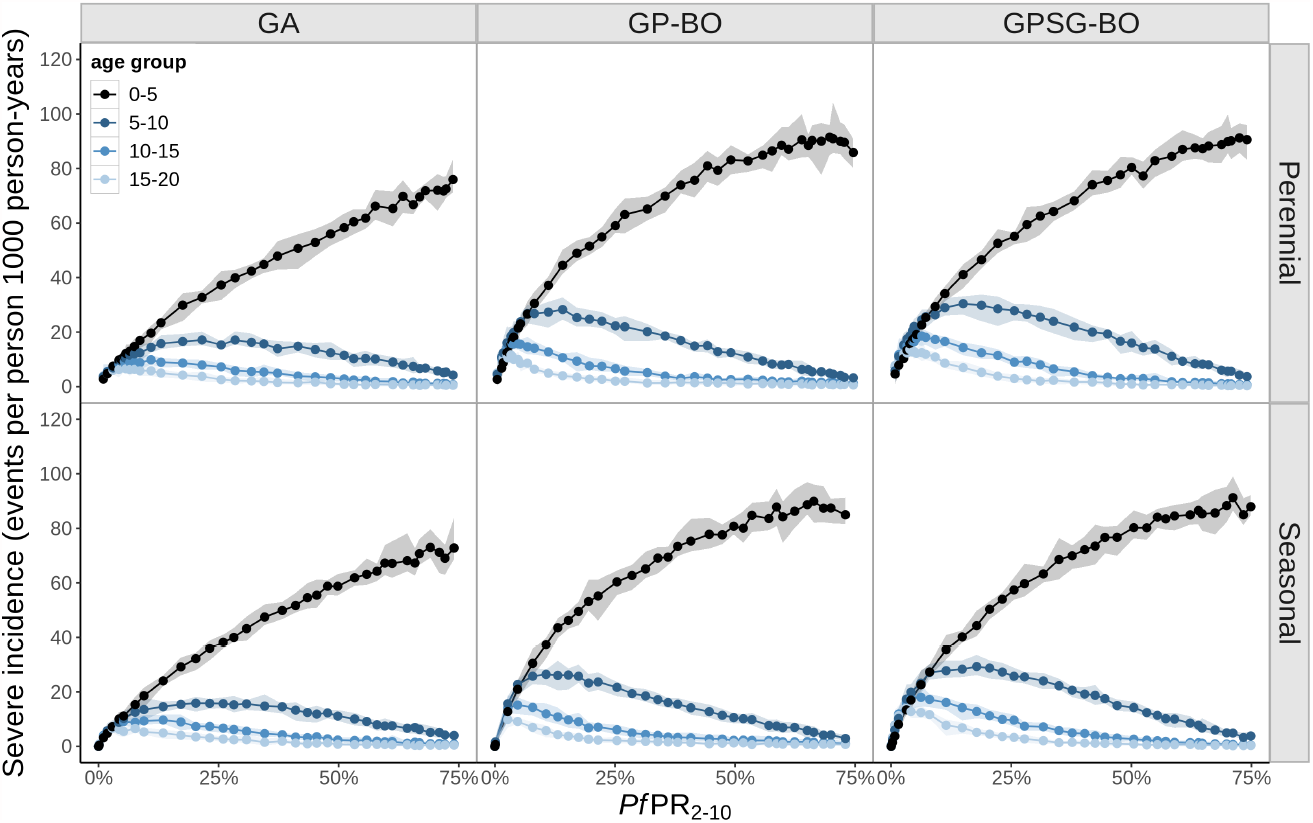
Yearly incidence of total severe malaria as a function of PfPR_2-10_, displayed by parameterization and age group. Incidence is presented in terms of the yearly number of events in a population of 1000 individuals. It is assumed that 48% of severe malaria cases seek official care at a heath care facility (hospital). We assume a probability of effective treatment within 14 days of uncomplicated malaria of 36%

**Figure S24.**
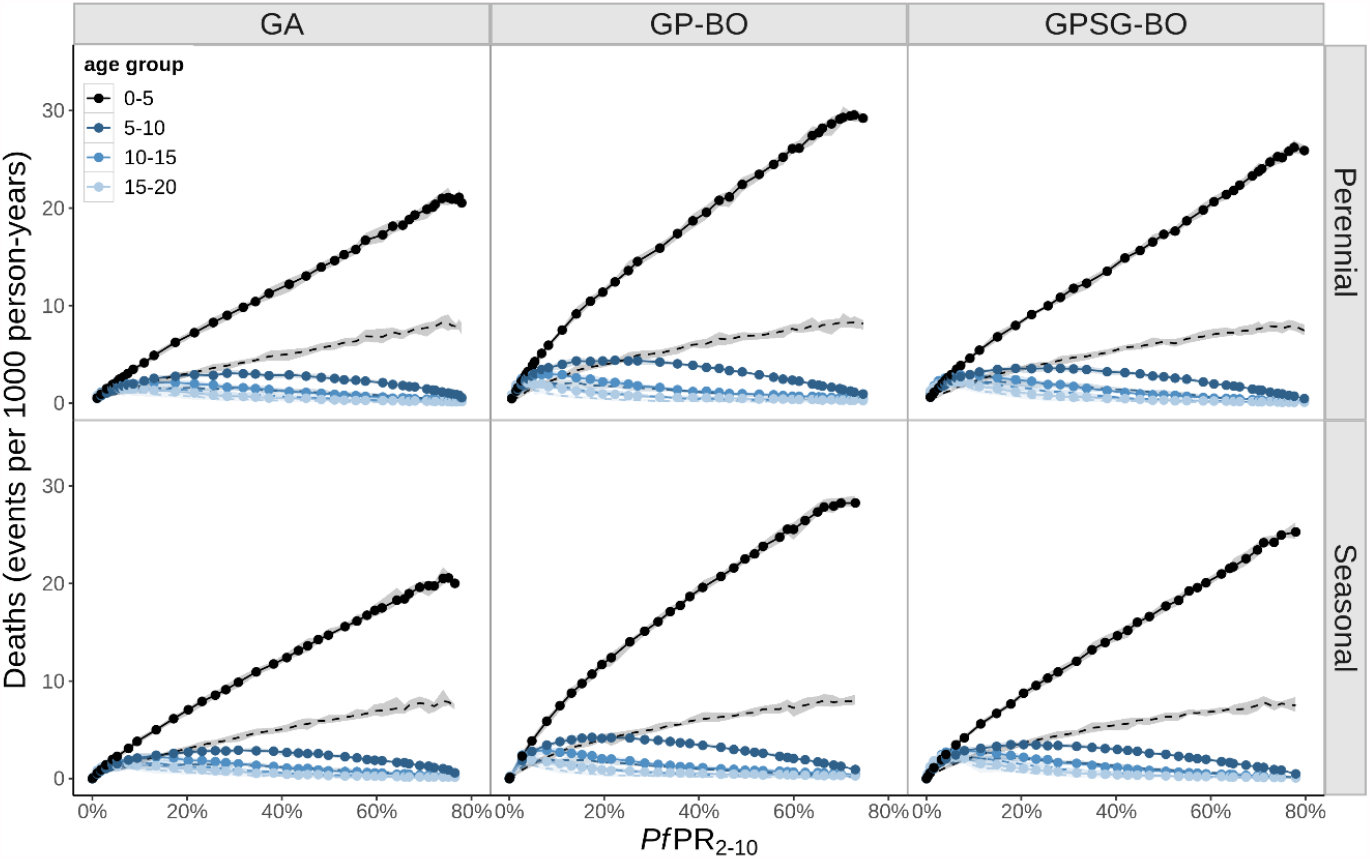
Yearly number of malaria-related deaths as a function of PfPR_2-10_, displayed by parameterization and age group. Malaria mortality incidence is presented in terms of the yearly number of deaths in a population of 1000 individuals. For the OpenMalaria model both deaths directly attributed to malaria (dotted curve) and all deaths associated with malaria (including both deaths directly attributable to malaria and those associated with comorbidities) are shown (full line). See Box S1.2 for definitions of deaths attributable to malaria in the models

**Figure S25.**
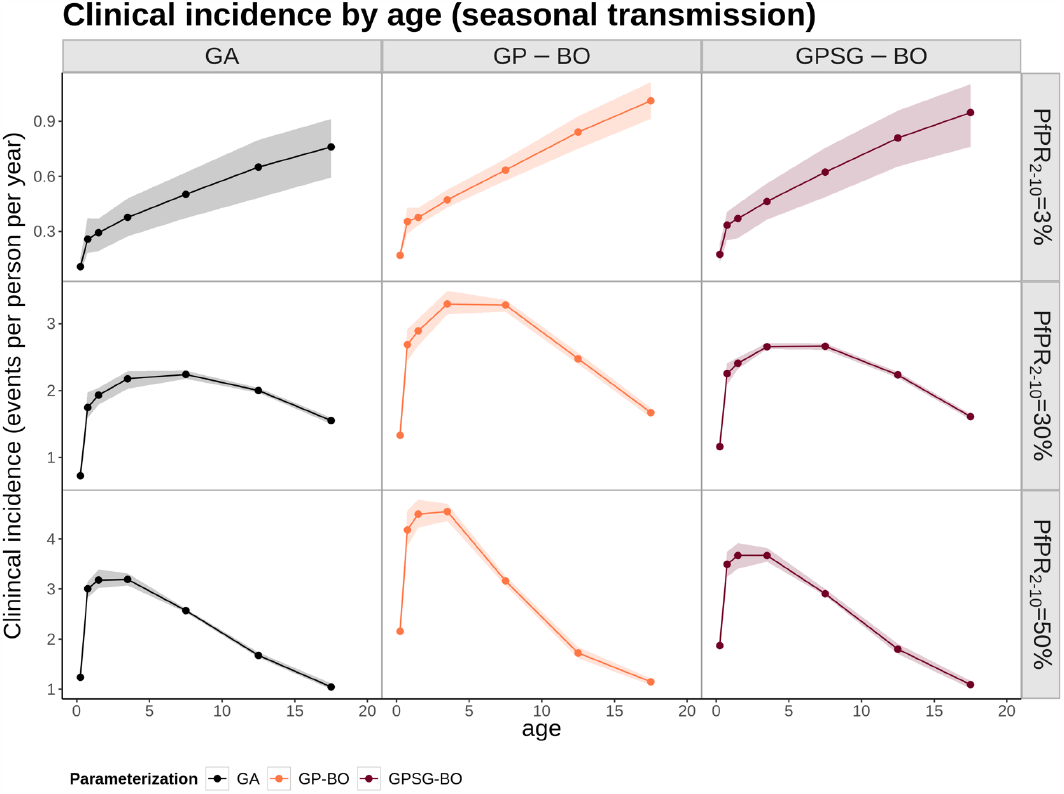
Yearly incidence of clinical malaria in a seasonal transmission setting as a function of age, displayed by transmission intensity (PfPR_2-10_) and parameterization. Clinical incidence is presented in terms of the yearly number of events per person. The PfPR_2-10_ categories include simulated prevalences of 2.5-3.5%, 9-10%, 28-32%, and 47-53% labeled as 3%, 10%, 30%, and 50%, respectively.

**Figure S26.**
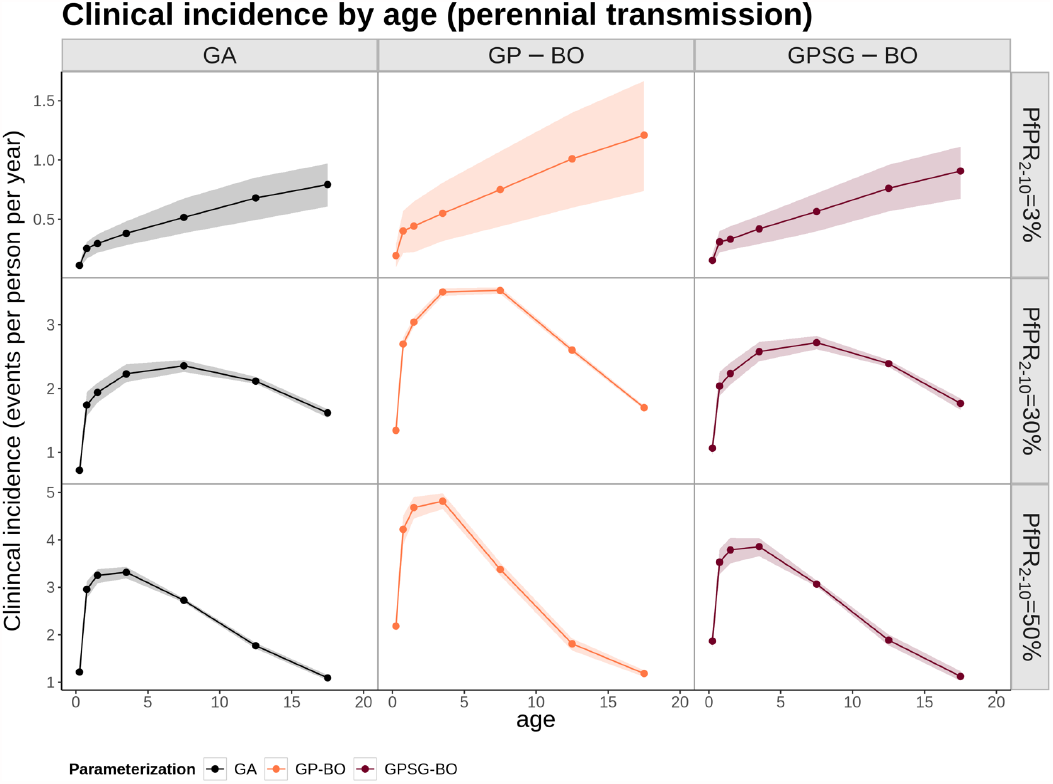
Yearly incidence of clinical malaria in a perennial transmission setting as a function of age, displayed by transmission intensity (PfPR_2-10_) and parameterization. Clinical incidence is presented in terms of the yearly number of events per person. The PfPR_2-10_ categories include simulated prevalences of 2.5-3.5%, 9-10%, 28-32%, and 47-53% labeled as 3%, 10%, 30%, and 50%, respectively

**Figure S27.**
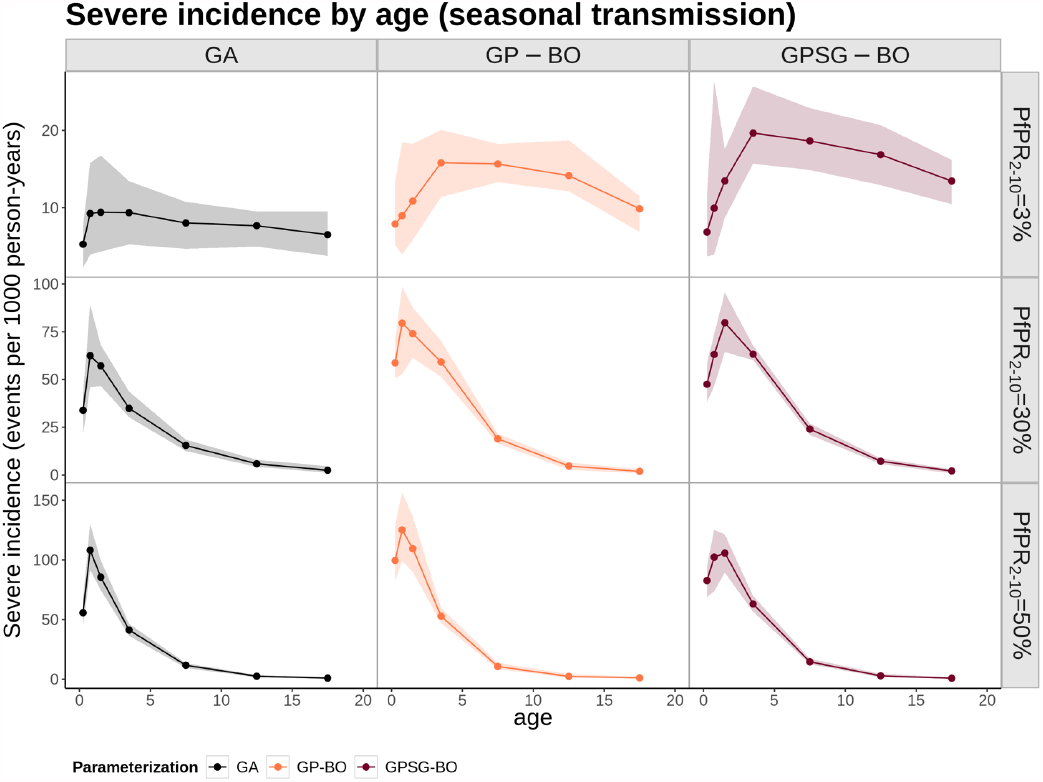
Yearly incidence of total severe malaria in a seasonal transmission setting as a function of age, displayed by transmission intensity (PfPR_2-10_) and parameterization. Incidence is presented in terms of the yearly number of events per 1000 person-years. It is assumed that 48% of severe malaria cases seek official care at a heath care facility (hospital). The PfPR_2-10_ categories include simulated prevalences of 2.5-3.5%, 9-10%, 28-32%, and 47-53% labeled as 3%, 10%, 30%, and 50%, respectively

**Figure S28.**
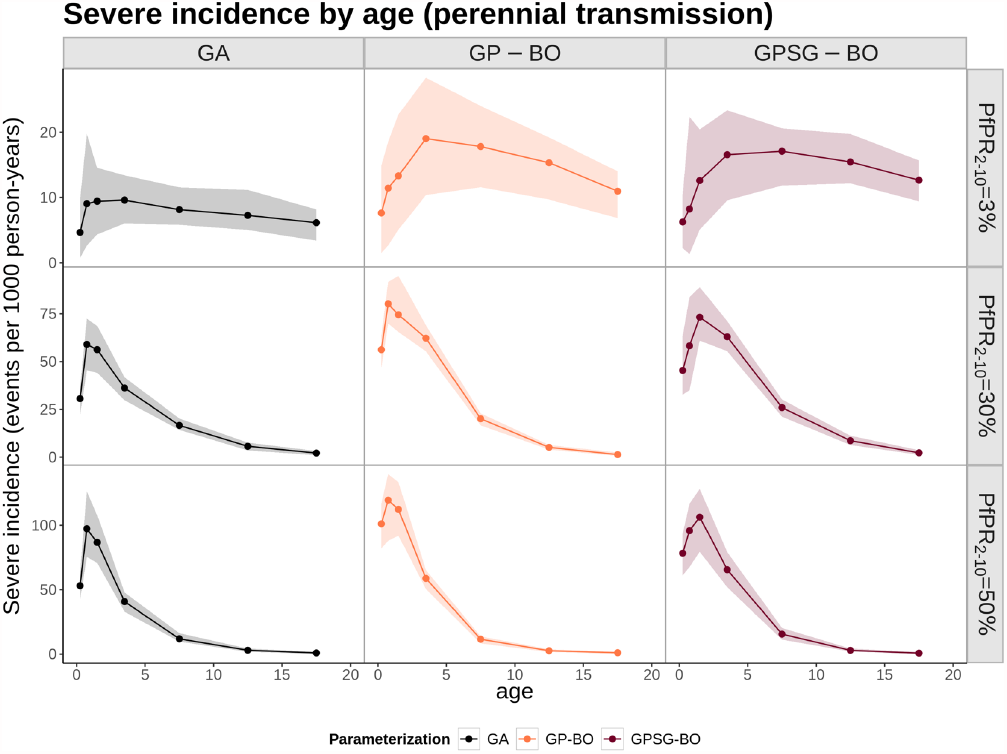
Yearly incidence of total severe malaria in a perennial transmission setting as a function of age, displayed by transmission intensity (PfPR_2-10_) and parameterization. Incidence is presented in terms of the yearly number of events per 1000 person-years. It is assumed that 48% of severe malaria cases seek official care at a heath care facility (hospital). The PfPR_2-10_ categories include simulated prevalences of 2.5-3.5%, 9-10%, 28-32%, and 47-53% labeled as 3%, 10%, 30%, and 50%, respectively

**Figure S29.**
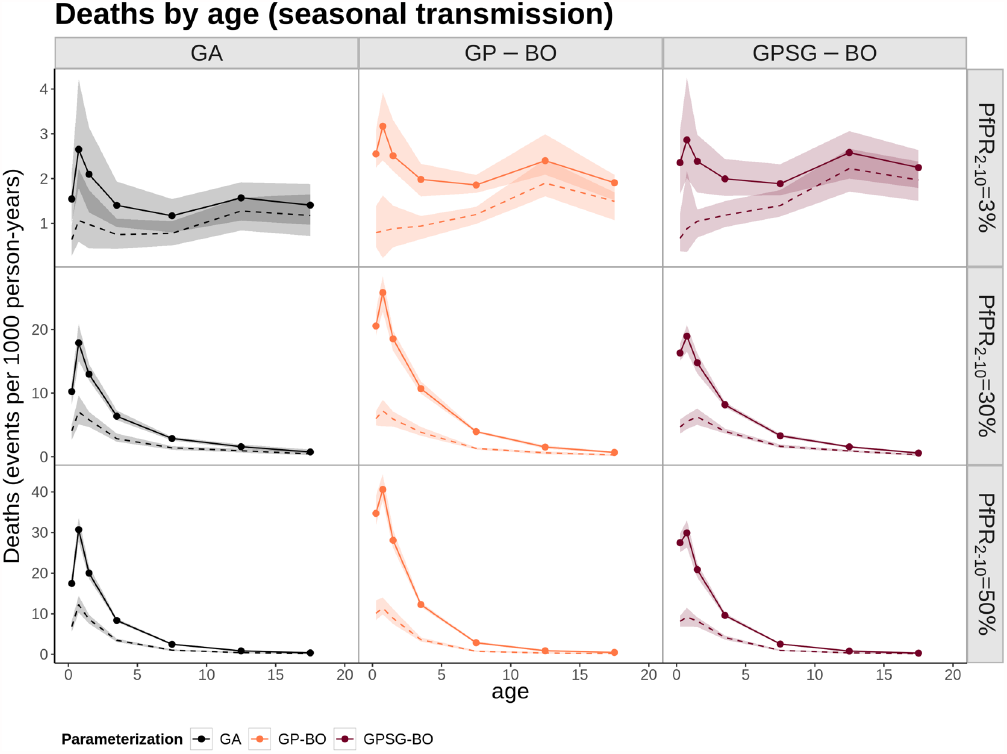
Yearly incidence of malaria-related deaths in a seasonal transmission setting as a function of age, displayed by transmission intensity (PfPR_2-10_) and parameterization. Malaria mortality incidence is presented in terms of the yearly number of deaths in a population of 1000 individuals. The dashed estimates represent direct malaria deaths, and the solid all malaria deaths (including those attributable to co-morbidities). The PfPR_2-10_ categories include simulated prevalences of 2.5-3.5%, 9-10%, 28-32%, and 47-53% labeled as 3%, 10%, 30%, and 50%, respectively

**Figure S30.**
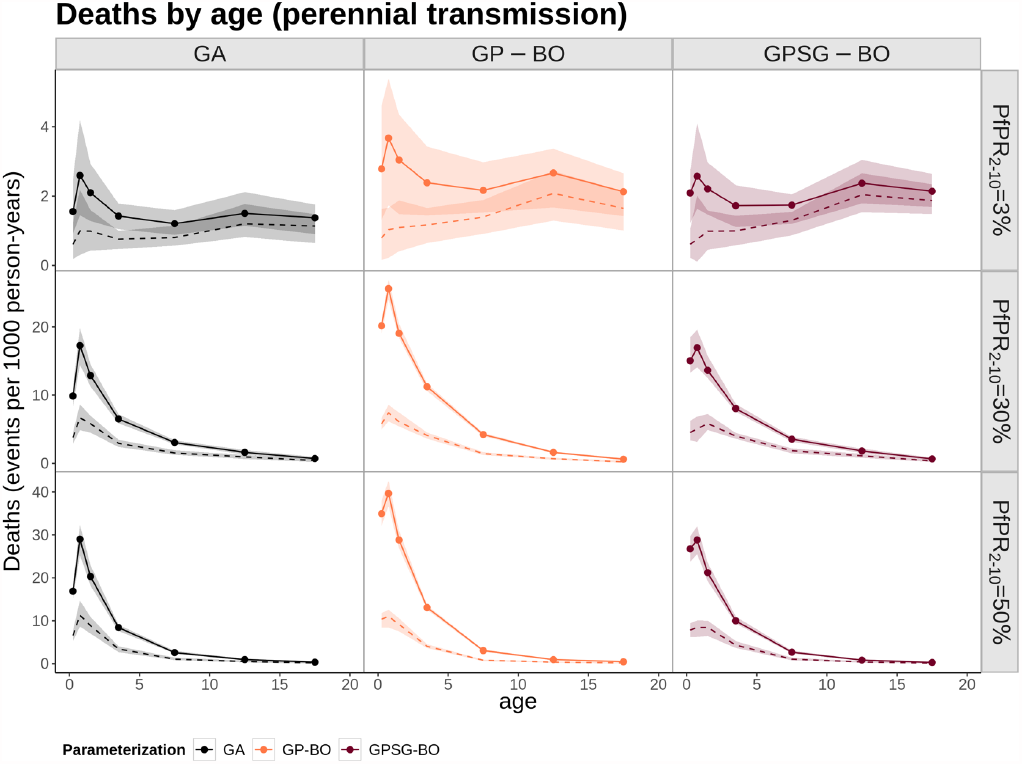
Yearly incidence of malaria-related deaths in a perennial transmission setting as a function of age, displayed by transmission intensity (PfPR_2-10_) and parameterization. Malaria mortality incidence is presented in terms of the yearly number of deaths in a population of 1000 individuals. The dashed estimates represent direct malaria deaths, and the solid all malaria deaths (including those attributable to co-morbidities). The PfPR_2-10_ categories include simulated prevalences of 2.5-3.5%, 9-10%, 28-32%, and 47-53% labeled as 3%, 10%, 30%, and 50%, respectively

## 8 LOG PRIOR DISTRIBUTIONS

**Figure S31.**
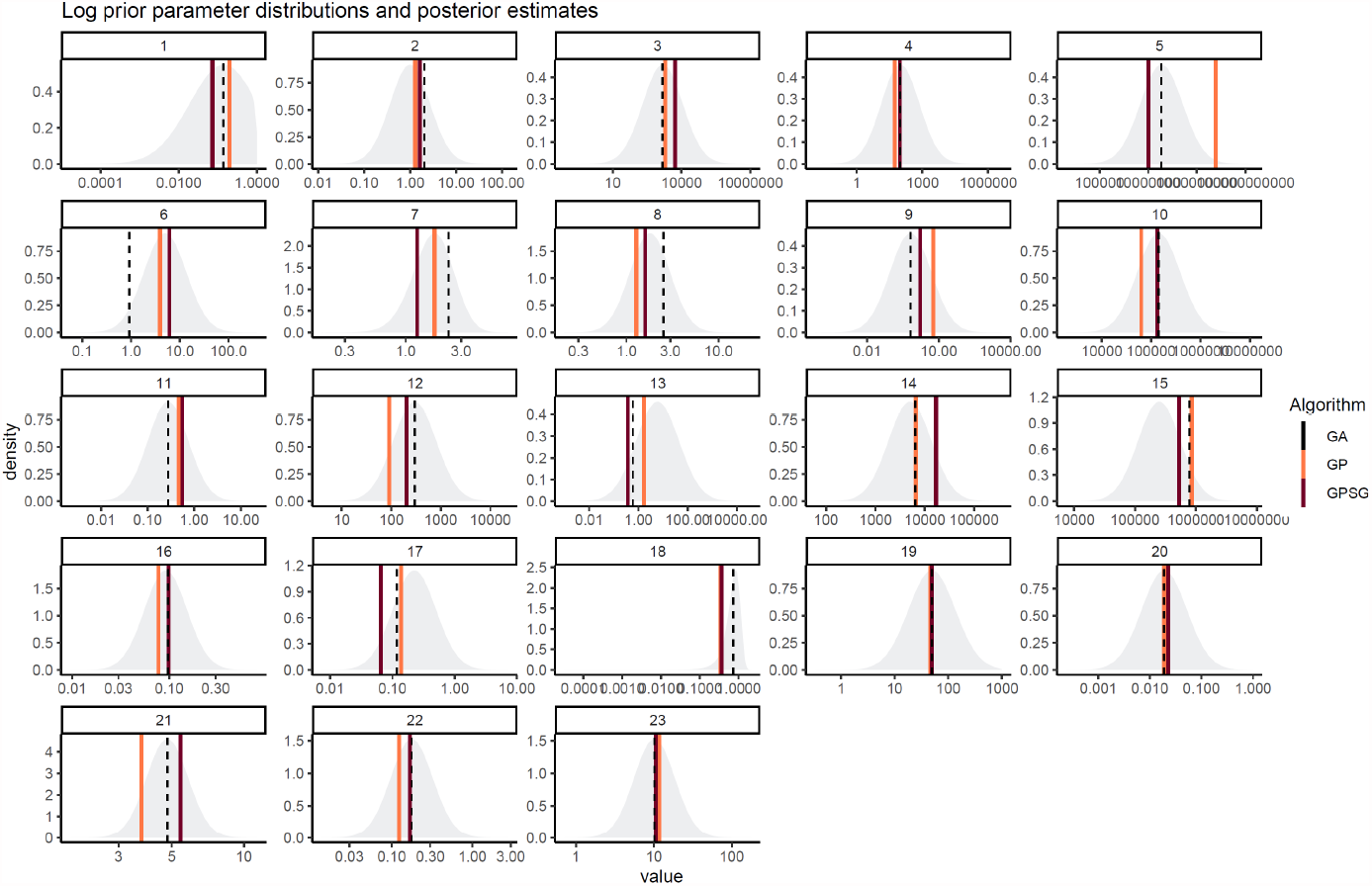
Log prior distributions and final posterior estimates. Prior distributions of each parameter and final parameter values identified by each optimization algorithm (GP-BO and GPSG-BO) and compared to the current parameterization (derived using a genetic algorithm, GA).

## 9 RANGER IMPORTANCE

**Figure S32.**
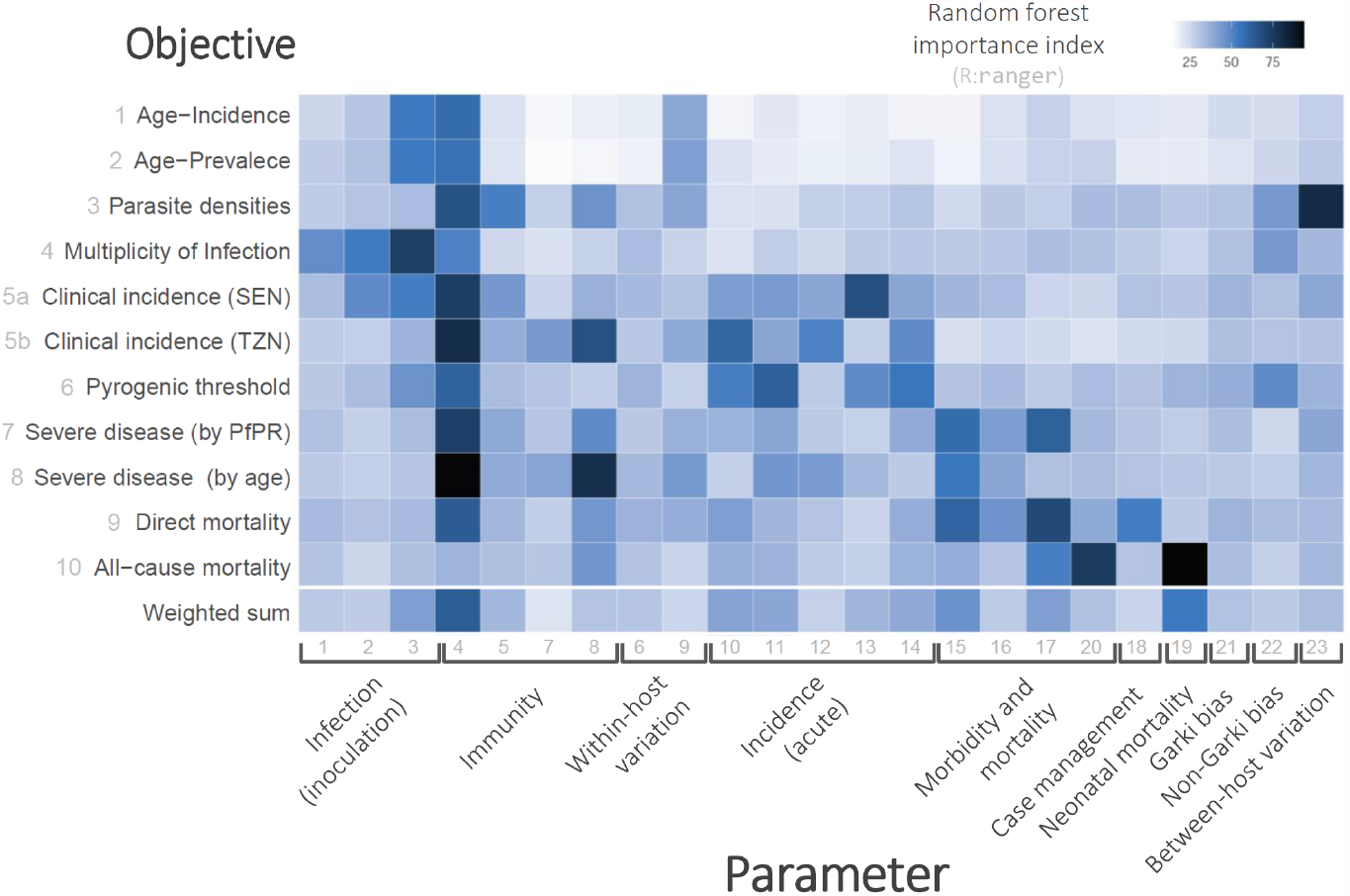
Random forest importance. Estimated parameter importance indices for all parameters and objectives. The indices were calculated using the ranger random forest package in R.

